# Individualized exercise in chronic non-specific low back pain: a systematic review with meta-analysis on the effects of exercise alone or in combination with psychological interventions on pain and disability

**DOI:** 10.1101/2021.12.16.21267900

**Authors:** Johannes Fleckenstein, Philipp Flössel, Tilman Engel, Laura Klewinghaus, Josefine Stoll, Martin Behrens, Daniel Niederer

**Author notes:** **Corresponding author:** Johannes Fleckenstein, Goethe-University Frankfurt, Institute of Sports Sciences, Department of Sports Medicine and Exercise Physiology, Ginnheimer Landstr. 39, 60487 Frankfurt am Main, Germany, mail. **Contributorship**. All authors contributed to the planning of the study. JF, PF and DN conducted the study, JF wrote and DN analysed data, JF wrote the first draft of the manuscript. All authors read, revized and approved the manuscript. JF accepts full responsibility for the work and/or the conduct of the study, has access to the data, and controlled the decision to publish. The corresponding author attests that all listed authors meet authorship criteria and that no others meeting the criteria have been omitted. **Funding** This research did not receive any specific grant from funding agencies in the public, commercial, or not-for-profit sectors.

## Abstract

This systematic review with meta-analysis and meta-regression investigated the effects of individualized exercise interventions consisting with or without combined psychological intervention on pain intensity and disability in patients with chronic non-specific low-back-pain. Databases were searched up to 31 January 2022 and we selected randomized controlled trials involving adults with chronic non-specific low-back-pain being treated with individualized/personalized/stratified exercise interventions with or without psychological treatment compared to any control.

Fifty-eight studies (n = 10084) were included.

At short-term follow-up (12 weeks), low-certainty evidence for pain intensity (SMD -0.28 [95%CI -0.42 to -0.14]) and very low-certainty evidence for disability (−0.17 [-0.31 to -0.02]) indicates effects of individualized versus active exercises, and very low-certainty evidence for pain intensity (−0.40; [-0.58 to -0.22])), but not (low-certainty evidence) for disability (−0.18; [-0.22 to 0.01]) compared to passive controls.

At long-term follow-up (1 year), moderate-certainty evidence for pain intensity (−0.14 [-0.22 to -0.07]) and disability (−0.20 [-0.30 to -0.10]) indicates effects versus passive controls.

Sensitivity analyses indicates that the effects on pain, but not on disability (always short-term and versus active treatments) were robust. Pain reduction caused by individualized exercise treatments in combination with psychological interventions (in particular behavioural-cognitive therapies) (−0.28 [-0.42 to -0.14], low certainty) is of clinical importance.

Certainty of evidence was downgraded mainly due to evidence of risk of bias, publication bias and inconsistency that could not be explained.

Individualized exercise can be recommended from a clinical point of view to treat pain and disability in chronic non-specific low-back-pain. Sub-group analysis suggests a combination of individualized exercise (especially motor-control based treatments) with behavioural therapy interventions to booster effects. Certainty of evidence was moderate for long-term follow-up.

**PROSPERO registration:** CRD42021247331

## Introduction

Back pain, in particular chronic non-specific low back pain, is one of the most serious public-health problems worldwide.^21^ Its global median 1-year period prevalence in the adult population is around 37%.^21^ Together with the consequences of low back pain, such as loss of labour productivity or rates of hospital admissions, this high number of involved people results in tremendous direct and indirect costs for societies’ healthcare systems and economies.^64^ For those affected, pain symptoms are often accompanied by functional limitations and decreased performance, substantially deteriorating the quality of their individual and family lives.^57, 63^

Exercise therapy and psychological (especially cognitive behavioural) therapy, both as stand-alone treatments but especially in combination, are considered being effective in the treatment of chronic low back pain.^45^ Still, current research reveals only relatively small and heterogeneous effects when combining exercise and cognitive-behavioural therapies.^19, 22, 25^ This is likely to be attributed to inter-individual heterogeneities in the effects.^33^ Stratified or even individualized/personalized therapies may be the potential key to increase the (group and individual) therapeutic effect.^23^ On a somatic-biological level, patients with low back pain predominantly present deficits in motor control strategies, though varying in its characteristics between individuals.^40, 59^ On a psychosocial level, dysfunctional cognition (fear-avoidance-beliefs, catastrophizing) and dysfunctional pain behaviour (kinesiophobia) are known risk factors for chronification or for a worsening of symptoms in chronic low back pain.^39, 55^

Functional strength/restoration, coordination (Pilates, McKenzie) and stabilization training are, based on meta-analytic evidence on comparative effectiveness, likely to be the most effective types of exercise in the management of chronic non-specific low back pain.^23, 42^ Simply and mechanistically expressed, all these training modalities improve motor control.^59^ Unfortunately, such effects are only of limited validity for the affected individuals due to the presence of heterogeneous influencing psychosocial factors in group analyses.^42^ The latter has been addressed by a variety of validated individualized psychological interventions against chronic low back pain. A prominent approach is the STarT Back Screening Tool, used to classify patients with low back pain into tailored focus therapy groups (exercises and cognitive-behavioural strategies) based on bio-psycho-social factors.^27^ Stratification, or even better personalisation/individualisation of exercise and psychological interventions may consequently lead to better therapeutic outcomes (fewer days of illness, improvement of symptomatology) than mono-interventions or, even non-personalized therapies only.^15^^32^

Consequently, individualized exercise with or without psychological interventions were investigated by a large number of clinical trials. However, individualisation as an independent variable defining exercise has not been assessed on a meta-analytical level before. There is one recently published review analysing the effects of classification-based exercise to treat chronic low back pain,^56^ showing small and no meaningful effects. However, this study focussed on classification systems and not on exercise that was individualized based on the patient’s needs. In addition, exercise was often combined with psychological interventions, which may have impacted the overall outcomes. Consequently, the aim of this systematic review with meta-analysis and –regression is to qualify and quantify the effects of individualized exercises as mono-therapies, or in combination with cognitive or behavioural types of psychological interventions. Adding many sensitivity analysis allows to distinguish active and passive controls, and to differentiate the type of individualized exercise. This consolidates previous knowledge and builds a bridge to known effects such as the above mentioned classification systems ^56^ or classical exercise.^22^

## Methods

The systematic review with meta-analysis and meta-regression protocol was developed using guidance from the Preferred Reporting Items for Systematic Review and Meta-Analysis Protocols (PRISMA-P) statement,^36^ registered in the PROSPERO database (CRD42021247331, Date of registration in PROSPERO: 10 May 2021, Date of first submission: 09 April 2021, update 10 February 2022, available at: https://www.crd.york.ac.uk/prospero/display_record.php?RecordID=247331).

Studies were identified by searching multiple databases, including PubMed, Cochrane Central Register of Controlled Trials, EMBASE, Clarivate Web of Science, and Google Scholar from their inception to March 2021. The search terms were identified after preliminary searches of the literature and by comparing them against previous systematic reviews. A sensitive search strategy was used based on the recommendations of the Cochrane Back and Neck Group for ‘randomized controlled trials’ (RCT) and ‘low back pain’,^14^ combined with search terms for ‘exercise’. The full search strategy is presented in the supplementary data appendix B. In addition, reference lists of relevant reviews and included RCTs were manually searched, and citation tracking of all included trials was performed. A grey literature search in google scholar was further performed. The searches and inclusion criteria were restricted to English, German, French, Spanish, and Italian.

We tested the eligibility criteria by piloting a small sample (10 trials). Two independent researchers (JF, PF) screened titles and abstracts of the publications retrieved by the search strategy and assessed the full texts for potential inclusion. Studies not meeting the inclusion criteria were discarded. Disagreements between researchers were resolved by discussion and consultation with a third reviewer (DN), if necessary.

For the sake of brevity, the term (chronic) low back pain in this manuscript refers to chronic non-specific low back pain if not stated otherwise.

### Eligibility criteria

This study followed the participants, interventions, comparisons, outcomes, and study design (PICOS) framework.

### Population

The population of interest was patients suffering from chronic non-specific low back pain that have been treated with any approach of stratified, personalized and/or individualized exercise. Chronic non-specific low back pain is defined as pain without a known pathoanatomical cause ^34^, chronicity of pain lasting longer than 3 month, or recurrent low back pain defined as two episodes per year.^22^ All studies had to research populations were the aforementioned criteria were fulfilled for the majority of their included patients to be considered potentially eligible.

### Intervention (exposure)

The interventions/exposures were all forms of individualized, tailored, personalized, or stratified exercise of any frequency, intensity, volume or type, with a duration of at least 4 weeks. Interventions that consisted of exercise alone or exercise combined with psychological interventions (i.e. behavioural or cognitive, or combined behavioural-cognitive therapy) were considered.

### Comparison

Eligible comparators were all matched or non-matched conservative (non-surgery) control/comparator groups, that could be either active, passive, or no treatment (real) controls

### Outcomes

We reported the following outcomes up to 1 year after randomisation:

Pain intensity, measured with a pain scale (for example, visual analogue scale (VAS), numerical rating scale (NRS), or McGill pain score)

Disability, measured with a back pain-specific scale (for example, the Roland-Morris Disability Questionnaire (RMDQ)^49^, or the Oswestry Disability Index (ODI)^12^). The two scales can be matched.^5^

All scores were normalized to VAS-scores (range 0 to 10, with 0 indicating no burden and 10 = maximum suffering from symptom).^54^

Planned secondary outcomes were the number of days of absence from work and a measure of quality of life.

Study design: Only RCTs were eligible.

Data extraction: We designed and piloted a data collection form created with Excel (Microsoft). Two researchers (JF, PF) independently extracted the study characteristics and outcome data. Disagreements were resolved through discussion or with assistance from a third researcher (DN), if necessary. From each study we extracted: number of study arms, predetermined duration of chronic non-specific low back pain, real duration of low back pain, number of participants (total and in study arms), age, sex, type of interventions (individualized, exercise, psychological), type of control, length of training period (both intervention and controls, training frequency (sessions per week), single training duration (minutes), number of adverse events.

For the effect size calculations, change scores of the outcomes and standard deviations of the change scores of the outcomes were retrieved. If not directly retrievable from the studies, the required data was calculated/imputed from the available data (baseline, change score, follow-up-values, and their standard deviations) according to the formulas defined in the Cochrane handbook. In addition (or, were not applicable), missing data was requested from the corresponding authors.

Effect sizes were processed as negative values (indicating a decrease in pain, which equals a symptom improvement), i.e. higher negative values indicate larger effect sizes.

Study risk of bias assessment: Two researchers independently assessed the risk of bias (RoB) for the outcomes of interest of the included trials. We assessed the RoB for each study/outcomes using the following RoB assessment tools recommended by the Cochrane Collaboration:^26^ random sequence generation, allocation concealment, blinding of participants, providers and outcome assessment, incomplete outcome data (dropouts) and selective outcome reporting. In the selective outcome data, we accounted for a broader assessment considering also the selective non-reporting RoB due to missing results in index meta-analyses (e.g., missing or unavailable outcome results crosschecked from method plans) according to published criteria.^43, 44^ For each study, the score of the items indicated high, low or some concerns (not enough information reported) regarding RoB.^26^ As both pain and disability are considered comparable in terms of the impact of bias on the effect sizes, they were rated cumulatively.

### Certainty assessment

Certainty of evidence was assessed applying the framework as provided by the Grading of Recommendations Assessment, Development, and Evaluation (GRADE) system of rating quality of evidence and grading strength of recommendations in systematic reviews (see supplementary data appendix F.^17^ The five GRADE domains were applied: study limitations, indirectness, inconsistency, imprecision and publication bias.

To interpret the clinical relevance of the meta-analyses results, we considered a 1.5-point difference in pain and a 1.0-point difference on a 10 point VAS in disability to be clinically important for the comparisons of exercise versus any comparator group .^22^ We interpreted smaller differences in the effectiveness of exercise treatments compared to other conservative treatments as ’probably meaningful’, when the 95% confidence interval was entirely on one side of the no effect line. This is relevant given similar inconveniences and adverse effects for comparison of treatments considered in this review.^47^

### Statistics:Meta-analysis and -regression

For data pooling, random-effects meta-analyses were modelled. Main analyses were calculated separated by outcome (pain versus disability), by the combined comparator (active/exercise or passive comparator group), and by the intervention duration (short-and long-term-effects). Weighted standardized mean differences between the intervention and control/comparator groups (Hedge’s g) were used as effect size estimators. For pooling analyses, mean effect sizes and their 95% confidence intervals were calculated; summary estimate data were displayed using Forest plots. To test for overall effects, Z-statistics at a 5% alpha-error-probability level were calculated.

Between-effects heterogeneity was assessed using the I^2^- and, using restricted maximum-likelihood estimators, Tau^2^-statistic. In case of heterogeneity (tau^2^ > 0) a prediction interval for the true outcomes was also provided.

Sensitivity analyses were performed including only effect sizes from studies without a serious overall RoB and using the exercise type, the type of psychological intervention, and the characteristics of the comparator group.

To detect outliers on study level, Studentized residuals and Cook’s distances were used. Outliers were Bonferroni-corrected with two-sided alpha = 0.05.

To check the included studies for a potential publication bias, Funnel plots and Egger’s regression test, to check a potential funnel plot asymmetry using the standard error of the observed outcomes as predictor, were used.

All pooled effects analyses were performed using the MAJOR package for jamovi (version 1.6.23, jamovi.org, Sydney, Australia).

Meta-regressions were further calculated. Independent variables were the intervention duration, the type of comparator (active versus passive), known effective versus rather ineffective exercise types, control group matched to the intervention group “yes versus no”, behavioural treatment “yes versus no”, number of the participants, overall RoB rating, and mean age of the participants. The dependent variables were the effect sizes for pain and disability (continuous). For the regression model, a syntax for IBM SPSS was used (D.B. Wilson; Meta-Analysis Modified Weighted Multiple Regression; MATRIX procedure Version 2005.05.23). Inverse variance-weighted regression models with random intercepts and fixed slopes were calculated. Homogeneity analysis (Q and p-values), meta-regression estimates (95% confidence intervals and p-values), and Z-statistics were calculated.

## Results

Our electronic database searches identified 1,172 unique citations. We screened the full-texts of 109 publications, identified 73 as potentially eligible, and included 58 RCTs of individualized exercise treatment for chronic low back pain in this systematic review with meta-analysis and meta-regression. At least, 53 studies contained sufficient data to be included into the qualitative analysis, with five studies lacking sufficient follow-up data. Figure 1 provides detail about the flow of citations and studies through our search and selection process.References to included and excluded studies at the full-text stage are provided within supplementary data appendix D.

**Figure 1:**
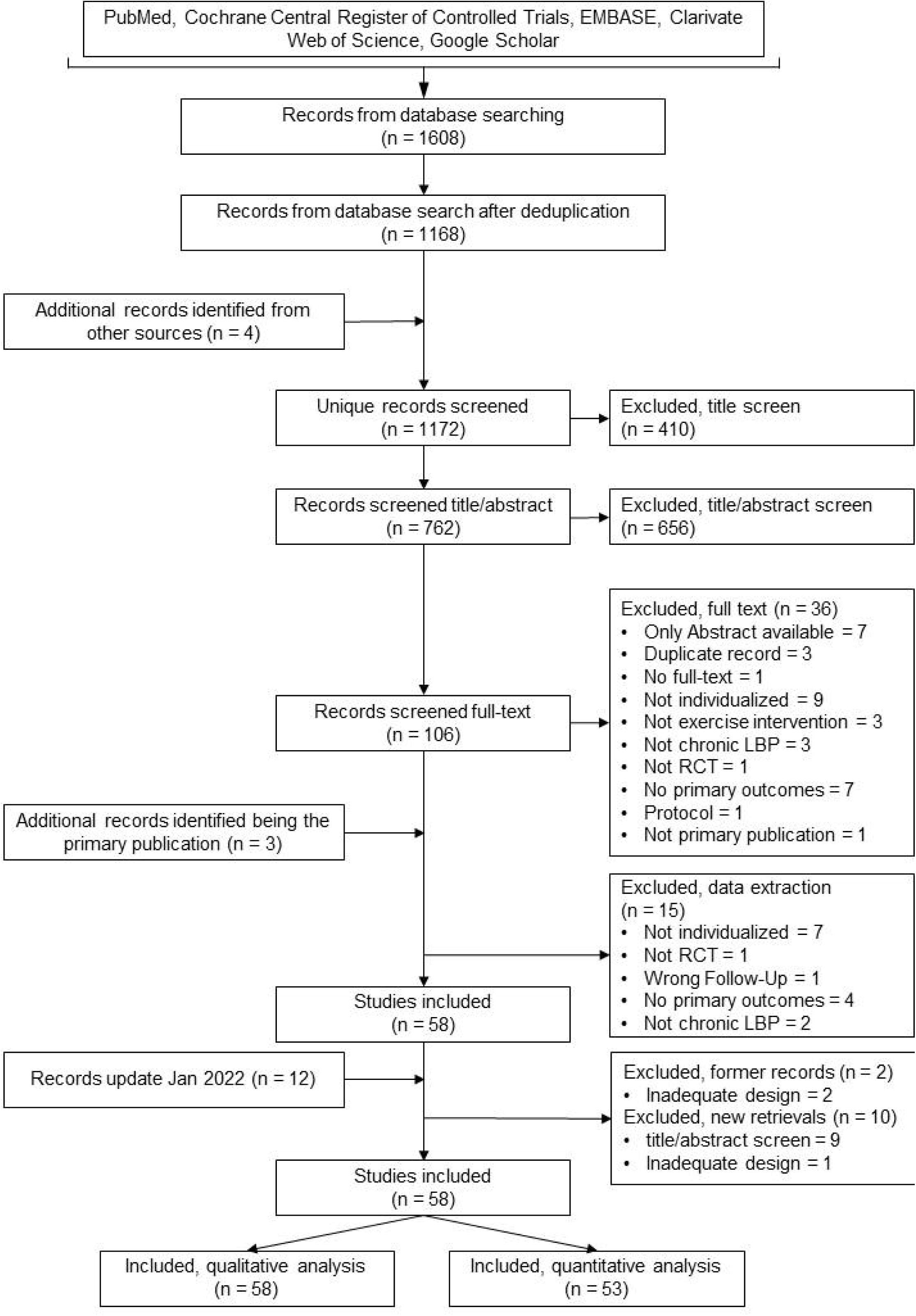
Flow Chart. RCT = randomized controlled trial, LBP = chronic non-specific low back pain

Study characteristics: A total of 10084 participants were included in 58 trials (Table 1, supplementary data appendix D). The sample size of trials ranged between 69 and 238 participants (IQR) with a median of 121 participants for each study. Reported inclusion criteria for low back pain in trials was more than 12 (6 to 12) weeks prior to study inclusion, and if reported the median time of perceived burden was 111 (55-450) weeks. Thirteen studies included as well sub-chronic pain duration and sub-acute intervals of low back pain,^2, 4, 8, 13, 16, 20, 24, 29, 31, 46, 48, 52, 60^ but consisted with a major share of participants with chronic or chronic-recurrent pain.

**Table 1.**
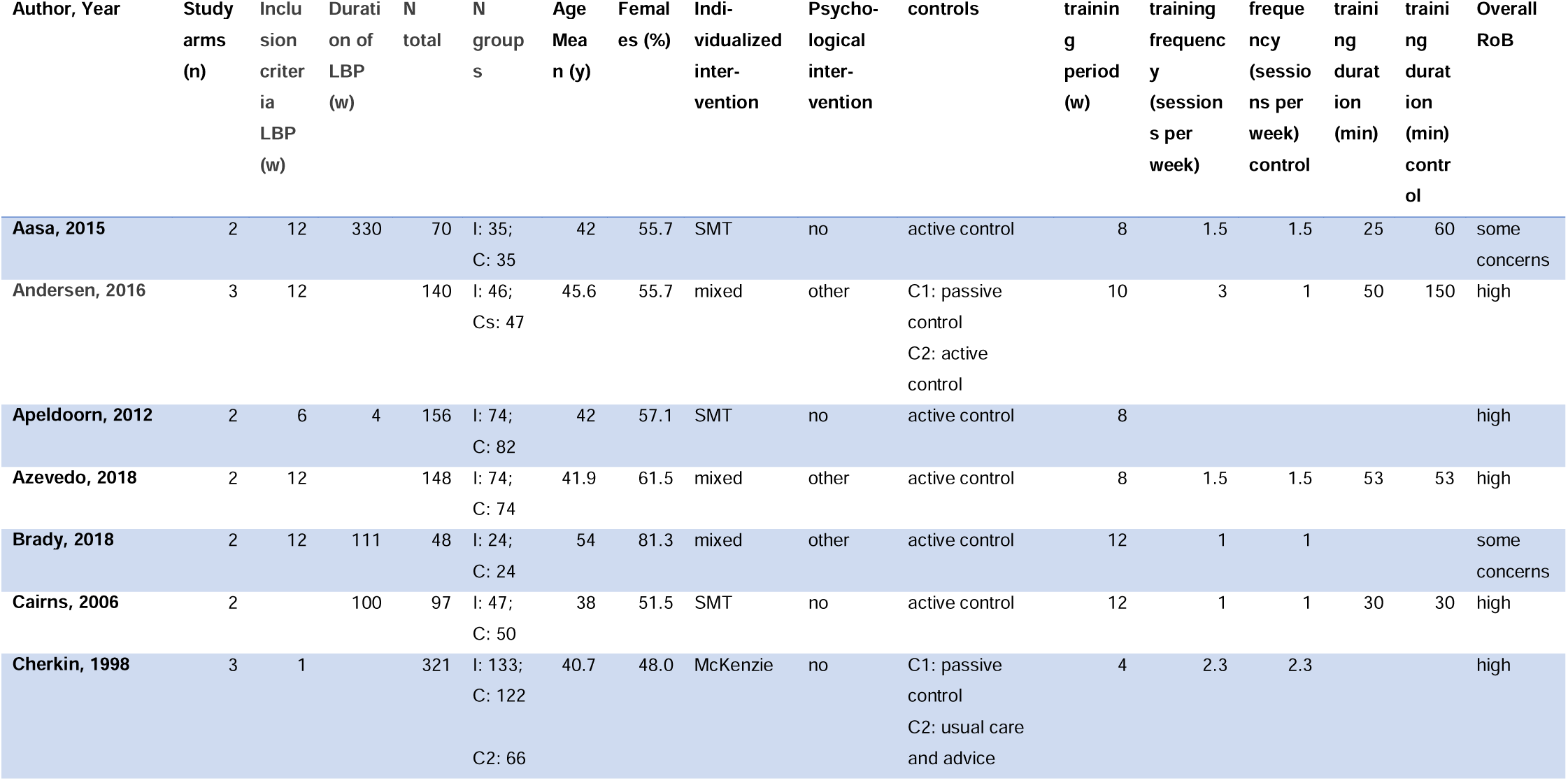
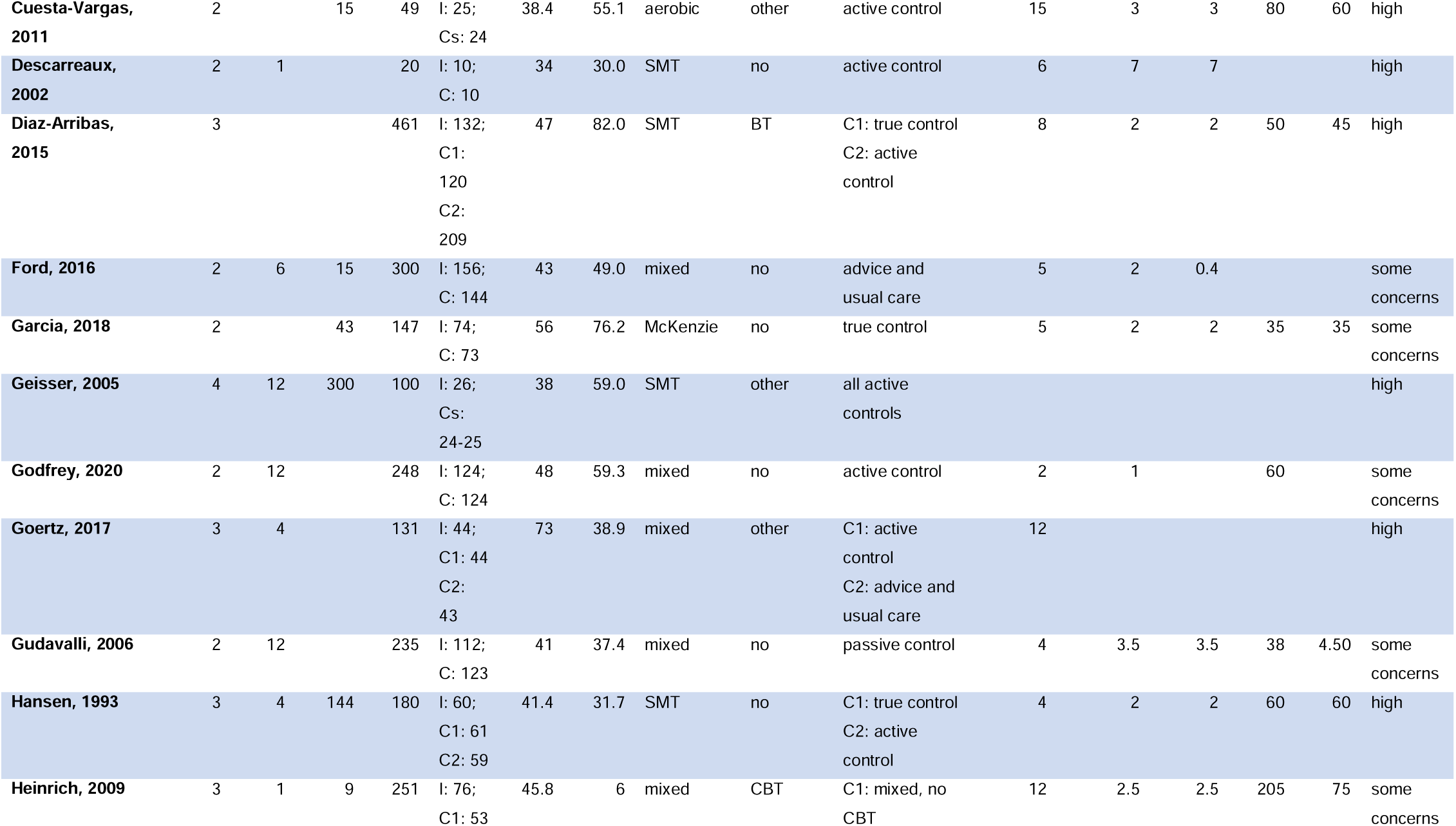
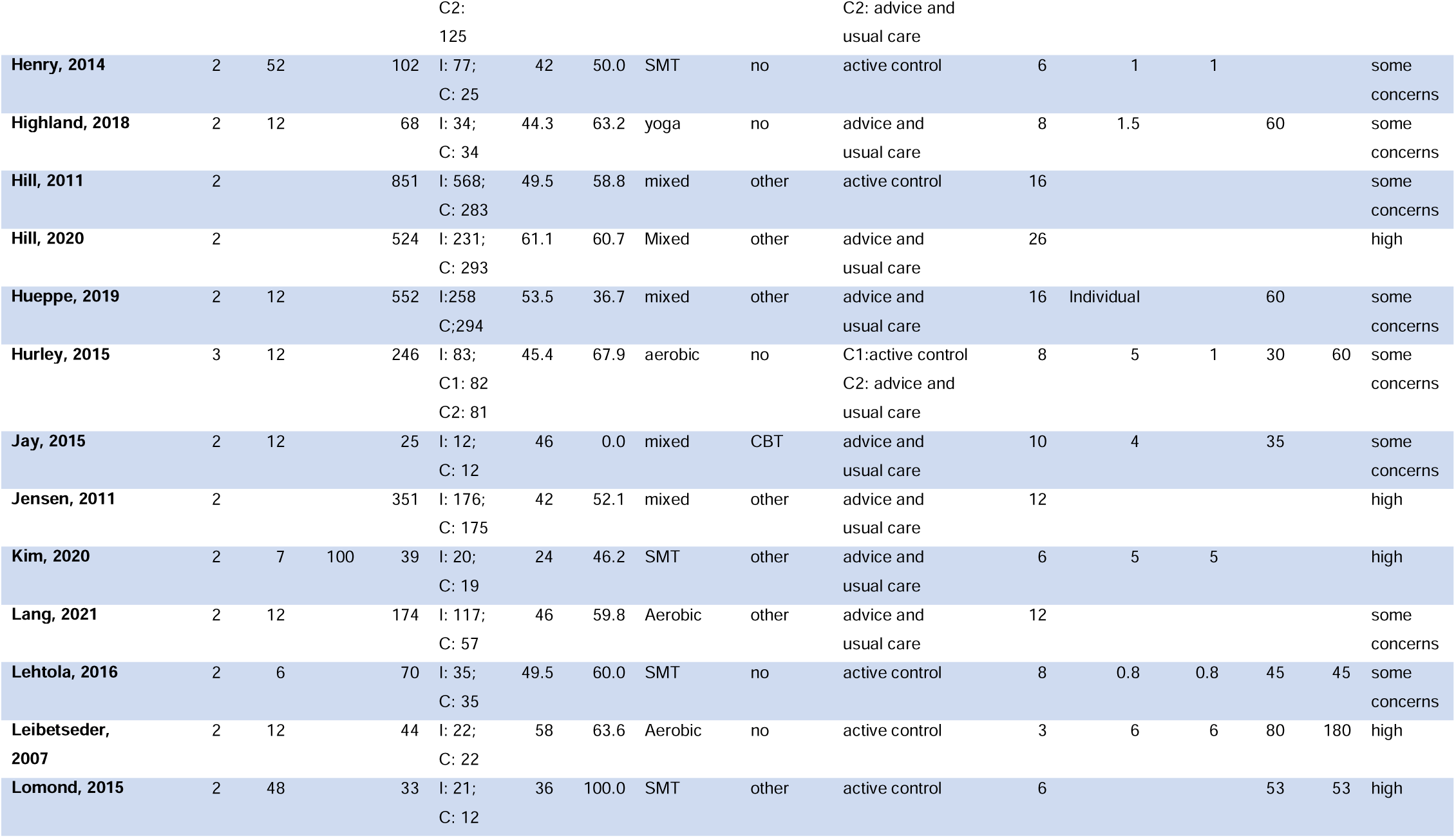
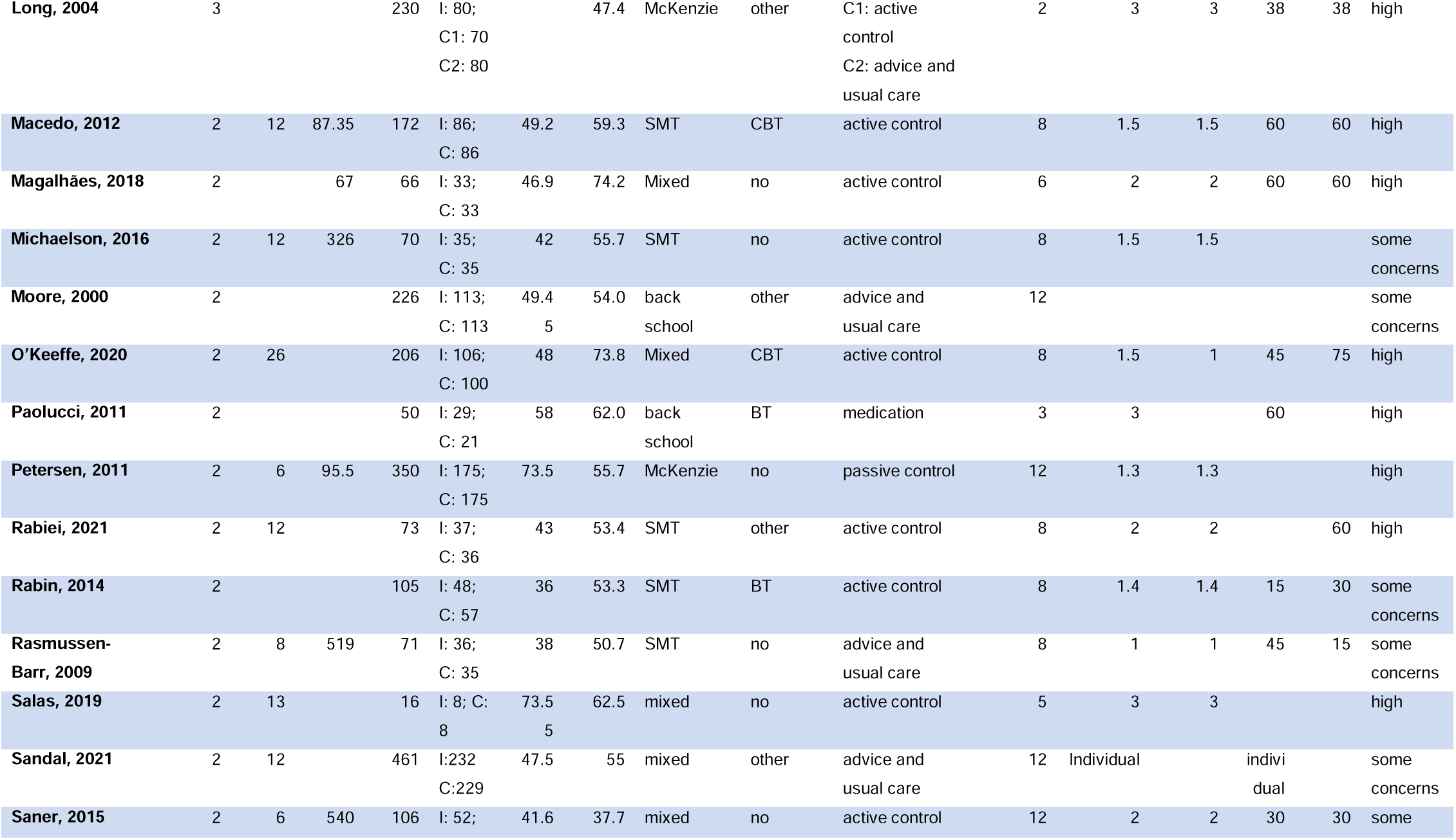
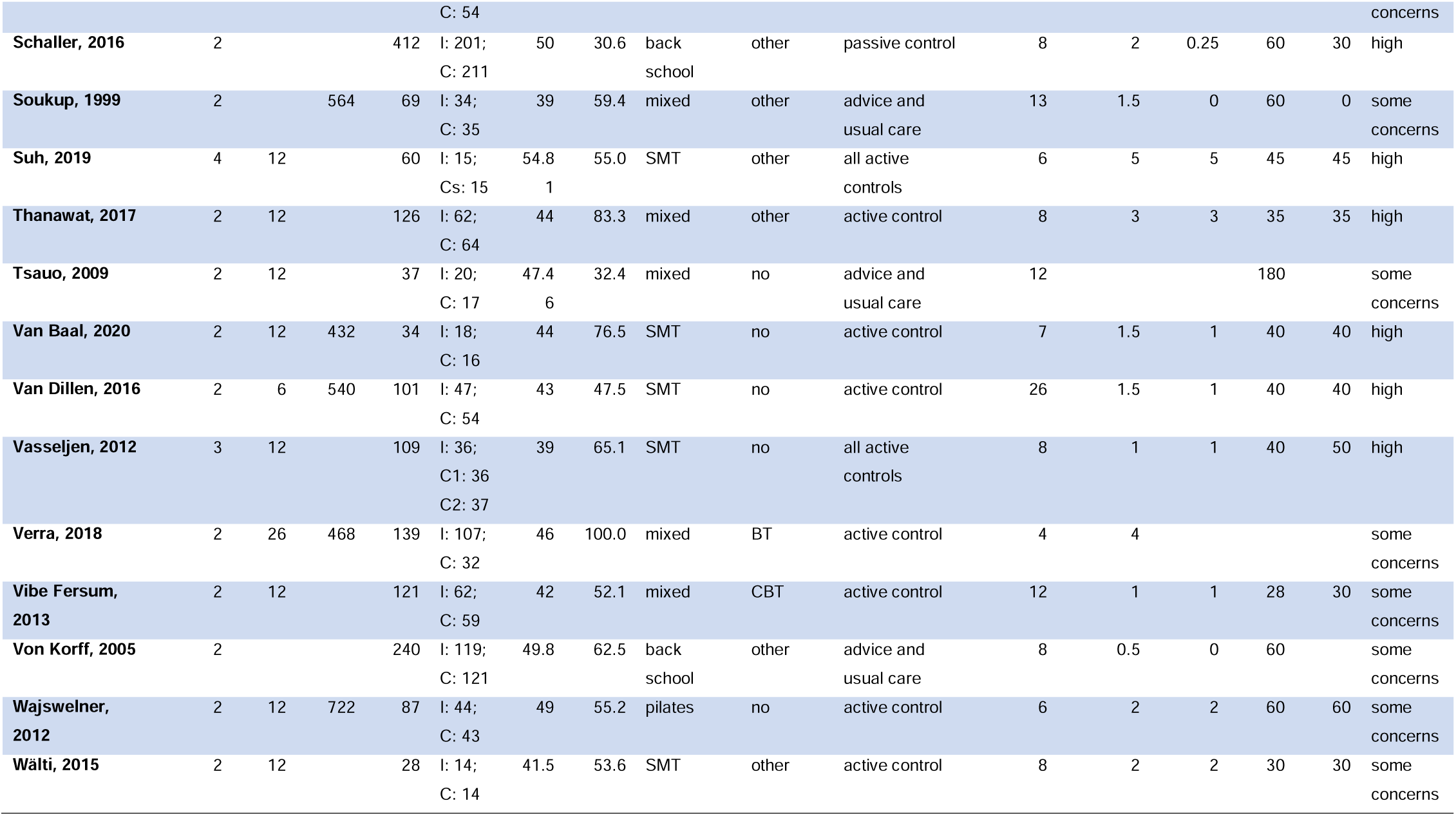
Characteristics of included studies. Inclusion criteria of LBP indicates if the study had a defined onset of the current LBP episode, whereas duration of LBP refers to the overall duration of the complaint if indicated. I = individualized group, C(s) = control group(s), SMT = sensorimotor training, BT = behavioural therapy, CBT = cognitive behavioural therapy, LBP = chronic non-specific low back pain, w = weeks, y = years, n = number, min = minute, RoB = Risk of Bias

The median year of publication of RCTs was 2015 (range 1993–2021). The median age of participants was 45.4 years (IQR 41.7-49.4), and the median percentage of females was 55.7% (50.2%–62.5%). The median training period was 8.0 (6.0-12.0) weeks, and the median number of training sessions per week was 2.0 (1.5-3.0) for the individualized exercise and 1.5 (1.0-2.4) for the control exercises. Duration of training was 50.0 (35.0-60.0) minutes for individualized, and 50.0 (30.0-60.0) minutes for control exercise. Twenty-four (41.4%) studies investigated mixed individualized exercises, and 22 studies (37.9%) sensorimotor training (SMT). Other individualized interventions included aerobic (n = 5), pilates (n = 1), McKenzie (n = 4), back schools (n = 4), and yoga (n = 1).

Thirty-four (58.6%) studies added some form of psychological intervention to the exercise procedure, i.e., cognitive-behavioural therapy (n = 9, 15.5%), behavioural therapy (n = 2, 3.4%), or other (n = 24, 34.8%).

In total, 61 control groups could be identified, with eleven studies presenting more than one control group. Forty-two (68.9%) of control interventions were non-individualized active exercises, and 21 (34.4%) no treatments with advice to stay active and usual care. Other controls were true controls (no treatment, n = 3), passive controls (n = 5), and medications (n = 1). Eleven of these control groups were matched (i.e., same type of exercise except individualisation).

There was not sufficient data to analyse secondary data.

### RoB assessement

RoB assessments on study level (as derived from the outcomes) are summarized in Figure 2 and supplementary data appendix E. Of the 58 studies, 26 (44.3%) had an overall unclear RoB (i.e., some concerns), and 32 (55.7%) a high overall RoB. In summary, there was a considerably high overall RoB and this due to deviations from intended interventions (Figure 2, supplementary data appendix C).

**Figure 2.**
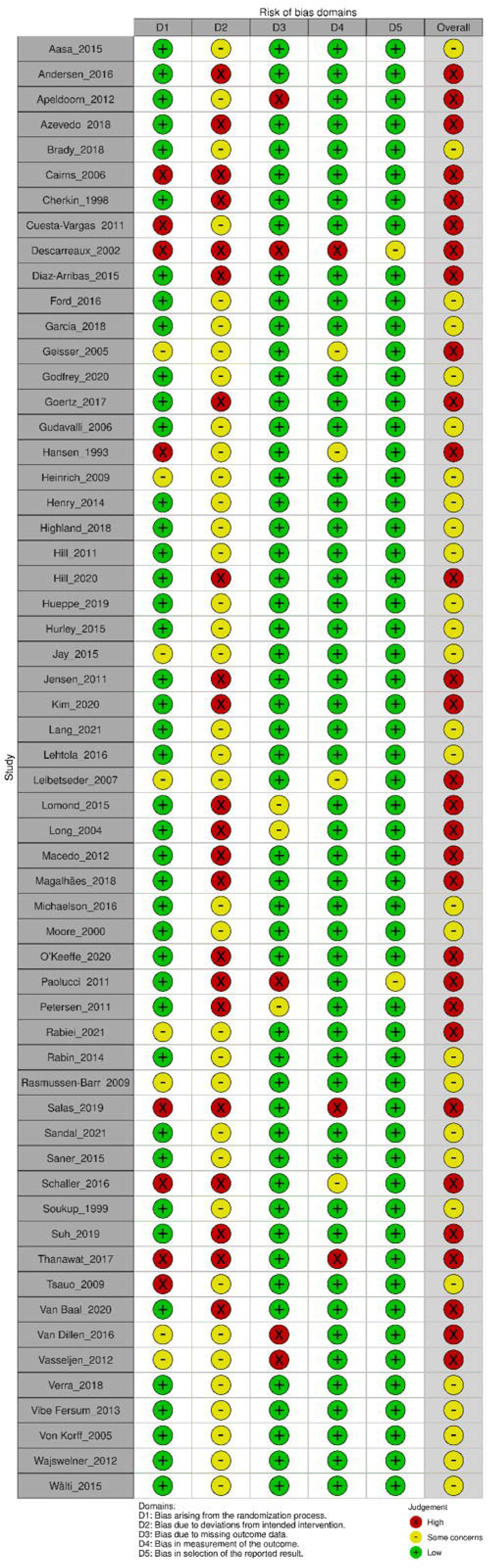
Risk of bias rating on an individual study level, displayed as traffic light plot for each study/outcome with green lights = low, yellow = unclear/moderate, and red = high risk of bias. The aggregate Cochrane Risk-of-bias appraisal results summary plot is displayed in the supplementary data appendix E.

The funnel plot (supplementary data appendix E) highlights the RoB across studies (publication bias on meta-level for the main outcomes pain and disability (short-term, in comparison to any comparator group). For the detailed main analyses, both the rank correlation and the regression test indicated no funnel plot asymmetry for pain in the short-term follow-up versus active treatments (Egger’s regression test = -1.407, p = 0.2; rank correlation: -0.202, p = .1) but versus passive treatments (test = -3.313, p = 0.001; rank = - 0.373, p = 0.009). In the long-term follow-ups on pain, no publication bias was indicated (rank = 0.117, Egger = 0.18, both p > .05; versus active; rank = 0.029, Egger = 0.288, both p > .05 versus passive). No funnel plot asymmetry was detected in any disability model (rank between -0.029 and -0.173, Eggers between -0.623 and -0.030, all p> .05). Only for the pain model of short-term follow-up versus passive measures, a publications bias is supposable, whereas it is unlikely for the pain contrast to active measures or long-term pain and disability models.

### Overall GRADE assessment

The overall certainty of the evidence was rated as low for meta-analytic outcomes of self-reported pain and disability. The main reasons for downgrading the evidence were RoB, inconsistency, and publication bias. Imprecision and indirectness were no problem, as this systematic review encompassed specific populations, types of interventions, and outcome measures (see Table 2 and the certainty of evidence section in supplementary data appendix F,).

**Table 2.**
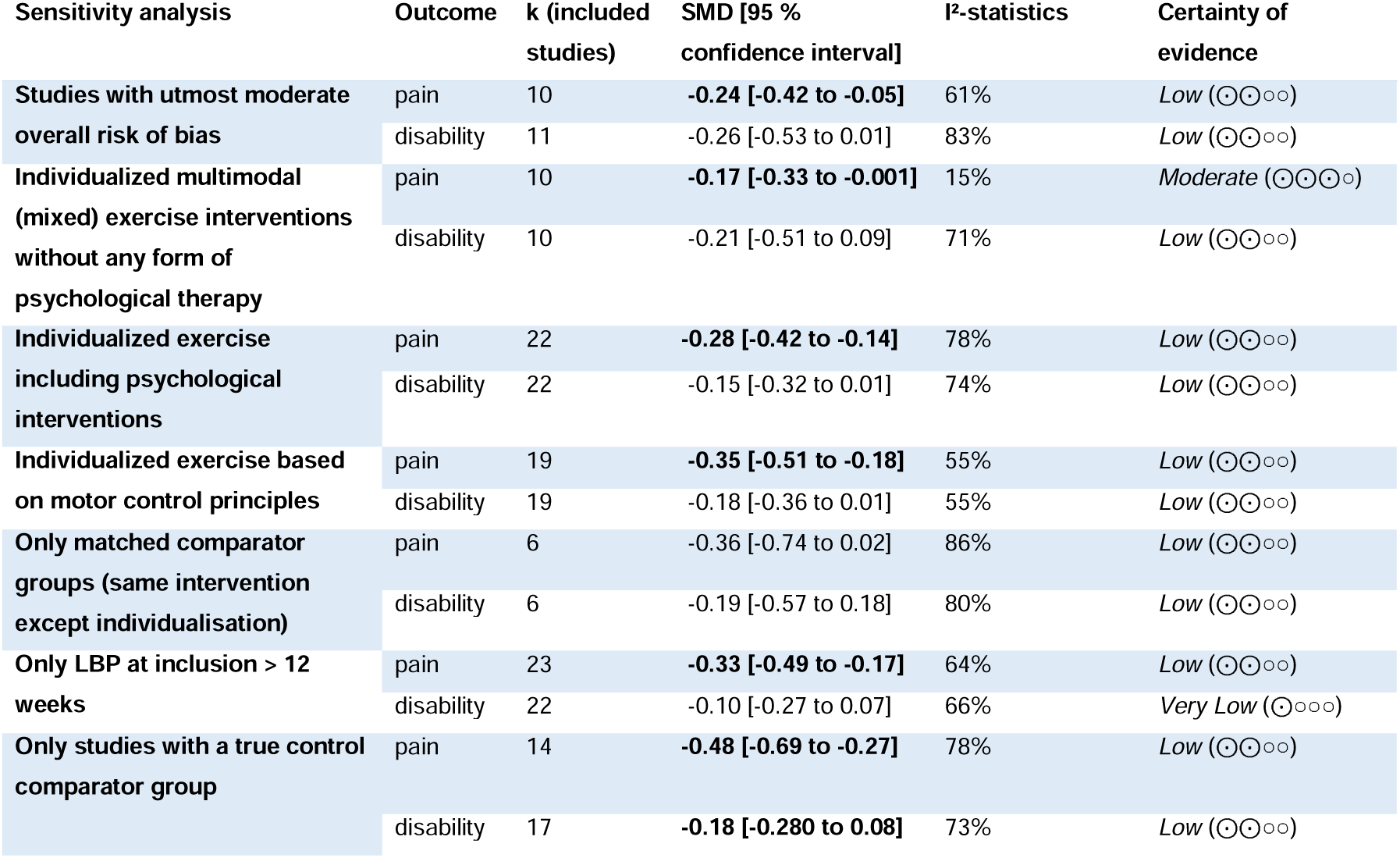
Sensitivity analysis of outcomes pain and disability at short-term follow-up comparing individualized exercise with active controls. Bold standardized mean differences (SMD) indicate significant effects towards the individualized group. Certainty of evidence was rated according to the GRADE system.^18^ SMT = sensorimotor training

### Results of individual studies

Pain intensity was assessed in 46 studies at short-term (closest to 12 weeks after randomisation or after 12 weeks of intervention) or at long-term follow-up (closest to 12 months after randomisation).

Mean pain intensity at baseline was 4.90 (standard deviation 1.38) on a VAS in the individualized groups and 4.89 (1.45) in controls. Pain following individualized exercise was reduced to 2.88 (1.25) at 12 weeks and 3.15 (1.09) at 52 weeks. Pain intensity in controls decreased to 3.62 (1.24) at 12 weeks and 3.49 (1.14) at 52 weeks.

Disability was assessed in 45 studies at short-term or at long-term follow-up. Mean disability at baseline was 3.53 (1.42) VAS in the individualized groups and 3.46 (1.51) in controls. Disability following individualized exercise was reduced to 2.41 (1.48) at 12 weeks and 2.47 (1.39) at 52 weeks.

At 12 and 52 weeks, disability in controls was reduced to 2.65 (1.51) and 2.70 (1.21),. For individual differences in studies see supplementary data appendix C.

### Pain intensity

Individualized exercise significantly reduced pain compared with all active comparators at short-term follow-up (closest to 12 weeks after randomisation; I^2^ = 73%, k = 32 SMD −0.28; 95% CI −0.42 to – 0.14; Figure 3), but not at long-term follow-up (closest to 12 months; I^2^ = 41%, k = 16 SMD −0.10; 95% CI −0.21 to 0.01; supplementary data appendix H). Mean clinically important differences did not suggest relevance at short-term (−0.67 cm VAS) or long-term follow-up (−0.19).

**Figure 3.**
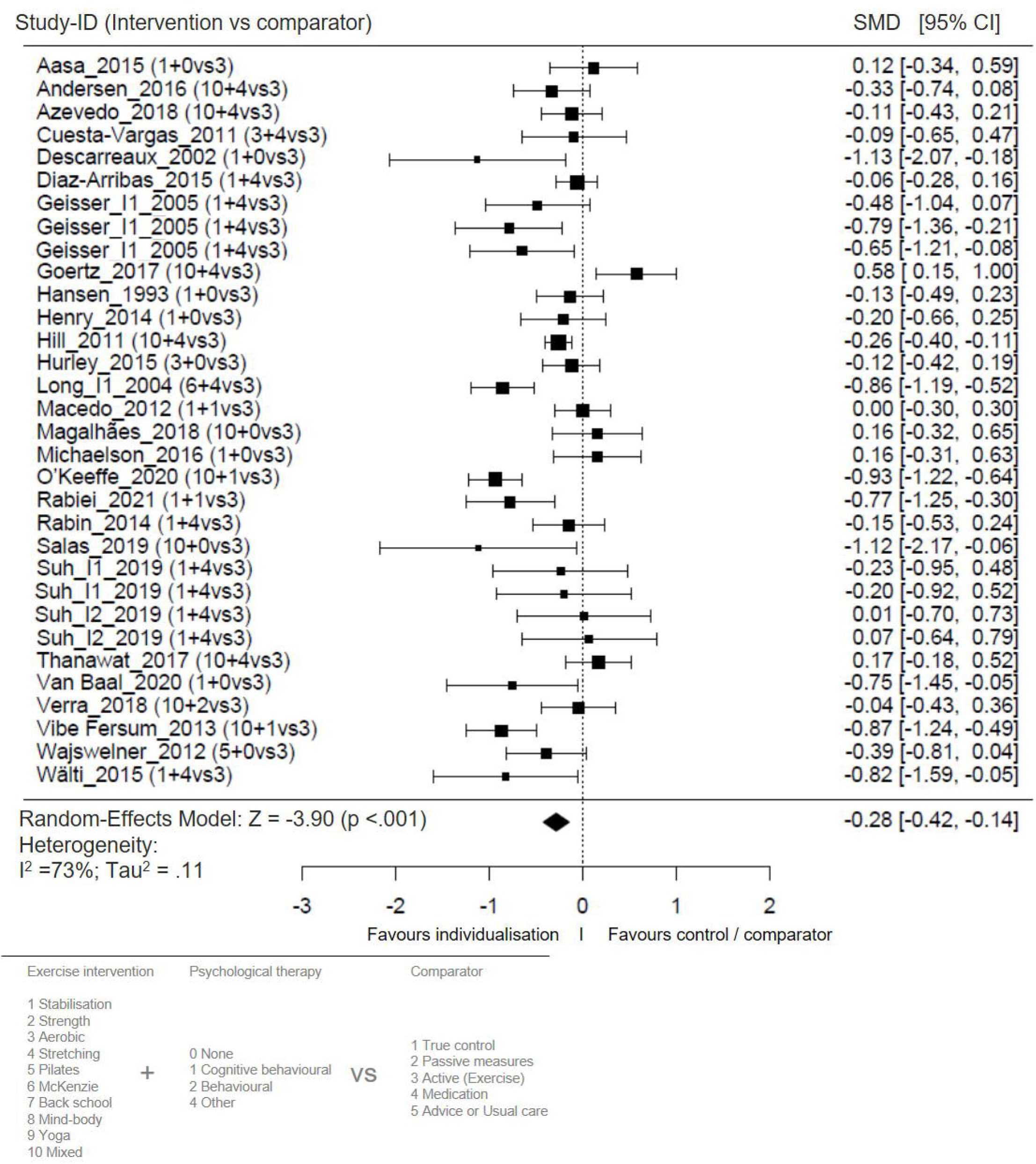
Forest plot for the effect sizes for the short-term follow-up (closest to 12 weeks following randomisation) of individualized exercise versus other active treatments on pain intensity. The plot depicts model fit, individual study and pooled effect size estimates (standardized mean differences (SMD) and corresponding 95% confidence intervals (95% CI)). The size of the boxes corresponds to the respective studies’ (inverse variance) weighting.

The intensity of pain following the intervention showed that individualized exercise significantly reduced pain compared with passive treatments or true control at short-term follow-up (I^2^ = 89%, k = 32 SMD −0.40; 95% CI −0.58 to –0.22; Figure 4), and at long-term follow-up (I^2^ = 29%, k = 17 SMD −0.14; 95% CI −0.22 to –0.07; supplementary data appendix H). Mean clinically important differences did not suggest relevance at short-term (−0.33 cm VAS) or long-term follow-up (−0.18).

**Figure 4.**
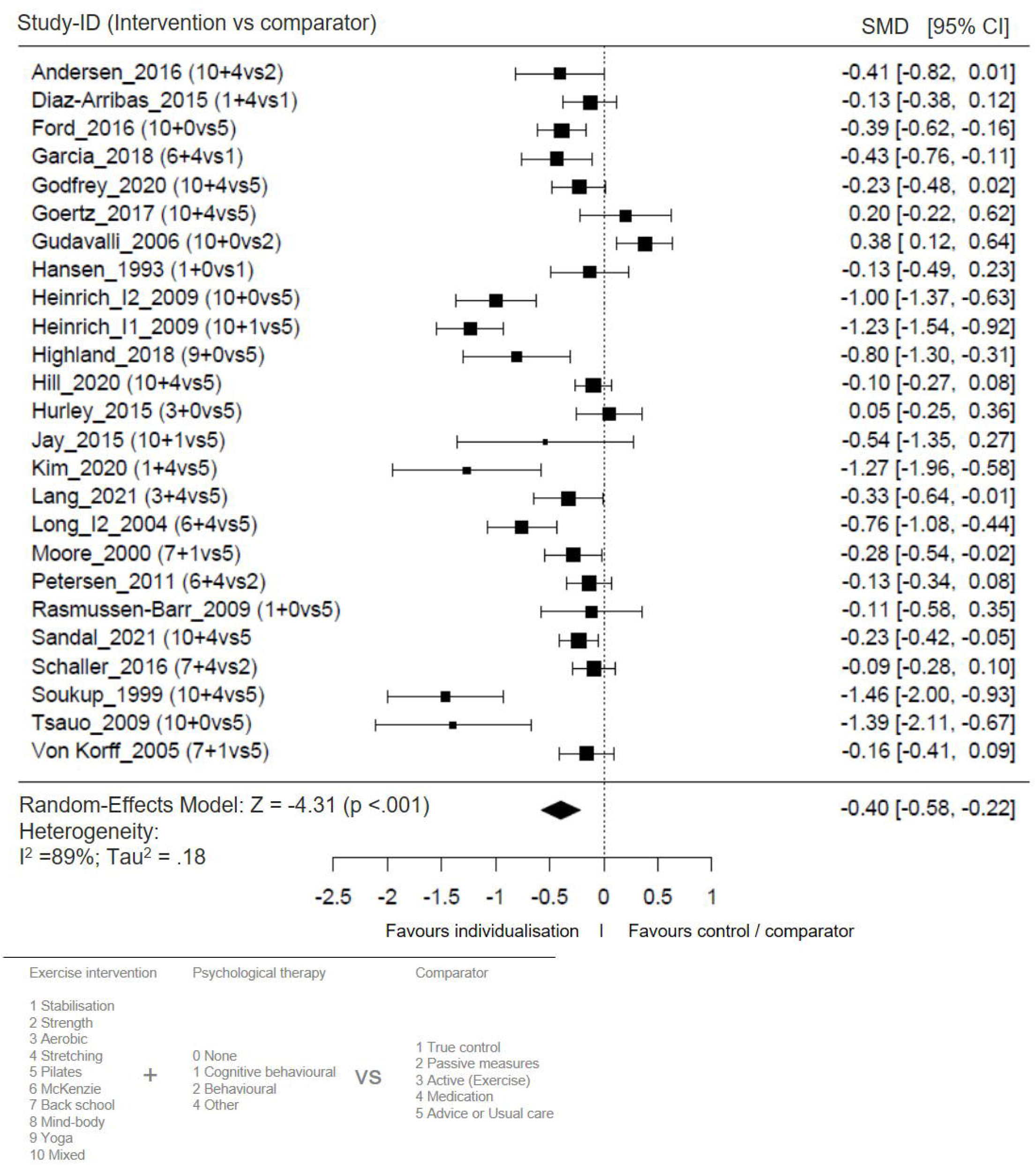
Forest plot for the effect sizes for the short-term follow-up (closest to 12 weeks following randomisation) of individualized exercise versus passive treatments, true control, and usual care on pain intensity. The plot depicts model fit, individual study and pooled effect size estimates (standardized mean differences (SMD) and corresponding 95% confidence intervals (95% CI). The size of the boxes corresponds to the respective studies’ (inverse variance) weighting.

### Disability

Under consistency, individualized exercise did significantly reduce disability compared with all active comparators at short-term follow-up (I^2^ = 73%, k = 32 SMD −0.17; 95% CI −0.31 to –0.02, Figure 5), but not at long-term follow-up (I^2^ = 69%, k = 15 SMD −0.12; 95% CI −0.27 to 0.03, supplementary data appendix H). Mean clinically important differences did not suggest relevance at short-term (−0.33 cm VAS) or long-term follow-up (−0.18).

**Figure 5.**
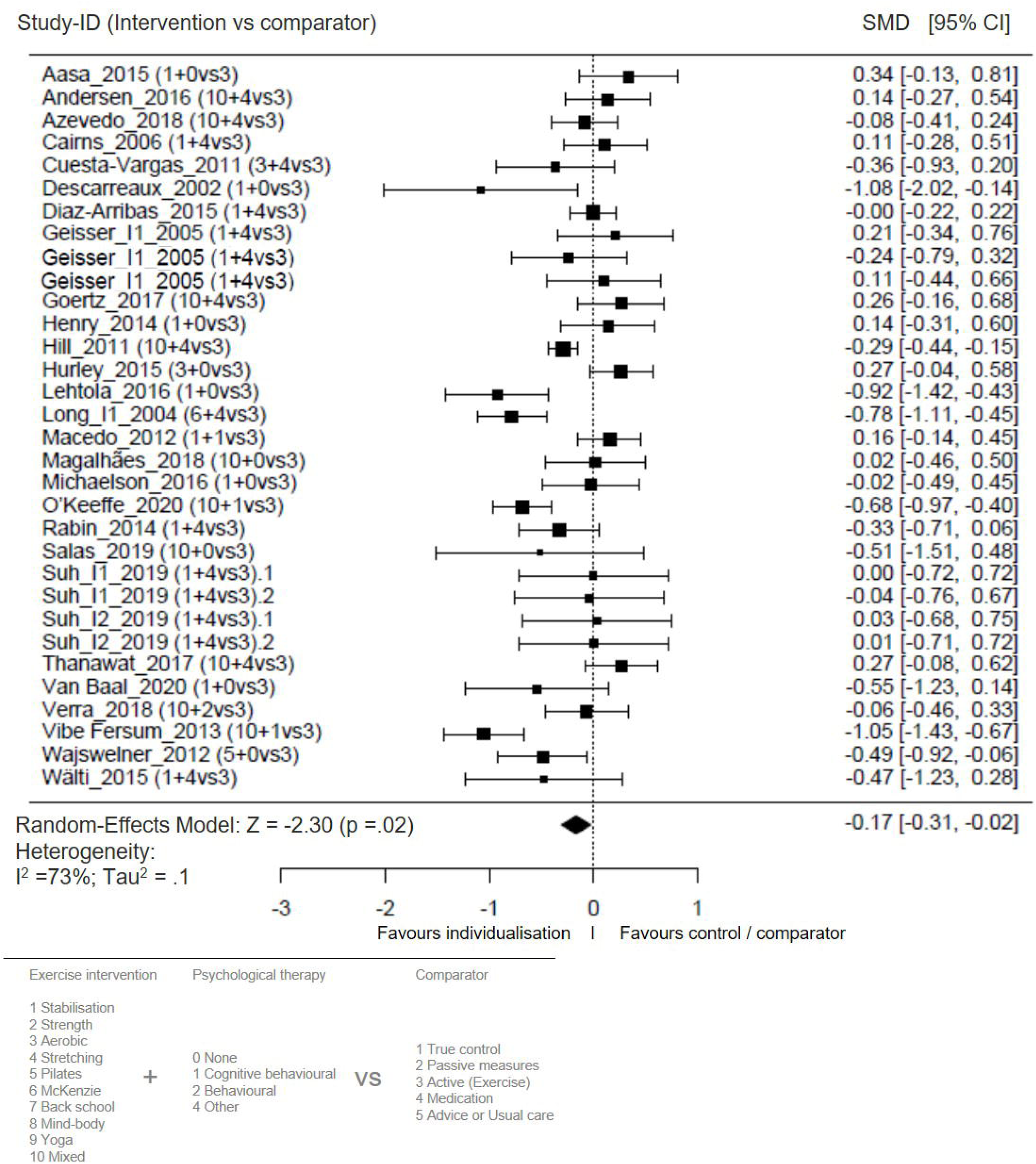
Forest plot for the effect sizes for the short-term follow-up (closest to 12 weeks following randomisation) of individualized exercise versus other active treatments on disability. The plot depicts model fit, individual study and pooled effect size estimates (standardized mean differences (SMD) and corresponding 95% confidence intervals (95% CI). The size of the boxes corresponds to the respective studies’ (inverse variance) weighting.

Individualized exercise did not reduced disability compared with passive treatments or true control at short-term follow-up I^2^ = 54%, k = 22 SMD −0.09; 95% CI −0.18 to 0.01, Figure 6) but significantly at long-term follow-up (I^2^ = 51%, k = 15 SMD −0.20; 95% CI −0.30 to –0.10, supplementary data appendix H). Mean clinically important differences did not suggest relevance at short-term (0.18 VAS) or long-term follow-up (−0.96).

**Figure 6.**
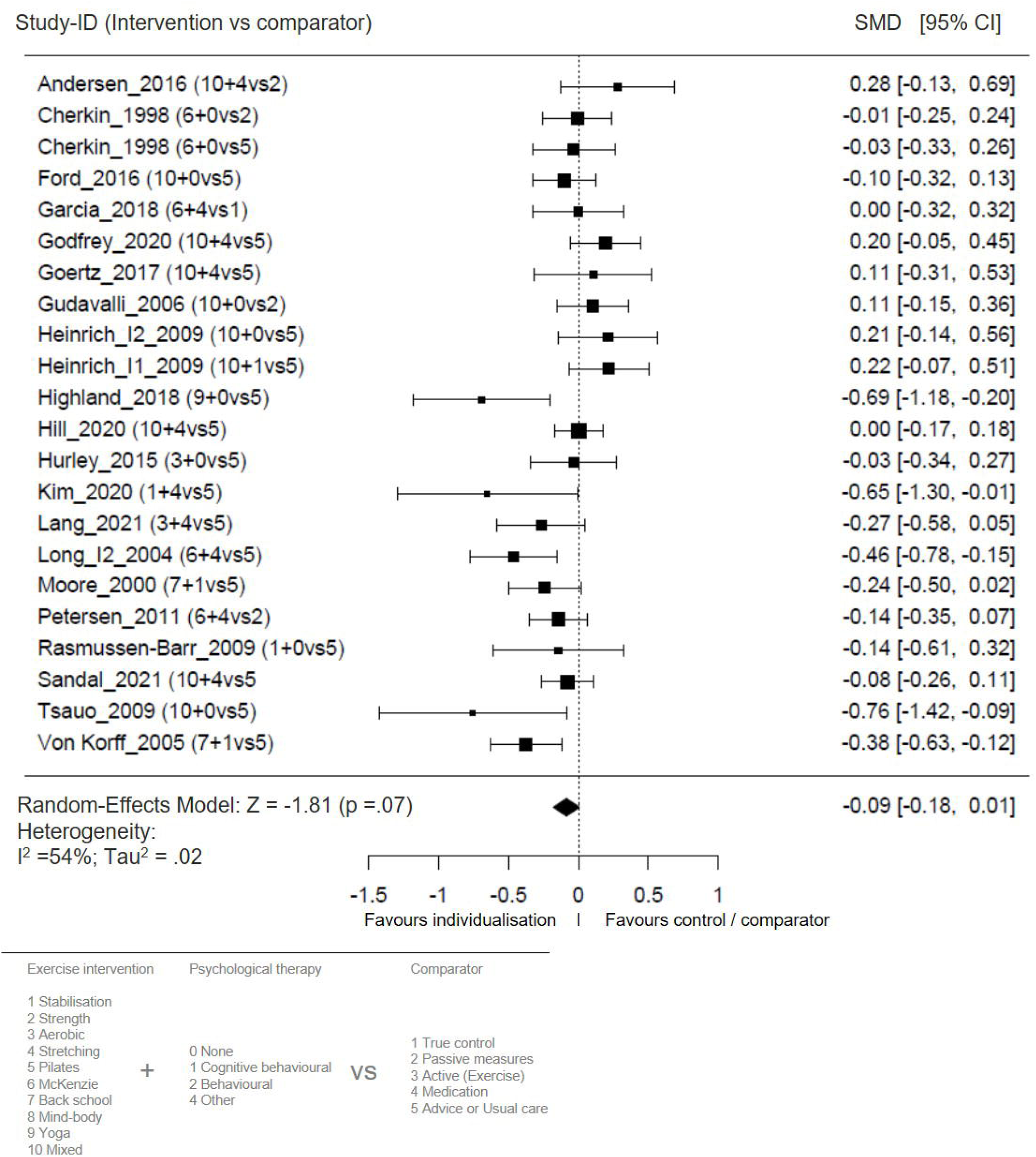
Forest plot for the effect sizes for the short-term follow-up (closest to 12 weeks following randomisation) of individualized exercise versus passive treatments or true control on disability. The plot depicts model fit, individual study and pooled effect size estimates (standardized mean differences and corresponding 95% confidence intervals). The plot depicts model fit, individual study and pooled effect size estimates (standardized mean differences (SMD) and corresponding 95% confidence intervals (95% CI). The size of the boxes corresponds to the respective studies’ (inverse variance) weighting.

### Certainty of evidence

Main outcomes on pain intensity have been graded with low (vs. active at short-and long-term) and very low at short-term and moderate at long-term (vs. passive interventions). Certainty of disability outcomes was graded as very low (vs. active) at short-and long-term) and moderate (vs. passive at short- and long-term, see *supplementary data appendix F*).

#### Sensitivity and moderator analyses

All effects at short-term follow-up were calculated in comparison to other active treatments (exercises). The effect on pain, but not on disability, was robust when only studies with utmost moderate overall RoB were pooled (according to Table 1, and *supplementary data appendix E)*

Most further sensitivity analyses, i.e., i) individualized multimodal (mixed) exercise interventions, ii) individualized exercise in combination with psychological intervention, iii) individualized exercise interventions based on SMT, iv) only studies with matched control groups, v) only studies with the inclusion criterion of low back pain lasting for at least 12 weeks, and vi) only studies with an additional behavioural treatment showed individualized exercise to significantly reduce pain intensity at short-term follow-up compared with other active treatments, but not disability (Table 2, supplementary data appendix I1-I7). In contrast, the comparisons to vi) only true control comparators revealed superior effects by the individualisation in both pain and disability.

Mean differences when compared to controls ranged from -0.09 to -1.27 VAS (depending on the sensitivity scenario) for pain intensity, with largest clinical effects for combined treatments of individualized exercise and cognitive behavioural therapy (−1.27 VAS), and from -0.09 to -0.80 for disability (see supplementary data appendix G). On an individual level, all effects of individualized exercise on pain were clinically important (> 1.50 VAS). Effects on disability were clinically important at both short-term and long-term follow-up, except for individualized exercise not including psychological interventions and studies including low back pain episodes shorter than 12 weeks.

Controls showed clinical important effects when including psychological interventions, and on long-term disability.

Dose-response relationships have been performed by means of meta-regression analysis (supplementary data appendix I8). The independent variables (intervention duration, type of comparator, distinction between rather effective or ineffective exercise, matched control group, additional psychological intervention, total study size (n), overall risk of bias rating, and mean age) explained 13% of the pain and 6% of the disability effect size heterogeneity. For pain, the only significant contributor was the training duration (in weeks, longer durations mediated larger effect sizes). For disability, no significant regressors could be identified (see supplementary data appendix I8).

### Adverse Events

Adverse events have been reported by 14 (24%) of studies, with a total of 553 events, of which 50 (9%) were at least possibly treatment-related. Retrieved data did not allow to distinguish between types of active interventions or perform further analysis.

## Discussion

This systematic review and meta-analysis with meta-regression found that individualized exercise interventions versus other active exercise interventions, or passive controls and usual care were associated with reduced self-reported pain intensity and disability at short- and long-term follow-up. Our analysis revealed medium effect sizes for the reduction of pain intensity by individualized exercise with low levels of certainty when compared to active and very low certainty compared to passive treatments at the follow-ups closest to 12 weeks. Disability was reduced with low certainty and small effects size compared to active controls. At long-term follow-up (closest to 12 months) moderate certainty suggests small effect sizes comparing individualized exercise to passive treatments. We could not detect effects on pain intensity at long-term follow-up compared to active controls (low to moderate certainty). Sensitivity analysis (Table 2) revealed on a low level of certainty that the combination of individualized exercise in combination with psychological interventions (e.g., cognitive and/or behavioural therapy) showed moderate effect sizes on pain and disability compared to both active and passive treatments. Effects obtained following individualized exercise were mostly clinically relevant. Sensitivity analysis also showed superiority of motor-controlled individualized interventions (low certainty), and of individualized exercise when compared to real controls (moderate certainty).

Meta-regression did not reveal any substantial influencers.

These data indicate that general individualized exercise can be recommended in principle to achieve improvements in in pain intensity and disability in low back pain patients

From a clinical and therapeutic point of view, we calculated MCIDs implicating individualized exercise to better reduce pain and disability compared to both, active and passive controls. However, MCID mostly failed to be relevant by definition, i.e. below 1.5 points on the VAS for pain and 1.0 point for disability outcomes. Anyhow, we consider (based on the confidence intervals) most effects on pain as probably clinically relevant when individualized exercises are compared to passive measures or other exercises. The clinical relevance may be strengthened due to the following reasons: i) adding a stratification and /or personalisation to the exercise intervention is simple and almost time and cost neutral, ii) at 12 weeks pain intensity was reduced by individualized exercise by 40% and at 26 weeks by 38%, which from a patient’s perspective is of clear benefit, iii) all results are at least consistent in the point that there is a trend (or, even, mostly a significant between-group difference leading to a probable relevant effect) towards individualized treatments that becomes larger when not comparing to active but passive controls, iv) taking into account the strong known placebo effects in low back pain^61^ emphasizes the small effects retrieved in our study. As all studies were patient-examiner blinded -what hands-on trials cannot be blamed for-this observation seems robust.

Compared to other low back pain treatments, the observed effect sizes in our study are superior to, e.g., paracetamol (which has been declared being not effective compared to placebo),^53^, somewhat larger than therapeutic ultrasound (small to no effects versus placebo),^10^ non steroidal anti inflammatory drugs (small effects versus placebo),^58^ spinal manipulative therapy (small effects versus standard care),^7^ and comparable to acupuncture (low certainty evidence to reduce pain and improve function compared to sham and no treatment).^37^ Hayden et al. recently assessed the effects of exercise on chronic low back pain and disability to be small compared to no treatment, usual care or placebo comparisons, and other conservative treatments.^22^ Certainty of evidence is low to moderate, and studies did not meet the threshold for minimal clinically important difference.^22^ The mean difference between groups as assessed by the meta-analysis on classification-based exercise in low back pain,^56^ was presented as the difference to all comparators, suggesting small effects. All above mentioned reviews are principally in line with our results, suggesting effects on very low to moderate certainty, which are at first sight not clinically important. The problem of the above made assumptions is that the clinical relevance of the effect cannot just be defined by the MCID.^6^ Medical assessment should include other factor as well, such as regression, baseline severity, or even expectations. If the MCID is calculated on the basis of a comparison to no treatment controls, it reflects well the clinical state of art.^6, 22^ In the case of the present systematic review, patients usually received at least advice, and a larger number of no-treatment controlled studies is missing (see Table 1). Calculating the mean differences compared to no treatment controls (n = 4) or usual care and advice only (n = 16; see supplementary data appendix G) resulted in clinically relevant intragroup differences for the individualized exercise group, but not controls, making our findings more likely. Consequently, the MCID is a context-specific value rather than a fixed number,^3^ and gives insight into relative but also absolute effects. The absolute symptom reduction varied between 40% (pain) and 33% (disability) from baseline, which can indeed be considered clinically relevant from a pain physician’s perspective. Especially with regard to the combined approaches (adding psychological interventions or exercise based on motor control) the reduction of pain intensity (on a low to moderate certainty level) is promising from a clinical point of view. In this context, the impact of the results as shown by Tagliaferri et al. can be regarded clinically relevant.^56^ Summarising our data, the overall low certainty, RoB, MCID, and SMDs, data support the conclusion that individualized exercise interventions versus other active exercise interventions or usual care or passive controls are associated with clinically relevant reduced self-reported pain intensity and disability.

Recent clinical practice guidelines^1, 30, 38, 47^ are basically in line with our findings. The German national guideline on chronic non-specific low back pain^1^ strongly recommends the combination of exercise treatments with education, the latter based on behavioural therapy, to treat sub-acute and chronic low back pain, and to promote physical activity.

The NICE guidelines^38^ universally recommend to consider a combined physical and psychological intervention, incorporating a cognitive behavioural approach. Physical programs refers to a group exercise program (biomechanical, aerobic, mind–body or a combination of approaches) for people with specific episodes or flare-up of low back pain with or without sciatica that should take people’s specific needs, preferences and capabilities into account when choosing the type of exercise.

The guideline of the North American Spine Association^30^ recommends cognitive behavioural therapy in combination with physical therapy, as compared with physical therapy alone, to improve pain levels in patients with low back pain over 12 months on level A. Cognitive behavioural therapy in combination with physical therapy is recommended to improve functional outcomes (disability) and return to work in low back pain patients on a level B.

We belief that our results strengthen the use of exercise therapies in multimodal settings for low back pain therapies. On the basis of our data, the guideline statements in regard of patients’ needs and differentiation among exercise modalities seem justified. Personalisation is an important option in tailoring exercise to patient’s needs. Different exercise modalities seem to have different impact on pain and disability. Furthermore, the bio-psycho-social dimension of low back pain should also be addressed using adequate psychological interventions.

Promising research deficits regarding that should be subject to future research include aspects like digitalisation^9^ or cost-effectiveness.^35^. From a therapist’s point of view, different forms of exercise therapy should be compared with each other to determine a potentially superior mode of exercise training for rehabilitation. However, given the current paucity of literature, pairwise meta-analyses may be limited in drawing such conclusions. Network meta-analysis may be more suitable for determining potentially superior modes of exercise training and have recently gained momentum in the field of sports medicine.^42^ Studies should focus on strong methodological rigor and larger sample sizes to reduce the RoB and increase the certainty in observations. To ensure a low risk of bias, placebo- or sham-controlled trials should be considered.^28^ Furthermore, the studies should follow current guidelines for intervention description (e.g., the template for intervention description and replication checklist^28^) to enable transparent evaluation and replication of intervention programs and should report factors potentially influencing the findings (e.g. comorbidities and pain management). Despite of a low adverse effects risk being attributed to exercise therapies,^41^ the reporting of adverse events in retrieved studies was poor. Our observations mainly rely on one large study,^16^ including almost 75% (n = 414) of reported adverse events, of which 98 are treatment-related. The strengths of our study include the overall assessment of individualized exercise intervention to give a concise overview of the different modalities how personalisation in exercise therapy might be performed (Supplementary File Appendix C). We also retrieved studies that included patients with sub-chronic low back pain, and sensitivity analysis suggested these patients not to distort our analysis. This group of patients could also benefit from individualized exercise. Statistical sensitivity analyses were finally conducted for all outcomes to further check the robustness and validity of the results.

### Limitations

There were no included studies with a low RoB. We also could not assess the effect of individualized exercise on the number of days of absence from work and measures of quality of life, because the included studies did not report these important variables consistently. Consequently, our outcome is restricted to pain or disability only. It should also be noted that the results only apply to land-based interventions. In regard to our certainty assessment downgrading was necessary due to supposed inconsistency. When individual study sample sizes are small, point estimates may vary substantially but, because variation may be explained by chance, *I*^2^ may be low. Conversely, when study sample size is large, a relatively small difference in point estimates can yield a large *I*^2^.^51^ This might also be a reason for the supposed publication bias regarding pain outcomes.

In addition, the terms personalized and especially stratified do not exactly mean the same, but address same aspects of medicine. We adhere to the definition, that the term ‘stratification’ more accurately “reflects the realistic effects of medicines at population level, while the term ‘personalized medicine’ reflects the possibly overambitious promise of specifically tailoring a treatment on an individual level.^11, 62^ In addition, individualisation within our studies covers a broad field of interventions, what should not automatically imply all types of intervention being equally efficient. We are aware that future studies will need to more accurately define exercise in this regard.

Finally, although exercise training is considered relatively safe in general,^41^ adverse events could not be adequately assessed given insufficient reporting.

## Conclusions

This systematic review shows on average low-certainty evidence for the effectiveness of individualized exercise on pain and disability in chronic non-specific low back pain. The effects are of clinical importance when assessing individualized exercise by its own. Given the little extra effort to individualise exercise, practitioners should be encouraged to use this approach. Effects are larger when combining exercise and cognitive-behavioural interventions for the treatment of chronic low back pain. With regard to the bio-psycho-social dimension of low back pain, it is important to address psychosocial characteristics, such as inappropriate beliefs about pain, pain catastrophizing, and/or depression, besides bodily functions.^50^

## Data Availability

All data produced in the present study are available upon reasonable request to the authors

