## Supplementary Files for "Individualized exercise in chronic non-specific low back pain: a systematic review with meta-analysis on the effects of exercise alone or in combination with psychological interventions on pain and disability"

Supplementary data

**Content**

Appendix A. PRISMA Checklist

| **Section and Topic** | **Item #** | **Checklist item** | **Location where item is reported** |
| --- | --- | --- | --- |
| **TITLE** | | |  |
| Title | 1 | Identify the report as a systematic review. | title |
| **ABSTRACT** | | |  |
| Abstract | 2 | See the PRISMA 2020 for Abstracts checklist. | p.3 in agreement with journal style |
| **INTRODUCTION** | | |  |
| Rationale | 3 | Describe the rationale for the review in the context of existing knowledge. | p.4 |
| Objectives | 4 | Provide an explicit statement of the objective(s) or question(s) the review addresses. | l.116ff and p.5-6 |
| **METHODS** | | |  |
| Eligibility criteria | 5 | Specify the inclusion and exclusion criteria for the review and how studies were grouped for the syntheses. | p.6 |
| Information sources | 6 | Specify all databases, registers, websites, organisations, reference lists and other sources searched or consulted to identify studies. Specify the date when each source was last searched or consulted. | p.5 l.137ff |
| Search strategy | 7 | Present the full search strategies for all databases, registers and websites, including any filters and limits used. | p.5 and suppl. Data appendix B |
| Selection process | 8 | Specify the methods used to decide whether a study met the inclusion criteria of the review, including how many reviewers screened each record and each report retrieved, whether they worked independently, and if applicable, details of automation tools used in the process. | p.6 |
| Data collection process | 9 | Specify the methods used to collect data from reports, including how many reviewers collected data from each report, whether they worked independently, any processes for obtaining or confirming data from study investigators, and if applicable, details of automation tools used in the process. | p.6, figure 1 |
| Data items | 10a | List and define all outcomes for which data were sought. Specify whether all results that were compatible with each outcome domain in each study were sought (e.g. for all measures, time points, analyses), and if not, the methods used to decide which results to collect. | p.6, table 1 |
|  | 10b | List and define all other variables for which data were sought (e.g. participant and intervention characteristics, funding sources). Describe any assumptions made about any missing or unclear information. | p.6 |
| Study risk of bias assessment | 11 | Specify the methods used to assess risk of bias in the included studies, including details of the tool(s) used, how many reviewers assessed each study and whether they worked independently, and if applicable, details of automation tools used in the process. | p.7 |
| Effect measures | 12 | Specify for each outcome the effect measure(s) (e.g. risk ratio, mean difference) used in the synthesis or presentation of results. | p.7-8 |
| Synthesis methods | 13a | Describe the processes used to decide which studies were eligible for each synthesis (e.g. tabulating the study intervention characteristics and comparing against the planned groups for each synthesis (item #5)). | Table 1, figures 3-7, appendix H |
|  | 13b | Describe any methods required to prepare the data for presentation or synthesis, such as handling of missing summary statistics, or data conversions. | p.6-7 statistics |
|  | 13c | Describe any methods used to tabulate or visually display results of individual studies and syntheses. | p.6-7 statistics |
|  | 13d | Describe any methods used to synthesize results and provide a rationale for the choice(s). If meta-analysis was performed, describe the model(s), method(s) to identify the presence and extent of statistical heterogeneity, and software package(s) used. | p.7-8 statistics |
|  | 13e | Describe any methods used to explore possible causes of heterogeneity among study results (e.g. subgroup analysis, meta-regression). | p.7-8 statistics |
|  | 13f | Describe any sensitivity analyses conducted to assess robustness of the synthesized results. | p.7-8 statistics |
| Reporting bias assessment | 14 | Describe any methods used to assess risk of bias due to missing results in a synthesis (arising from reporting biases). | p.6-7 |
| Certainty assessment | 15 | Describe any methods used to assess certainty (or confidence) in the body of evidence for an outcome. | p.7-8 statistics |
| **RESULTS** | | |  |
| Study selection | 16a | Describe the results of the search and selection process, from the number of records identified in the search to the number of studies included in the review, ideally using a flow diagram. | Figure 1, l. 267f |
|  | 16b | Cite studies that might appear to meet the inclusion criteria, but which were excluded, and explain why they were excluded. | Appendix D |
| Study characteristics | 17 | Cite each included study and present its characteristics. | p. 9, Table 1, Appendix C and D |
| Risk of bias in studies | 18 | Present assessments of risk of bias for each included study. | p. 10, Figure 2, Appendix E |
| Results of individual studies | 19 | For all outcomes, present, for each study: (a) summary statistics for each group (where appropriate) and (b) an effect estimate and its precision (e.g. confidence/credible interval), ideally using structured tables or plots. | p.9,-11 figure 3-6, appendix H, appendix C1 |
| Results of syntheses | 20a | For each synthesis, briefly summarise the characteristics and risk of bias among contributing studies. | p.10,  Appendix F |
|  | 20b | Present results of all statistical syntheses conducted. If meta-analysis was done, present for each the summary estimate and its precision (e.g. confidence/credible interval) and measures of statistical heterogeneity. If comparing groups, describe the direction of the effect. | Figures 3-6, Appendix H, Table 2 |
|  | 20c | Present results of all investigations of possible causes of heterogeneity among study results. | Appendix F, Appendix E |
|  | 20d | Present results of all sensitivity analyses conducted to assess the robustness of the synthesized results. | p.11, Table 2, Appendix I |
| Reporting biases | 21 | Present assessments of risk of bias due to missing results (arising from reporting biases) for each synthesis assessed. | Appendix E, F |
| Certainty of evidence | 22 | Present assessments of certainty (or confidence) in the body of evidence for each outcome assessed. | p. 11, Appendix F |
| **DISCUSSION** | | |  |
| Discussion | 23a | Provide a general interpretation of the results in the context of other evidence. | p. 13 |
|  | 23b | Discuss any limitations of the evidence included in the review. | p.13, 15 |
|  | 23c | Discuss any limitations of the review processes used. | p. 15 |
|  | 23d | Discuss implications of the results for practice, policy, and future research. | p.13-14 |
| **OTHER INFORMATION** | | |  |
| Registration and protocol | 24a | Provide registration information for the review, including register name and registration number, or state that the review was not registered. | p. 5  PROSPERO CRD42021247331 |
|  | 24b | Indicate where the review protocol can be accessed, or state that a protocol was not prepared. | p. 5 |
|  | 24c | Describe and explain any amendments to information provided at registration or in the protocol. | n/a |
| Support | 25 | Describe sources of financial or non-financial support for the review, and the role of the funders or sponsors in the review. | n/a |
| Competing interests | 26 | Declare any competing interests of review authors. | p. 2 |
| Availability of data, code and other materials | 27 | Report which of the following are publicly available and where they can be found: template data collection forms; data extracted from included studies; data used for all analyses; analytic code; any other materials used in the review. | All data can be obtained from the corresponding author |

Appendix B. Search strategy:

Search terms: (exercise OR training OR physiotherapy OR stabilisation OR stabilization OR strength OR resistance OR flexibility OR sensorimotor OR stretch* OR balance OR endurance OR "physical* activ*") AND ("back pain" OR lumbalgia OR dorsalgia OR backache OR lumbago OR LBP OR "spinal pain") AND ("individualised" OR "individualized" OR "personalised" OR "personalized" OR "stratified" OR "tailored" OR classification OR subclassification OR sub-classification OR sub-group* OR subgroup*) AND (RCT OR "randomized controlled" OR "randomised controlled") NOT surgery [title] NOT herniation [title] NOT orthosis [title] NOT fusion [title] NOT acupuncture [title]

Search terms have been adapted to the respective databases.

Appendix C. Characteristics of included studies

C1. Pain and disability outcomes in included studies

Supplementary table 2. Pain and disability outcomes in included studies. Data is presented as mean (standard deviation). SMT = sensorimotor training, w = weeks

| Author, Year | N individualised | type | Pain Baseline | Pain 12 w | Pain 52 w | Disability baseline | Disability 12 w | Disability 52 w | N controls | type | Pain Baseline control | Pain 12 w control | Pain 52 w control | Disability baseline control | Disability 12 w control | Disability 52 w control |
| --- | --- | --- | --- | --- | --- | --- | --- | --- | --- | --- | --- | --- | --- | --- | --- | --- |
| Aasa, 2015 | 35 | SMT | 4.7 (3.8) | 3 (2.1) | 2.5 (1.6) | 3.8 (3.3) | 7.8 (7.2) | 8 (7.3) | 35 | active control | 4.3 (3.5) | 2.2 (1.4) | 2.4 (1.3) | 4.8 (4.3) | 6.8 (6.1) | 7.3 (6.6) |
| Andersen, 2016 | 46 | resistance | 6.25 (2.65) | 4.78 (2.94) | (4.15) | 3.1 (2.7) | 4.8 (3.1) | (5) | 47 | passive control | 6.47 (1.97) | 6.06 (2.63) | (5.38) | 2.5 (2.4) | 3.4 (3) | (3.4) |
|  |  |  |  |  |  |  |  |  | 47 | true control | 6.21 (2.47) | 5.66 (2.99) | (5.16) | 2.9 (2.8) | 4.2 (3) | (4.8) |
| Apeldoorn, 2012 | 74 | SMT | 6.06 | 4.04 | 3.14 | 1.88 | 1.37 | 1.21 | 82 | active control | 6.11 | 3.61 | 3.2 | 2.25 | 1.67 | 1.35 |
| Azevedo, 2018 | 74 | mixed | 6.61 (1.84) | 3.68 (2.59) | 4.19 (2.95) | 4.43 (2.1) | 2.29 (2.1) | 2.22 (19.4) | 74 | active control | 6.51 (1.92) | 3.86 (3.05) | 3.79 (2.94) | 4.2 (2.1) | 2.24 (2.33) | 2.01 (2.07) |
| Brady, 2018 | 24 | mixed | 7.3 (1.8) |  |  |  |  |  | 24 | active control | 7.4 (1.3) |  |  |  |  |  |
| Cairns, 2006 | 47 | SMT | 5.7 (1.8) |  | 3.6 | 4.3 (1.8) | 2.1 (1.75) | 2.2 (4) | 50 | active control | 5.3 (2.3) |  | 3.1 (2.3) | 4.3 (1.7) | 1.9 (1.7) | 2 (1.7) |
| Cherkin, 1998 | 133 | McKenzie |  |  |  | 1.22 (1.23) | 0.41 (1.96) | (1.1) | 122 | passive control |  |  |  | 1.21 (1.06) | 0.41 (1.88) | (1.76) |
|  |  |  |  |  |  |  |  |  | 66 | advice and usual care |  |  |  | 1.17 (3.1) | 0.43 (2.54) | (2.82) |
| Cuesta-Vargas, 2011 | 25 | mixed | 5.25 (2) | 1.64 (2.44) |  | 2.54 (1.33) | 1.38 (1.33) |  | 24 | active control | 5.76 (1.41) | 2.34 (2.06) |  | 2.17 (1.21) | 1.46 (1) |  |
| Descarreaux, 2002 | 10 | SMT | 0 | -1.45 (0.97) |  | 0 | -1.02 (0.53) |  | 10 | active control | 0 | -0.35 (0.9) |  | 0 | -0.35 (0.65) |  |
| Diaz-Arribas, 2015 | 132 | SMT | 5.4 (4.1) | 4 (2.34) | 4.7 (3.52) | 2.88 (3.81) | 1.96 (2.93) | 2.21 (4.69) | 209 | active control | 5.4 (2.95) | 4.2 (3.32) | 4.3 (2.58) | 3.04 (4) | 2.13 (4) | 2.23 (6.15) |
|  |  |  |  |  |  |  |  |  | 120 | true control | 5.5 (1.84) | 4.6 (4.79) | 4.4 (5.53) | 3.21 | 2.46 | 2.79 |
| Ford, 2016 | 156 | mixed | 5.3 | 3 (2) | 2.5 (2.1) | 2.92 (1.17) | 2.22 (1.29) | 1.45 (1.17) | 144 | advice and usual care | 5.5 (1.9) | 4 (2.3) | 3.5 (2.5) | 2.96 (1.3) | 2.39 (1.51) | 2.04 (1.6) |
| Garcia, 2018 | 74 | McKenzie | 7.19 (1.81) | 3.32 (2.75) | 4.47 (2.73) | 5.16 (2.58) | 3.32 (2.75) | 3.47 (2.63) | 73 | true control | 6.99 (1.73) | 4.18 (2.8) | 5.03 (2.97) | 5.97 (2.43) | 4.13 (2.73) | 4.12 (2.89) |
| Geisser, 2005 | 26 | SMT | 4.45 (2.3) | 2.4 (2) |  | 3.61 (1.41) | 3.11 (1.36) |  | 25 | active control | 5.2 (2.2) | 4.29 (2.7) |  | 5.11 (1.86) | 4.25 (1.93) |  |
|  |  |  |  |  |  |  |  |  | 24 | active control | 3.84 (2) | 3.46 (2) |  | 3.43 (1.96) | 3.33 (1.94) |  |
|  |  |  |  |  |  |  |  |  | 25 | active control | 3.91 (2.5) | 3.39 (2.5) |  | 3.85 (1.6) | 3.18 (1.8) |  |
| Godfrey, 2020 | 124 | mixed | 6 (2.1) | 4.8 (2.5) | 4.8 (2.7) | 4.54 (2.33) | 3.63 (2.67) | 3.42 (2.92) | 124 | active control | 6 (1.9) | 5.3 (2.1) | 4.8 (2.5) | 4.46 (2.42) | 3.04 (2.67) | 3.17 (2.71) |
| Goertz, 2017 | 44 | mixed | 5.3 (1.9) | 3.5 (2.71) |  | 3.75 (2.17) | 3 (1.69) |  | 44 | active control | 6 (1.9) | 3 (1.06) |  | 2.96 (1.83) | 1.71 (1.69) |  |
|  |  |  |  |  |  |  |  |  | 43 | advice and usual care | 6.1 (1.9) | 3.8 (2.88) |  | 2.63 (2.04) | 1.67 (1.86) |  |
| Gudavalli, 2006 | 112 | mixed | 3.57 (2.15) | 2.34 |  | 2.85 (2.03) | 1.89 |  | 123 | passive control | 3.8 (2.2) | 1.74 |  | 2.77 (1.96) | 1.6 |  |
| Hansen, 1993 | 60 | SMT | 4 (1.93) | 2.5 (5.68) | 3 (4.25) |  |  |  | 59 | active control | 4 (1.92) | 3 (1.5) | 3.5 (3.5) |  |  |  |
|  |  |  |  |  |  |  |  |  | 61 | true control | 4.5 (2.22) | 3.5 (1.88) | 3.5 (3.5) |  |  |  |
| Heinrich, 2009 | 76 | mixed+CBT | 5.9 (1.2) | 1.5 (1.2) | 2.1 (1.2) | 3.8 (1.3) | 1.5 (1.3) | 1.6 (1.3) | 125 | advice and usual care | 5.7 (1.8) | 3.28 | 2.2 | 3.6 (1.4) | 1 | 1.6 |
|  | 53 | mixed | 5.9 (1.2) | 2 | 2.3 | 3.7 (1.4) | 1.4 | 1.7 |  |  |  |  |  |  |  |  |
| Henry, 2014 | 77 | SMT | 2.76 (0.22) | 1.59 (0.18) | 1.79 (0.25) | 2.06 (1) | 1.26 (1) | 0.94 (1.08) | 25 | active control | 2.42 (1.8) | 1.42 (1.45) | 1.24 (2) | 1.87 (0.99) | 0.92 (1) | 1.02 (1) |
| Highland, 2018 | 34 | yoga | 4.68 (1.51) | 2.48 (2.34) | 2.79 (2.43) | 3.84 (2.05) | 1.84 (1.95) | 1.35 (1.93) | 34 | advice and usual care | 4.32 (1.61) | 3.67 (1.86) | 2.86 (1.79) | 3.62 (2.01) | 3.17 (2.69) | 2.93 (1.9) |
| Hill, 2011 | 231 | mixed | 5.3 (2.2) | 2.1 (2.5) | 2.3 (2.8) | 4.1 (2.33) | 2.14 (2.46) | 2.31 (2.67) | 283 | advice and usual care | 5.2 (2.2) | 2.6 (2.4) | 2.4 (2.6) | 4.04 (2.42) | 2.79 (2.46) | 2.67 (2.58) |
|  |  |  |  |  |  |  |  |  | 293 | active control | 6.21 (2.32) | 4.18 (2.88) |  | 3.91 (2.32) | 2.68 (2.42) |  |
| Hueppe, 2019 | 189 | mixed | 4.63  (1.91) |  | 3.99  (2.18) | 4.33  (3.5) |  | 3  (3.5) | 255 | advice and usual care | 4.43  (2.04) |  | 4.06  (2.22) | 3.67  (3.67) |  | 3.17  (3.5) |
| Hurley, 2015 | 83 | endurance | 5.46 (2.05) | 4.48 (6.68) | 4.58 (2.16) | 3.48 (1.52) | 3.05 (1.39) | 2.79 (1.39) | 82 | active control | 5.63 (2.06) | 5.17 (2.08) | 5.06 (2.08) | 3.81 (1.41) | 2.99 (1.39) | 2.79 (1.29) |
|  |  |  |  |  |  |  |  |  | 81 | advice and usual care | 5.77 (2.16) | 4.55 (2.08) | 4.74 (2.08) | 3.32 (1.69) | 2.94 (1.39) | 3.05 (1.16) |
| Jay, 2015 | 12 | mixed | 3.7 (2.6) | 2 (2.6) |  |  |  |  | 12 | advice and usual care | 3.1 (3.1) | 3 (3.1) |  |  |  |  |
| Jensen, 2011 | 176 | mixed | 3.16 (1.21) |  | 2.09 (1.2) | 6.54 (1.92) |  | 3.71 (1.9) | 175 | advice and usual care | 3.27 (1.24) |  | 2.14 (1.2) | 6.5 (2.17) |  | 3.54 (2.2) |
| Kim, 2020 | 20 | SMT | 4 (1.56) | 1.58 (0.84) |  | 1.13 (0.54) | 0.51 (0.41) |  | 19 | advice and usual care | 3.9 (1.54) | 3.35 (1.53) |  | 1.29 (0.67) | 1.05 (0.64) |  |
| Lang, 2021 | 117 | streching | 2.2 (1.8) | 2 (1.66) | 1.8 (1.66) | 2.05 (1.17) | 1.51 (1.1) | 1.31 (1.38) | 57 | advice and usual care | 2.4 (1.4) | 2.8 (2.31) | 2.4 (2.7) | 2.12 (0.98) | 1.88 (1.16) | 1.68 (1.27) |
| Lehtola, 2016 | 35 | SMT |  |  |  | (0.5) | (0.85) | (0.6) | 35 | active control |  |  |  | (0.46) | (1.09) | (1.06) |
| Leibetseder, 2007 | 22 | endurance |  |  |  |  |  |  | 22 | active control |  |  |  |  |  |  |
| Lomond, 2015 | 21 | SMT | 3.6 (1.6) |  |  | 1.99 (0.94) |  |  | 12 | active control | 2.8 (1.6) |  |  | 1.72 (0.79) |  |  |
| Long, 2004 | 80 | McKenzie | 5.86 (2.39) | 2.51 (1.96) |  | 7.44 (2.34) | 4.74 (3.15) |  | 70 | active control | 6.08 (2.17) | 4.65 (2.33) |  | 6.95 (2.49) | 6.43 (2.88) |  |
|  |  |  |  |  |  |  |  |  | 80 | advice and usual care | 5.97 (2.06) | 4.34 (2.51) |  | 7.65 (2.23) | 6.44 (4.06) |  |
| Macedo, 2012 | 86 | SMT | 6.1 (2.1) | 4.1 (2.5) | 4.1 (2.7) | 4.67 (2.21) | 3.33 (1) | 3.58 (1) | 86 | active control | 6.1 (1.9) | 4.1 (2.5) | 4.1 (2.5) | 4.75 (2) | 3.13 (0.88) | 3.33 (0.96) |
| Magalhães, 2018 | 33 | mixed | 7.2 (1.9) | 2.4 (1.4) | 4.1 (2.1) | 5.38 (2.03) | 2.71 (1.75) | 3.08 (2.08) | 33 | active control | 7.6 (2) | 2.5 (1.9) | 4.1 (3.1) | 5.33 (2) | 2.63 (2.29) | 3.08 (2.5) |
| Michaelson, 2016 | 35 | endurance | 4.7 (2.8) | 3 (2.6) | 2.5 (2.2) | 2.96 (1.63) | 1.5 (1.75) | 1.38 (1.5) | 35 | passive control | 4.3 (2.4) | 2.2 (2.1) | 2.4 (2.7) | 3 (1.79) | 1.58 (1.67) | 1.5 (1.75) |
| Moore, 2000 | 113 | back school | 5.4 (1.89) | 3.69 (2.05) | 3.15 (2.14) | 3.58 (2.7) | 2.25 (2.4) | 2.18 (2.5) | 113 | advice and usual care | 5.2 (1.95) | 4.06 (2.17) | 3.71 (2.28) | 3.45 (2.45) | 2.73 (2.56) | 2.67 (2.5) |
| O’Keeffe, 2020 | 106 | mixed | 6.17 (2.17) | 2.91 (2.47) | 3.77 (2.72) | 3.21 (1.26) | 1.62 (0.97) | 2.02 (1.55) | 100 | active control | 5.69 (2.23) | 4.6 (2.39) | 4.44 (2.36) | 3.35 (1.26) | 2.61 (1.4) | 2.85 (1.7) |
| Paolucci, 2011 | 29 | back school | 6.5 (3) |  |  |  |  |  | 21 | medication | 7.5 (1.5) |  |  |  |  |  |
| Petersen, 2011 | 175 | McKenzie | 3 (1.12) | 1.47 (1.12) | 1.5 (1.1) | 5.42 (2) | 2.71 (2) | 2.46 (2) | 175 | passive control | 2.9 (1.13) | 1.52 (1.1) | 1.6 (1.1) | 5.42 (2.08) | 3 (2) | 3.25 (2) |
| Rabiei, 2021 | 37 | SMT | 6.45 (1.21) | 3.79 (1.02) |  | 6.08 (1.06) | 3.31 (1.36) |  | 36 | active control | 6.36 (1.14) | 4.91 (2.17) |  | (1.35) | (1.35) |  |
| Rabin, 2014 | 48 | SMT | 4.9 (1.7) | 2.4 (1.8) |  | 3.78 (1.06) | 1.61 (1.12) |  | 57 | active control | 5.3 (1.7) | 3.1 (2.5) |  | 3.76 (1.25) | 2.02 (1.6) |  |
| Rasmussen-Barr, 2009 | 36 | SMT | 3.2 (6.28) | 1.7 (6.19) | 1.65 (6) | 2 (2.14) | 1.3 (2.11) | 1.1 (2) | 35 | advice and usual care | 3.8 (6) | 3 (6) | 2.9 (6) | 2.2 (2) | 1.8 (2) | 1.8 (2) |
| Salas, 2019 | 8 | mixed | 5.6 (0.77) | 5.06 (0.77) |  | 5.39 (0.72) | 5.19 (0.77) |  | 8 | active control | 5.75 (0.75) | 6.06 (0.49) |  | 6.05 (0.43) | 6.19 (0.5) |  |
| Sandal, 2021 | 232 | mixed | 4.8  (2) | 3.3  (2.2) | 3  (2.3) | 4.29  (1.83) | 2.79  (1.96) | 2.5  (2.21) | 229 | advice and usual care | 4.9  (1.9) | 3.9  (2.4) | 3.7  (2.4) | 4.42  (1.83) | 3.08  (2.25) | 2.88  (1.83) |
| Saner, 2015 | 52 | mixed | 4.83 | 3.37 | 2.9 | 3.79 | 1.88 | 1.79 | 54 | active control | 5.5 | 4.33 | 3.33 | 3.42 | 2.21 | 1.88 |
| Schaller, 2016 | 201 | mixed | 4.5 (1) | 3.5 (1.3) |  |  |  |  | 211 | passive control | 4.6 (0.9) | 3.7 (1.2) |  |  |  |  |
| Soukup, 1999 | 34 | mixed | 4.1 (1.5) | 2.3 (1.6) | 2.6 (1.9) |  |  | 4.2 | 35 | advice and usual care | 2.4 (2.1) | 3.2 (1.7) | (2.3) |  |  |  |
| Suh, 2019 | 15 | SMT | 3.75 (2.18) | 2.42 (2.02) | 2.25 (2.37) | 3.14 (1.32) | 2.53 (0.99) |  | 15 | active control | 4.19 (2.33) | 2.79 (1.85) | 3.25 (2.18) | 3.8 (2.11) | 3.19 (1.86) |  |
|  | 15 | mixed | 3 | 1.83 | 2.08 | 2.94 | 4.43 |  | 15 | active control | 3.06 (2.09) | 2.5 (2.01) | 2 (1.76) | 2.81 (1.28) | 2.25 (0.74) |  |
| Thanawat, 2017 | 62 | mixed | 3.77 (1.66) | 2.7 (1.38) | 2.03 (0.76) | 2.44 (0.86) | 1.48 (0.48) | 1.1 (0.34) | 64 | active control | 4.14 (1.82) | 2.8 (1.1) | 3.04 (0.71) | 2.87 (1.16) | 1.66 (0.87) | 1.67 (0.48) |
| Tsauo, 2009 | 20 | mixed | 5.9 (1.8) | 2.8 (1.8) |  | 2.2 (0.9) | 1.6 (0.9) |  | 17 | active control | 5.25 (1.6) | 4.6 (1.65) |  | 1.3 (0.6) | 1.3 (0.6) |  |
| Van Baal, 2020 | 18 | SMT | 3.5 (1.48) | 1.13 (2.22) |  | 2.2 (1.48) | 1.1 (1.48) |  | 16 | active control | 3 (1.48) | 2 (1.63) |  | 2 (1.04) | 1.6 (0.74) |  |
| Van Dillen, 2016 | 47 | SMT |  |  |  |  |  |  | 54 | active control | (3.04) | (2.09) | (2.64) | (1.29) | (1.23) |  |
| Vasseljen, 2012 | 36 | SMT | 3.4 (1.3) |  |  | 2 (0.72) |  |  | 36 | active control | 3.5 (1.8) |  |  | 1.99 (0.9) |  |  |
|  |  |  |  |  |  |  |  |  | 37 | active control | 3.1 (1.6) |  |  | 1.96 (0.83) |  |  |
| Verra, 2018 | 107 | mixed | 6.32 (2.09) | 5.66 (2.68) | 5.63 (2.46) | 4.61 (1.31) | 4.39 (1.39) | 4.24 (1.49) | 32 | active control | 6.32 (1.96) | 5.75 (2.4) | 5.89 (2.68) | 4.46 (1.33) | 4.33 (1.68) | 4.41 (1.44) |
| Vibe Fersum, 2013 | 62 | mixed | 4.9 (2) | 1.7 (2) | 2.3 (3.21) | 2.13 (0.75) | 0.76 (0.75) | 0.99 (0.75) | 59 | active control | 5.3 (1.9) | 3.8 | 3.8 | 2.4 (0.8) | 1.85 | 1.97 |
| Von Korff, 2005 | 119 | back school | 5.7 (1.8) | 4.9 (2) | 4.2 (2) | 5.13 (2.29) | 4.25 (2.63) | 3.83 (2.75) | 121 | advice and usual care | 5.8 (1.8) | 5.3 (1.9) | 4.7 (2.2) | 4.75 (2.38) | 4.79 (2.42) | 4.21 (2.67) |
| Wajswelner, 2012 | 44 | pilates | 4.9 (1.6) | 2.8 (1.6) | 2.5 (2.3) | 2.81 (1.14) | 1.53 (0.91) | 1.41 (1.04) | 43 | active control | 4.6 (1.8) | 3.2 (2.1) | 2.4 (1.7) | 2.39 (1.4) | 1.71 (1.34) | 1.4 (1.53) |
| Wälti, 2015 | 14 | SMT | 4.86 (1.61) | 2.72 (1.61) |  | 4.25 (1.85) | 1.45 (1.85) |  | 14 | active control | 4.64 (1.81) | 3.95 (1.81) |  | 4.67 (1.64) | 2.72 (1.6) |  |

C2. Characteristics of interventions and individualisation

| **Author, Year** | **Individ-ualised intervention** | **detailled intervention** | **cognitive-behavioural component** | **controls** | **detailled controls** |
| --- | --- | --- | --- | --- | --- |
| **Aasa, 2015** | SMT | low-load motor control (LMC) exercise: The physical therapist took a detailed anamnesis, performed a physical examination, and selected individual exercises accordingly. The exercises aimed to normalize the dominating movement impairment for each participant. The strategy was to start from a basic level and continue to a gradually increased level of difficulty. | none | active control | high-load lifting (HLL) exercise: we chose the deadlift exercise, which activates the stabilizing muscles and focuses on main tenance of an optimal alignment of the spine during the lift. The physical therapist selected appropriate initial weight on the bar, based on the anamnesis and findings in the physical examination. The load was slowly progressed during the intervention period by gradually increasing the number of lifts and/or the weight on the bar. The participants were encour aged to use the same lifting technique during daily activities. |
| **Andersen, 2016** | **mixed** | Tailored Physical Activity: teams of up to 10 participants, a standardized combination of aerobic fitness and strength training supervised by physiotherapists. It started with awarm-up, followed by aerobic fitness training. After that the participants were referred to 1 of 3 standardized strength training programmes based on their primary region of musculoskeletal problems (neck and shoulder pain; arm and/or hand pain; lower back pain). During the following weeks, training and progression was tailored to the participant’s current training status and pain problems. The physiotherapists used their professional judgement to match demands in individual programmes. | Health guidance was a 1.5-h dialogue with a health supervisor, centred around the participants’ lifestyle, motivation, resources and power to act. The participant was offered the chance to prepare a health plan, and the health supervisor provided ideas and support for increasing well-being in everyday life. | C1: passive control | health guidance only |
|  |  |  |  | C2: active control | This active comparator group received Chronic Pain Self-Management Program in addition to health guidance, it is a standardized programme of 2.5 h in a weekly workshop lasting 6 weeks. Workshops were led by 2 trained facilitators (non-health professionals) who had chronic pain. Topics covered in the teaching sessions included techniques to deal with problems, such as fatigue, exercises, the use of medications and communication tools. Classes built the participants’ confidence to manage their own health and to help them stay active in their daily live. |
| **Apeldoorn, 2012** | SMT | treated according to their primary classification category ( i.e., direction-specific exercises, spinal manipulation, or stabilization exercises) for a minimum of 4 weeks. After this period, the physical therapist was allowed to change treatment strategy according to the current Dutch LBP guidelines. | none | active control | Patients assigned to usual physical therapy care received individually tailored treatment according to the current Dutch LBP guidelines. |
| **Azevedo, 2018** | mixed | Treatment based on the MSI model included (1) patient education, (2) analysis and modification of performance of daily activities, and (3) prescription of specific exercises. The prescription of specific exercises included performing modifications of the movement tests from the initial assessment per the participant’s LBP classification. | I: education | active control | Treatment consisting of symptomguided stretching and strengthening exercises. After walking or pedaling a stationary bicycle for 5 minutes, participants performed stretching exercises addressing the lumbar and abdominal muscles |
| **Brady, 2018** | mixed | Culturally adapted physiotherapy assessment and treatment Participants received a combination of group and individual physiotherapy sessions, adapted to reflect the ethnocultural beliefs and values of the community to which the participant identified. Sessions were delivered once per week for 6 weeks, included a combination of education and exer cise | education | active control | evidence-informed ‘usual physiotherapy care’ Participants allocated to this condition attended physiotherapy in the outpatient department where they were referred, for treatment informed by evidence-based recommendations for chronic pain. All treating physiotherapists underwent a training session to familiarise them with evidence-based management of chronic pain. Treatment adherence to these guidelines was monitored by review of therapist treatment logs. Treating physiotherapists used their clinical judgement to guide the specifics of treatment according to principles of patient-centred care.2 |
| **Cairns, 2006** | SMT | Specific Spinal Stabilization Exercise Group. Endurance training for the deep abdominal and back extensor muscles was the predominant component of this treatment group. A treatment manual for clinicians outlined appropriate exercise progression, but treatment was individualized at the discretion of the clinician. A patient booklet was developed to emphasis the specific nature of the exercises, outlining anatomy and function of the muscles and the concept of endurance training. The majority of patients received manual therapy, such as Maitland mobilizations, exercise and advice, with little use of electrotherapy or mechanical lumbar traction. | standardized educational information based on the best available evidence regarding continuing normal activities and avoiding rest (The Back Book) | active control | Conventional Treatment Group. Exercises using low load, high repetition muscle activity were excluded. All participating departments had adopted an active approach to back pain management, with encouragement to remain active and the minimal use of more “passive” forms of treatment. |
| **Cherkin, 1998** | McKenzie | McKenzie approach, patients are placed in one of three broad categories (derangement, dysfunction, and postural syndrome) that determine therapy. Subjects received McKenzie’s Treat Your Own Back book19 and a lumbar-support cushion. Therapists were asked to avoid adjuncts such as heat, ice, transcutaneous electrical nerve stimulation, ultrasonography, and back classes. | none | C1: passive control C2: usual care | C1: Chiropractic manipulation : a short-lever, high-velocity thrust directed specifically at a “manipulable lesion.” No other hiropractic manipulation C2: A minimal-intervention control group received an educational booklet to minimize potential disappointment with not receiving a physical treatment. The booklet discussed causes of back pain, prognosis, appropriate use of imaging studies and specialists, and activities for promoting recovery and preventing recurrences. |
| **Cuesta-Vargas, 2011** | endurance | Multimodal physical therapy program with additional individualised aerobic exercise in the form of a 20-min session of deep water running (DWR) at the aerobic threshold (AT). The individual workload of the AT was estimated during the deep-water running test. A physiotherapist supervised the intensity and technique during DWR in a group session of ten subjects with individual workloads. | pain education, and information on an active lifestyle | active control | The multimodal physical therapy program was was individualized. In addition to the clinical and functional outcome measures described previously, an individual evaluation consisting of a clinical, occupational, and physical activity; a semistructured interview; and a physical examination was performed by a physiotherapist blinded to the trial. |
| **Descarreaux, 2002** | SMT | home exercise program: prescription for force and extensibility exercises was based on the initial evaluation. Exercise for the experimental group targeted increased muscular force and extensibility of trunk and hip muscles. Exercises and training volume (quantity and intensity) were chosen in relation to the initial deficit. | none | active control | In contrast, every participant of the control group received the same exercise program. This program was based on the classical “back school” recommendation for low back exercising.34 This program included 5 exercises: (1) flexion mobilization exercises (erect position), (2) passive extension mobilizations, (3) stretching exercises of the erector spinae (supine position), (4) abdominal reinforcement exercises, and (5) combined back and hip extension exercises. Volume and training intensity were the same for every subject |
| **Diaz-Arribas, 2015** | SMT | Godelieve Denys-Struyf method (GDS) is a motor control intervention that classifies muscles influencing lumbarpelvic and spinal stability into 6 groups (“muscle chains”), according to their anatomy and role in postures and movements. Participants received the collective sessions provided to the true control group. Additionally, each participant was physically examined for 20 minutes and received 4 additional 50-minute individualized, one-on-one sessions of manual therapy, stretching, and massage focusing on the muscle groups (chains) determined to require more attention in that particular case. | Short education program on active management, comprising a 15-minute talk given to groups of no more than 15 participants and the delivery of the Spanish version of the “back book.” | C1: true control | C1: 11 collective GDS sessions were provided to groups of 10 to 12 participants. These sessions focused on the muscle imbalances that are most commonly found in patients with LBP |
|  |  |  |  | C2: active control | C2: physical therapy regimen implemented was the one routinely used within the department; it consisted of fifteen 40-minute sessions, applied twice per week, and included microwave treatment, transcutaneous electrical nerve stimulation, and standardized exercises. The exercises were implemented progressively across sessions, in accordance with the physical therapist’s criteria, and expected to be continued at home. |
| **Ford, 2016** | mixed | physiotherapy that was individualised based on pathoanatomical, psychosocial and neurophysiological barriers to recovery combined with guideline-based advice (10 sessions) | none | advice and usual care | 2 sessions of physiotherapist-delivered advice alone |
| **Garcia, 2018** | McKenzie | McKenzie Method of mechanical diagnosis and therapy (MDT) prescribes repeated exercises in a specific direction, combined with an educational approach to treat patients with mechanical pain. All patients also received a translated version of ‘The Back Book | education | true control | placebo group treated with detuned pulsed ultrasound for 5 min with patients in side lying. They also received detuned short wave diathermy in pulsed mode for 25 min (in a supine position). The devices were used with the internal cables disconnected to obtain the placebo effect. All patients also received a translated version of ‘The Back Book |
| **Geisser, 2005** | SMT | Patients were assigned to a specific adjuvant exercise program (SE) designed to help improve specific musculoskeletal dysfunctions observed during the standardized manual medicine screening evaluation. Specific exercises were taken from Sahrman and Bookhout and included self-corrections, stretches, and strengthening exercises. | All patients watched a 12-minute videotape that provided educational information on musculoskeletal pain and oriented patients on how exercise might be beneficial in terms of improving their pain. | all active controls | Patients received nonspecific exercises. These exercises were not designed to treat specific musculoskeletal dysfunctions, as they did not target stretching or strengthening dysfunctional muscles or improving joint mobility in a restricted area. Patients in each group received either manual therapy (MT) or were administered a sham manual therapy procedure (sham MT). |
| **Godfrey, 2020** | mixed | brief physical therapy intervention, guided by principles of ACT, designed to promote selfmanagement. PACT consisted of 3 individual treatment sessions as follows: two 60-minute face-to-face sessions 2 weeks apart conducted in a private room, plus one 20-minute telephone call 1 month later. Treatment included an initial physical assessment with feedback, identification of value-based goals, individualized physical exercise prescription, addressing barriers and facilitators to self-management, and skills training to promote psychological flexibility. It excluded manual therapy. T | education | advice and usual care | physical therapy as part of usual care |
| **Goertz, 2017** | mixed | individualized chiropractic care that included clinical history and exams and self-care recommendations, including exercises. collaborative medical and chiropractic care (Shared Care): to enhance interdisciplinary communication and practice through interprofessional education, clinical record sharing, and team-based case management | education | C1: active control | C1: concurrent medical and chiropractic care |
|  |  |  |  | C2: advice and usual care | C2: guideline-based medical care from a study-assigned resident physician |
| **Gudavalli, 2006** | mixed | active trunk exercise protocol (ATEP) administered by licensed physical therapist, consisted of flexion or extension exercises, weight training, flexibility exercises, and cardiovascular exercises dependent on patient symptoms. The aim of the program was to strengthen the muscles surrounding the spine and increase flexibility. Further individualisation was possible, dependent on clinically relevant symptoms. | none | passive control | a series of flexion–distraction procedures administered by licensed chiropractors |
| **Hansen, 1993** | SMT | Standardized physical therapy {PT). An obligatory and an individual program were used. The obligatory program consisted of soft-tissue treatment, manual traction, flexibility exercises for the lumbar spine and the pelvis, ergonomics counseling, exercises for coordination, and a slowly progressive exercise program that included isometric back and abdominal muscle exercises. The individual program was designed according to the primary objective examination | none | C1: true control | Placebo-control {CTRL). After resting 20 minutes on semihot packs, patients were given intermittent traction (Tru Trac)-7 seconds of gradual traction and 7 seconds of relief-for the next 20 minutes. A force equivalent to 10% of the body weight was used. During the last 20-min sequence, the patient rested on the packs, which were not reheated. |
|  |  |  |  | C2: active control | Intensive dynamic back-muscle training {DYN). Three exercises were performed: 1) trunklifting in prone position; 2) leg-lifting in prone position; and 3) pull down to the neck. |
| **Heinrich, 2009** | mixed | I1: Physical Training with a Cognitive Behavioural component and Workplace specific Exercises (referred to as PTCBWE) The physical training component did not differ from the intervention described above regarding PT except the fact that during PTCBWE co-intervention (e.g. physiotherapy) was not allowed. | I1: cognitive-behavioral component to detect dysfunctional thinking habits and to change those thinking habits into a more realistic or functional way of thinking (e.g. reconceptualisation of pain) | C2: advice and usual care | usual guidance by their general practitioner according to the guidelines of the Dutch College of General Practice for musculoskeletal disorders |
|  |  | I2: Physical Training without a cognitive behavioural component and workplace specific exercises (referred to as PT) The physical training took place two or three times a week, for 1–1.5 hours, during three months, also if someone had already fully returned to work again. It consisted of cardiovascular training, strengthening, relaxation exercises and posture exercises. Because the physical training exists of multiple components, a general level of intensity, which represents the whole training, can not be described. The level of intensity for every component was decided during an intake (as described below). For every participant an individual level of intensity and gradually increase schedule was determined. |  |  |  |
| **Henry, 2014** | SMT | Movement System Impairment-based treatment: Based on the subject’s direction-specific LBP classification, the PT tailored the Movement System Impairment protocol to focus on: (1) education about positions or postures to control his symptoms; (2) ‘Exercises for Precision of Trunk Movement” where patients were taught specific trunk movements and postures that were painfree; and (3) functional activity modifications (based on their Patient Specific Functional Scale) to change his trunk-movement and alignment patterns | none | active control | stabilization exercises ocused on 3 components of spinal stability: (1) motor control of the deep trunk muscles; (2) strengthening of the flexor, extensor, and oblique trunk muscles by focusing on repeated submaximal efforts to mimic the function of these muscles in spine stabilization and (3) patient education in the form of an education booklet |
| **Highland, 2018** | yoga | RESTORE is based on therapeutic yoga, targeting major muscles affected by chronic LBP including back and core strengthening/stretching for postural alignment. RESTORE participants were asked to attend two individual yoga sessions. Each session included breath-work and centering (10 minutes), poses based on the ability to intensify or modify each pose (40 minutes), and a guided final meditation (10 minutes). Depending on instructor judgement and participant receptivity, participants used props (e.g., blocks, straps) and repeated lifting and lowering, combined lifting/lowering with holding for9 five breaths, or held for up to ten breaths for each pose. | none | advice and usual care | n/a |
| **Hill, 2011** | mixed | decisions about referral were made by use of the STarT Back Screening Tool classification. with advice focusing on promotion of appropriate levels of activity, including return to work, and a pamphlet about local exercise venues and self-help groups. Participants were shown a 15-min educational video entitled Get Back Active20 and given the Back Book. | education | active control | baseline clinical assessment and treatment session, decisions about referral were made on the basis of the physiotherapists’ clinical judgment, without knowledge of a participant’s STarT Back Tool classifi cation. Participants received a 30-min physiotherapy assessment and initial treatment including advice and exercises, with the option of onward referral to further physiotherapy. |
| **Hill, 2020** | mixed | Stratified care intervention using the prognostic stratification tool (a development version of the Keele STarT MSK tool) and recommended matched treatment options. | education | advice and usual care | received clinical care as usual for MSK pain. Usual primary care is known to be variable; for example, some patients may receive advice, prescriptions for medications and nothing more, some may be asked to return to the GP for follow-up assessment or treatment, whereas others may be referred to other services, including for tests and investigations, or treatment services such as Hill et al. BMC Family Practice (2020) 21:30 Page 3 of 18 physiotherapy, orthopaedics or pain clinics. |
| **Hueppe, 2019** | mixed | comprehensive health program comprising medical exercise therapy and life style coaching. Tailor-made therapy program for the back muscles, safe from a medical point of view, was put together for each participant in specialized back centers. | intervention group: received personal health coaching over the phone from an external professional coach. Coaching aimed at encouraging life style changes and the consolidation of physical activities. | usual care and advice | longitudinal back pain survey and care according to the prescriptions of their health care providers (family doctors or medical specialists). |
| **Hurley, 2015** | endurance | Walking programme was individualized to each participant, with the focus being to increase PA through a graded volume-based WP based on the American College of Sports Medicine (ACSM) guidelines. participants were provided with an educational walking booklet developed for the trial that detailed the aim of the programme, health benefits of walking, correct walking technique, and advice on appropriate clothing and footwear. It was aimed to progress to the ACSM-recommended levels (30 minutes of moderate-intensity PA for 5 days per week) by week 5. the treating physiotherapist made weekly contact with each participant via telephone to evaluate and progress their weekly walking prescription according to their achievement | none | C1:active control | Exercise class followed a group-based circuit format based on the “Back to Fitness” programme endorsed by the UK NICE guidelines. Each class consisted of a programme of progressive or graded exercises, and a back care education message in the form of a “Tip for the Day.” The exercise components included warm-up and stretching, up to 10 individual exercises (3 levels of difficulty progressed as appropriate of aerobic, trunk, upper limb, and lower limb strengthening), cool down, and relaxation. |
|  |  |  |  | C2: advice and usual care | Usual physiotherapy was defined as a combination of individualized education/advice, exercise therapy, and manipulative therapy at the discretion of the treating physiotherapist based on usual practice in the Republic of Ireland.7 |
| **Jay, 2015** | mixed | ysical/ mindfulness group-based training consisted of 4 major elements: 1) individualized motor control training, 2) individualized resistance training specific to the pain affected area 3) cognitive and behavioral modification education emphasizing individual specific concerns about pain and movement, and 4) general mindfulness. I | cognitive and behavioral modification education focusing on pain de-catastrophizing and fear-avoidance beliefs . Additionally, the psychological elements also involved pain management education and information also grouped in the physical training sessions. | advice and usual care | received a single email after randomization with encouragement to par ticipate in the company’s on-going health initiatives, e.g., weekly elastic band group training sessions (only available in some departments) and was encouraged to continue to take “active breaks” whenever needed. As this is part of the existing and currently on-going program at the company it can be considered “usual care.” |
| **Jensen, 2011** | mixed | Multidisciplinary Intervention:standardised interview with a case manager that included questions of work history, private life, and questions on how pain and disability was perceived. It normally lasted for 1 to 2 hours. The participant was seen once or more times by the case manager depending on need and progress. The case manager and the participant together made a tailored rehabilitation plan aiming at full or partial return to work. Each case was discussed several times by the entire multidisciplinary team including the rehabilitation physician, a specialist in clinical social medicine, a physiotherapist, a social worker, and an occupational therapist | cognitive and behavioural education as part of the multidisciplinary setting | advice and usual care | Brief Intervention: tandard clinical LBP examination , The participants were advised to resume work when possible. The physiotherapy examination included a standardized, mechanical evaluation, and advice on exercise was chosen accordingly. General advice was given to increase physical activity and exercise. |
| **Kim, 2020** | SMT | classification-specific treatment (based on the movement-system impairment (MSI) classification system), which included exercise to control or prevent lumbopelvic motion during lower-extremity movement | education | advice and usual care | encouraged to perform general exercises and were educated about LBP |
| **Lang, 2021** | **endurance** | participants in the walking group were prescribed a personalized pedometer-driven walking program wearing a pedometer. the walking program was tailored on a week by week basis to the individual | education | advice and usual care | standard package of education and advice |
| **Lehtola, 2016** | SMT | Participants were taught the Specific movement control exercises and advised on the intensity at which they should exercise. The exercises were performed under supervision of a physical therapist. The participant performed the previously taught exercises and the physical therapist corrected the performance when necessary. Home exercises were to be performed | none | active control | Participants were taught the exercises and advised on the intensity of performance. The exercises were performed under supervision of a physical therapist. The intensity of the exercises was progressed over the 5 treatments sessions, with participants being encouraged to improve their own performance. |
| **Leibetseder, 2007** | endurance | performed additional aerobic training. Depending on the relative cardiorespiratory fitness the quantity of aerobic training was individually set. Training intensity was adjusted to 60% of the individual heart rate reserve | none | active control | atients stayed in the spa resort at Bad Tatzmannsdorf for the entire duration of the study (3 weeks) and received 2–4 therapies daily except sundays. We applied mud applications, carbon dioxide baths, classic massages, under water massages, exercise therapy, and spinal traction |
| **Lomond, 2015** | SMT | tailored the MSI protocol to focus on (1) education regarding how the subject’s lumbopelvic movement patterns and postures repeated daily might accelerate lumbar tissue stress as well as education about positions or postures to control symptoms, (2) exercises to modify the subject’s specific trunk movements and postures in particular directions that were pain free, and (3) functional activity modifications (based on their Patient Specific Functional Scale) to change the subject’s trunk movement and alignment patterns | education | active control | The physical therapist (PT) progressed subjects through the standardized treatment protocols, combined with a home exercise program |
| **Long, 2004** | McKenzie | Subjects were taught unidirectional end-range lumbar exercises matching the direction of their DP identified during baseline assessment. | Both control groups were provided education consistent with LBP clinical guidelines, including advice aimed at minimizing fear avoidance behavior and to remain active. Members of the intervention group were likewise instructed to remain active but also to avoid activities and positions that increase intensity or radiation of symptoms. | C1: active control | Subjects were also taught unidirectional end-range exercises, but in a direction opposite to their DP identified during baseline assessment. |
|  |  |  |  | C2: advice and usual care | Evidence-based care (EBC): Subjects were taught commonly prescribed multidirectional, midrange lumbar exercises, and stretches for the hip and thigh muscles. |
| **Macedo, 2012** | SMT | motor control exercise program was based on the treatment program reported by Hodges. A primary goal of the exercise was to enable the patient to regain control and coordination of the spine and pelvis using principles of motor learning such as segmentation and simplification. The intervention was based on assessment of the individual participant’s motor control impairments and treatment goals (set collaboratively with the therapist). | Cognitive-behavioral principles were used to help the participants overcome the natural anxiety associated with pain and activities. | active control | graded activity program was based on the treatment program originally reported by Lindstrom and similar to the protocol previously used by Pengel and Smeets. A primary goal of the program was to increase activity tolerance by performing individualized and submaximal exercises, |
| **Magalhães, 2018** | mixed | The graded activity group followed the program described by Macedo and Smeets and was based on individual sessions of progressive and sub-maximal exercises aimed at improving physical fitness and stimulate changes in behavior and patients’ attitudes toward pain. The program consisted of aerobic training on a treadmill and lower limb strengthening exercises. | none | active control | physiotherapy exercise group was based on the protocol reported by Franca. The program comprised of stretching exercises of main muscle groups (e.g. erector spinae, hamstrings, and triceps surae), strengthening exercises (e.g. abdominal curl-ups, trunk extension), and motor control exercises (e.g. low load exercises of deep trunk muscles such as tranversus abdominus and multifidus). The physiotherapy exercise group did not receive hands-on interventions such as manual therapy techniques. All sessions had the same protocol exercise, with no progression of exercise levels implemented. |
| **Michaelson, 2016** | **SMT** | Low load motor control exercises to retrain identified faulty movement patterns. The choice of the exercises was based on the anamnesis and physical examination performed to identify provocative and relieving postures and movements. The intervention was divided into 3 stages. In the first stage, the participants’ ability to activate the stabilizing muscles in order to control the lumbar spine in neutral positions was retrained. In the second stage, the participant was assessed and evaluated on provocative and relieving movements using postural correction exercises including static control. In the third stage, an implementation of the desired movement pattern into various dynamic tasks and functional positions used in everyday life was performed, based on activities the participants reported to be provocative. After each treatment session, the participant received 1–3 home exercises to perform each day until the next appointment | none | **active control** | High load lifting exercise intervention consisted of the dead-lift exercise that efficiently activates the stabilizing muscles of the lower back through optimal alignment of the spine. The load was gradually increased during the intervention period |
| **Moore, 2000** | back school | The intervention consisted of two two-hour group sessions, with 12±16 participants, led by one of two psychologists experienced in chronic pain management. Within 2 weeks following the group sessions, each participant met individually with his or her leader for approximately 45 min to develop a personal self care plan. Leaders made one brief (approximately 3 min) follow-up telephone call to each participant to encourage continued action on the self care plan. | cognitive-behavioral self care program. The groups were conducted according to a fully structured protocol. Standardized information was presented via brief lectures, flip charts, and visual aids. Topics included back anatomy, the `red flags' indicating a serious medical condition, the more common (and less worrisome) causes of back pain, factors contributing to fluctuations in pain, appropriate pacing of exercise and activity, basics of posture and body mechanics, cognitive restructuring, handling back pain flare-ups and working with health care providers. | advice and usual care | n/a |
| **O’Keeffe, 2020** | mixed | Cognitive functional therapy, all participants underwent a comprehensive one-to-one interview and physical examination by their phys iotherapist, to identify any relevant multidimensional factors considered to be key drivers of their pain and disability. The length of the intervention varied in a pragmatic manner based on the clinical progression of participants. There were then three components to the intervention: (1) cognitive component: making sense of pain; (2) exposure with ‘control’; and (3) lifestyle change | cognitive-behavioural setting | active control | Group-based exercise and education intervention did not involve any individual interview, physical examination or consideration of the patient’s detailed pain story. All participants in this intervention received a multidimensional intervention addressing the same principles of rehabilitation, but this was not specifically targeted to their individual needs or presentation. There were three components to the intervention: (1) pain education; (2) exercise; and (3) relaxation |
| **Paolucci, 2011** | back school | Back School program was an intensive four weeks intervention carried out by a multidisciplinary professional team. It was conducted in a rehabilitation center and formed by 10 intervention sessions. | behavioural counceling | medication | Control was undertaken to medical treatment (NSAIDs and myorelaxant) self administered during the period of this study under physician supervision similarly to the treatment group. Physicians were instructed to not start or use any new therapy during the study using different drugs (antidepressants, antiepileptics or other) and if necessary patients were dropped out. |
| **Petersen, 2011** | McKenzie | cKenzie treatment was planned individually after the therapist’s pretreatment physical assessment. Manual vertebral mobilization techniques including high velocity thrust were not allowed. An educational booklet describing self care or a “lumbar roll” for correction of the seated position was sometimes provided to the patient at the discretion of the therapist. | All patients were informed thoroughly of the results of the physical assessment, the benign course of back pain, and the importance of remaining physically active. Guidance on proper back care was also given. ll patients were provided with a Danish version of “The Back Book,” | passive control | spinal manipulation treatment, all types of manual techniques including vertebral mobilization and high velocity thrust as well as myofascial trigger-point massage were used. The choice of technique, or combination of techniques, was at the discretion of the chiropractor dependent of the results of their pretreatment physical assessment. G |
| **Rabiei, 2021** | SMT | motor control exercise rogram provided in this study was identical to that described by Macedo. Sixteen sessions (twice a week) provided to the patients for eight weeks. In the first session, the patients were individually examined by the physical therapist, and prescribed exercises were based on the patients’ tolerance/ability. The intended exercises were designed in 2 parts with specific criteria met by each patient. Additional Pain neuroscience education | Intervention group with Additional Pain neuroscience education aimed to reconceptualize the patients’ negative beliefs about the pain. also targeted to reduce fearavoidance beliefs and avoidance behavior, and consequently to promote self-efficacy (cogbitive behavioural). | active control | group-based exercise performed low back strengthening exercises for 16 sessions (twice a week for eight weeks) under the supervision of a well-experienced physical therapist: specifically, a 10-minute group-based warmup, 45-minute strengthening exercises for the trunk and upper and lower limbs, and finally a 5-minute cool-down with light exercises. Based on the tolerance of each patient, exercise intensity (holding time and number of repetitions) was gradually increased |
| **Rabin, 2014** | SMT | Lumbar Stabilization Exercises program was largely based on the program described by Hicks. Exercises were ordered by their level of difficulty, and patients progressed from one exercise to the next after satisfying specific predetermined criteria. | education | active control | manual therapy ntervention included several thrust and nonthrust manipulative techniques directed at the lumbar spine that have been used previously with some degree of success in various groups with LBP. In addition, manual stretching of several hip and thigh muscles was performed, as flexibility of the lower extremity. inally, active range-of-motion and stretching exercises were added to the program |
| **Rasmussen-Barr, 2009** | SMT | low load endurance exercise to address the stabilizing muscles, after the protocol described by Richardson. The PT individually supervised and used clinical judgment in the progression of the graded stabilizing exercises. First, the subjects were informed of how the stabilizing muscles act, as hypothesized, in healthy people and in those with LBP. he progression of the exercises was based on the patients’ pain level and observed movement control and quality. | none | advice and usual care | were informed of the benefits of daily walks as physical activity. They were instructed to take a 30-minute walk every day. They were also given general home exercises but with no follow-up instructions. |
| **Salas, 2019** | mixed | multidimensional treatment received exercises focused on muscle flexibility and joint range of motion, muscle strengthening, and postural control. This was achieved by a variety of techniques, including specific exercises designed to address muscle tightness and weakness, MTrP release, MET, PNF stretching, area-specific muscle contraction exercises, and neural gliding techniques that are used to reduce and correct muscle tightnes. A multidimensional individually tailored approach was taken to address each participant’s unique problem. | none | active control | traditional senior group exercise therapy treatment included both seated and standing activities focused on improving strength, flexibility, balance, and cardiovascular health. Class exercises were inclusive of daily-living tasks, such as: ascending and descending stairs; chair stands; chairsupported squats; and seated aerobics. |
| Sandal, 2021 | mixed | selfBACK app provided weekly recommendations for physical activity, strength and flexibility exercises, and daily educational messages. Self-management recommendations were tailored to participant characteristics and symptoms. The exercise content consisted of a bank of 70 exercises organized in 6 targets: 1) flexibility exercises, 2) pain relieving exercises in addition to strength exercises for 3) back extensors, 4) gluteal and hip muscles, 5) abdominal muscles, and 6) core muscles. It is important to note that the individual user could adjust their time available for exercise in the tailoring session and consequently, the exercise dose performed would differ in both content, volume and intensity across the intervention group. | intervention group: education | usual care and advice | Usual care included advice or treatment offered to participants by their clinician. |
| **Saner, 2015** | mixed | Movement control treatment consisted of active exercises addressing the pain-provoking postures and control of the impaired movement(s). Exercises aimed specifically at local lumbar stabilising muscles, or treatment according to behavioural classification. Load, frequency and velocity can gradually be increased once the movement is retrained. | none | active control | General exercise treatment aimed to improve the muscular strength of the lumbar and pelvic region and legs. In a standardised programme, as described in a study manual, all relevant muscle groups were addressed in each treatment. Start load and progression were assessed individually and followed a submaximal training protocol, according to the guidelines of the American College of Sports Medicine |
| **Schaller, 2016** | **back school** | Movement Coaching was designed as a multicomponent approach and comprised of three different components: face-to-face contact (small group intervention, twice during inpatient rehabilitation), a tailored telephone aftercare (8 weeks and 12 weeks after rehabilitation) and an Internet-based aftercare | coaching and education | passive control | low intensity intervention merely comprising of two general presentations on physical activity during inpatient rehabilitation which could be downloaded from a homepage during aftercare. |
| **Soukup, 1999** | mixed | Mensendieck exercise program focused on basic pelvic, hip, back and am´bdominal movements in daily life. Variation in regard to exercise type was emphasized. Educational elements of the intervention. Exercise regimen was notb standardised. | educational component in the intervention group | advice and usual care | Written and oral informatuiin about the Mensendieck appraoch as a secondary prevention program. Hiever they did not attend any of the practical training sessions |
| **Suh, 2019** | SMT | I1: Invidualized graded lumbar stabilisation exercise consisted of 2 parts: stretching exercises and stabilisation. We gradually increased the degree of instability until the most unstable posture was achieved. At the beginning, participants were placed into a level with moderate difficulty. I2: Invidualized graded lumbar stabilisation exercise with additional walking exercise | education session was performed at the clinic by a trained physical therapist at the first visit. Moreover, a printed pamphlet with instructions on how to perform the exercises was given to each patient. | all active controls | C1: walking exercise on flat ground with abdominal bracing for 30 minutes C2: flexibility exercise received stretching exercise for the abdominal muscle, quadriceps, hamstring, tensor fascia lata, piriformis muscle, and quadratus lumborum muscles |
| **Thanawat, 2017** | mixed | Transtheoretical Model of behavioral change (TTM) on back muscle endurance, physical function and pain. participants attended an initial baseline assessment. After that, an 8-week intervention program including health education and exercises were administered to all participants. However, strategies used for providing the intervention to the participants in each sub-group were different. In addition Seven home-based exercises for individuals with LBP ) were recommended. The exercises were progressed by increasing exercise sets or advancing to a more difficult program. | health education | active control | conventional health education and exercise training utilizing a slide presentation and an illustrated booklet. In addition home exercise |
| **Tsauo, 2009** | mixed | individualised functional training programme aimed to decrease pain in the back, improve the patients’ endurance, strength, flexibility, general fitness and activity capacity. It consisted of a warm-up exercise (jogging or walking), a strengthening exercise, work/activity simulation training and fitness and endurance training (such as use of a stationary bicycle, stepping or trotting exercise). A strengthening programme focussed on trunk stabilisation training for the superficial and deep trunk muscles and the extremities. | none | **advice and usual care** | might maintain their current rehabilitation programme |
| **Van Baal, 2020** | SMT | sensorimotor treatment was adapted to the specific symptoms of the patients. Therefore, it was focussed on the physical training component with sensory (localisation and graphesthesia training) and motor (laterality recognition and motor control) retraining. Both the sensory and the sensorimotor training components were offered at three difficulty levels with several options for variation and progression in each level. The therapist selected the exercises and tailored the dose based on the previous and actual performance of each participant. | none | active control | General exercise programm consists of four basic exercises (quadripedal position/prone bridging, rowing in a standing position, standing balance/knee bend and side support) with 12 levels of difficulty. The level was selected based on a patient’s performance |
| **Van Dillen, 2016** | SMT | classification-specific (MSI) treatment included 3 components: education, exercise, and training to modify how functional activities were performed, hereinafter referred to as performance training. Progression was based on the participant’s ability to perform the appropriate number of repetitions of an item independently. The primary goals of training were to teach each participant to (a) move the lumbar spine later and reduce the amount of lumbar spine movement in the directions related to the participant’s classification, (b) increase use of other joints such as the hips and knees, and (3) avoid end-range positioning of the lumbar spine in the directions related to the participant’s classification | none | active control | treatment conditions included 3 components: education, exercise, and training. Analogous generic items were used to select treatment items.The primary goal of training was to teach the participant to maintain the “normal spinal curves” during performance of activities and assumption of postures |
| **Vasseljen, 2012** | SMT | core stability exercise was individualized according to protocols focusing on isolated activation of TrA during the abdominal drawingin maneuver (ADIM). When isolated TrA activation was achieved, pelvic floor and multifidus muscle co-contractions were included and the ADIM progressed to sitting and standing. The exercise program lasted 40 minutes and was carried out in a physiotherapy clinic. Written instruction to carry out the ADIM exercise at home was provided | none | all active controls | C1: sling exercise wasv adjusted to each patient’s ability to keep the lumbar spine stable in neutral position throughout a range of leg/ arm positions and movements. Elastic bands were attached to the pelvis to help the patient maintain a neutral spine position at all times and for exercises to progress without pain. Exercise progression was achieved by gradually reducing the elastic band support and placing the patients in progressively more demanding but pain-free positions to provide forceful activation of deep and superficial trunk muscles. |
|  |  |  |  |  | C2: general exercise eceived general strengthening and stretching exercises as recommended in the treatment of nonspecific LBP. |
| **Verra, 2018** | mixed | Multidimensional Pain Inventory to provide subgroup-specific pain management. Patients received specific exercise interventions, which were tailored to the deficits | relaxation therapy, participation in a pain coping group, information and education about the pathophysiology of pain mechanisms | active control | general pain management. no differentiation of interventions between subgroups was done. all patients in the control group participated in state-of-the-art progressive resistance training and stretching exercises according to the recommendations of hayden |
| **Vibe Fersum, 2013** | mixed | classification-based cognitive functional therapy has four main components: (1) a cognitive component, for each patient, their vicious cycle of pain was outlined in a diagram based on their findings from the examination; (2) specific movement exercises designed to normalize maladaptive movement behaviours as directed by the movement classification; (3) targeted functional integration of activities in their daily life, reported to be avoided or provocative by the patient; and (4) a physical activity programme tailored to the movement classification | intervention group with cognitive behavioural management approach, treated with joint mobilization or manipulation techniques applied to the spine or pelvis consistent with best current manual therapy practice. Home exercise. | active control | traditional manual therapy and exercise. |
| **Von Korff, 2005** | back school | The intervention included four in-person visits. An initial 90-min visit with a psychologist: identified and addressed patient fears about back pain; discussed the relationship between resuming normal activities and quality of life; set an activity or exercise goal to enhance quality of life; and developed an action plan to achieve the goal. The second 60-min visit, with a physical therapist, took place 7–10 days later. The physical therapist conducted a standardized mechanical examination of the back, discussed unresolved patient concerns identified in the initial visit, taught stretches and exercises relevant to the action plan, and offered guidance in overcoming barriers the patient had encountered in carrying out the action plan. A third visit (30 min) with a physical therapist occurred about 10 days later. This visit focused on the action plan and exercises relevant to the action plan. After a 2 week interval, a fourth visit (30 min) with the psychologist reviewed progress, encouraged use of relaxation, and developed plans for sustaining progress, managing flare-ups and resuming activities when a flare-up occurred. | contains components of CBT | advice and usual care | received care as usual, whose content is highly variable across patients. Usual care often included use of prescription and non-prescription pain medications, infrequent primary care visits for back pain, and use of ancillary services such as physical therapy by a minority of patients. |
| **Wajswelner, 2012** | pilates | received a tailor-made, directionspecific exercise program prescribed by a physiotherapist based on history, aggravating factors, and physical examination. The clinical Pilâtes exercise program was a series of exercises performed on the reformer and trapeze equipment. The equipment both supports the patient and guides the direction and type of movement required for the prescribed exercises. The exercises were designed to work the patient in a specific direction, for example, flexion, extension, neutral, or to the left or right side. | none | active control | were taught a standardized generic set of exercises traditionally used by physiotherapists for the management of CLBP. These exercises were chosen via consensus of seven musculoskeletal physiotherapists with expertise in exercise prescription as well asfi-omprevious studies. Additional home exwercise |
| **Wälti, 2015** | SMT | Multimodal treatment group MMT included: 1. Education on the neurophysiology of pain; 2. Sensory retraining; and 3. Motor retraining. Treatment was aimed at reducing pain and disability and, potentially, addressing associated abnormal cortical processing. Patients from both groups had personalized access to a web-based home training interface | Intervention group: Patient education on the neurophysiology of pain was aimed at the reduction of patients’ perception of pain and disability, a reconsideration of protective behaviour and self-limitation resulting from fear | active control | basic patient education on adequate behaviour when having an exacerbation of LBP: a short period of protection followed, as soon as possible, by a return to normal movement, work and leisure activities. Further sessions addressed signs and symptoms. A maximum of ten minutes of passive applications per session was allowed (such as massage,manual therapy, electrotherapy, mud packs). |

Appendix D. References of Included and Excluded Studies

D1. Included Studies

D2. Excluded studies at full-text stage

10. Chan AFJHASLRMSSPTDMHR, Taylor N. 1 year results of a randomised controlled trial comparing subgroup specific physiotherapy against advice for people with low back disorders. Physiotherapy (united kingdom) 2015;101:eS207‐eS08.

27. Hallidy MPECHHM, Ferreira P. Short-term effects of mckenzie vs. motor control approach for patients with chronic lowback pain and a derangement classification. Physiotherapy (united kingdom) 2015;101:eS506‐eS07.

Appendix E. Risk of Bias

E1. Risk of Bias Assessment

Supplementary Figure 1. Aggregate Cochrane Risk-of-bias appraisal results

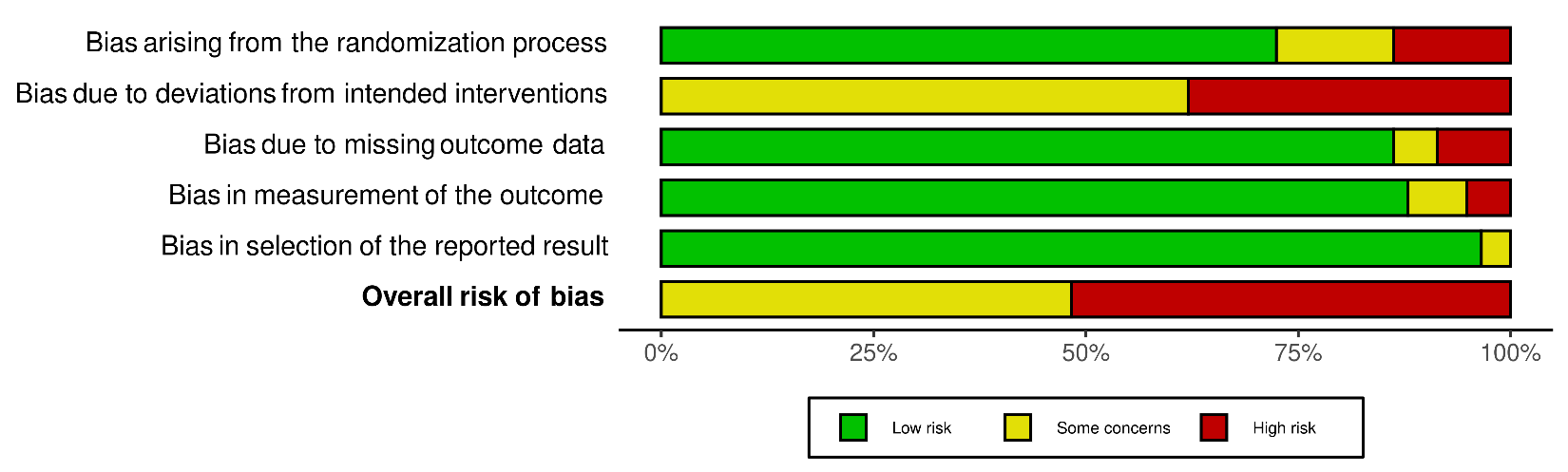

Supplementary Table 1. Cochrane Risk-of-bias global judgement

| Study | Selection bias: randomization | Performance bias: deviation from intended intervention | Attrition bias: Incomplete outcome data | Detection bias: Blinding of outcome assessment* | Reporting bias | Overall |
| --- | --- | --- | --- | --- | --- | --- |
| Aasa 2015 | low | some concerns | low | low | low | Some concerns |
| Andersen 2016 | low | high | low | low | low | high |
| Apeldoorn 2012 | low | some concerns | high | low | low | high |
| Azevedo 2018 | low | high | low | low | low | high |
| Brady 2018 | low | some concerns | low | low | low | some concerns |
| Cairns 2006 | high | high | low | low | low | high |
| Cherkin 1998 | low | high | low | low | low | high |
| Cuesta-Vargas 2011 | high | some concerns | low | low | low | high |
| Descarreaux 2002 | high | high | high | high | some concerns | high |
| Diaz-Arribas 2015 | low | high | low | low | low | high |
| Ford 2016 | low | some concerns | low | low | low | some concerns |
| Garcia 2018 | low | some concerns | low | low | low | some concerns |
| Geisser 2005 | some concerns | some concerns | low | some concerns | low | high |
| Godfrey 2020 | low | some concerns | low | low | low | some concerns |
| Goertz 2017 | low | high | low | low | low | high |
| Gudavalli 2006 | low | some concerns | low | low | low | some concerns |
| Hansen 1993 | high | some concerns | low | some concerns | low | high |
| Heinrich 2009 | some concerns | some concerns | low | low | low | some concerns |
| Henry 2014 | low | some concerns | low | low | low | some concerns |
| Highland 2018 | low | some concerns | low | low | low | some concerns |
| Hill 2011 | low | some concerns | low | low | low | some concerns |
| Hill 2020 | low | high | low | low | low | high |
| Hueppe, 2019 | Low | Some concerns | Low | Low | Low | Some concerns |
| Hurley 2015 | low | some concerns | low | low | low | some concerns |
| Jay 2015 | some concerns | some concerns | low | low | low | some concerns |
| Jensen 2011 | low | high | low | low | low | high |
| Kim 2020 | low | high | low | low | low | high |
| Lang 2021 | low | some concerns | low | low | low | some concerns |
| Lehtola 2016 | low | some concerns | low | low | low | some concerns |
| Leibetseder 2007 | some concerns | some concerns | low | some concerns | low | high |
| Lomond 2015 | low | high | some concerns | low | low | high |
| Long 2004 | low | high | some concerns | low | low | high |
| Macedo 2012 | low | high | low | low | low | high |
| Magalhães 2018 | low | high | low | low | low | high |
| Michaelson 2016 | low | some concerns | low | low | low | some concerns |
| Moore 2000 | low | some concerns | low | low | low | some concerns |
| O’Keeffe 2020 | low | high | low | low | low | high |
| Paolucci 2011 | low | high | high | low | some concerns | high |
| Petersen 2011 | low | high | some concerns | low | low | high |
| Rabiei 2021 | some concerns | some concerns | low | low | low | high |
| Rabin 2014 | low | some concerns | low | low | low | some concerns |
| Rasmussen-Barr 2009 | some concerns | some concerns | low | low | low | some concerns |
| Salas 2019 | high | high | low | high | low | high |
| Sandal, 2021 | Low | Some concerns | Low | Low | Low | Some concerns |
| Saner 2015 | low | some concerns | low | low | low | some concerns |
| Schaller 2016 | high | high | low | some concerns | low | high |
| Soukup 1999 | low | some concerns | low | low | low | some concerns |
| Suh 2019 | low | high | low | low | low | high |
| Thanawat 2017 | high | high | low | high | low | high |
| Tsauo 2009 | high | some concerns | low | low | low | some concerns |
| Van Baal 2020 | low | high | low | low | low | high |
| Van Dillen 2016 | some concerns | some concerns | high | low | low | high |
| Vasseljen 2012 | some concerns | some concerns | high | low | low | high |
| Verra 2018 | low | some concerns | low | low | low | some concerns |
| Vibe Fersum 2013 | low | some concerns | low | low | low | some concerns |
| Von Korff 2005 | low | some concerns | low | low | low | some concerns |
| Wajswelner 2012 | low | some concerns | low | low | low | some concerns |
| Wälti 2015 | low | some concerns | low | low | low | some concerns |

E2. Risk of Bias Assessment: Publication Bias

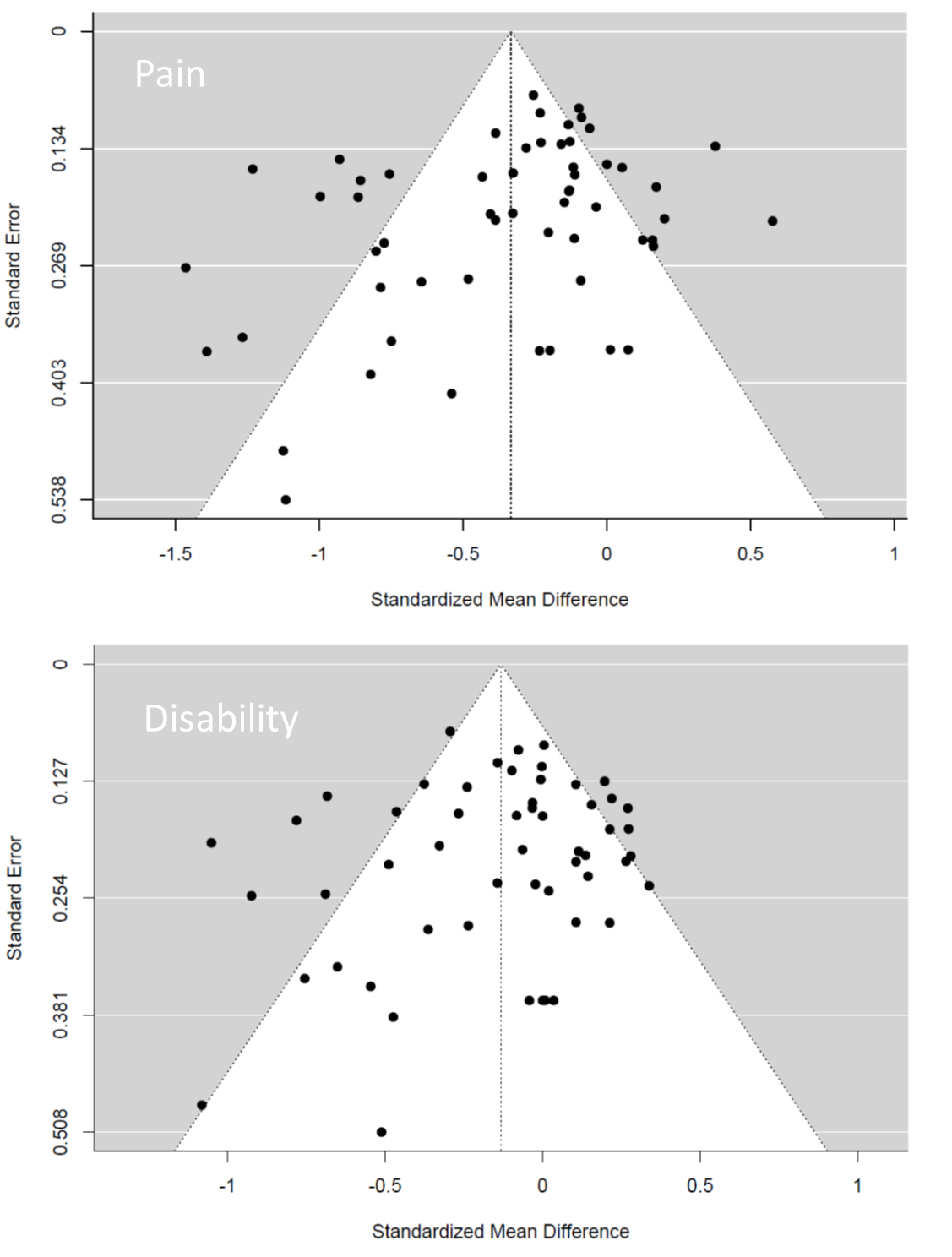

Supplementary Figure 2. Funnel plots for pain (above) and disability (below). The dots represent the individual studies. Standardized mean differences are plotted against standard errors.

Appendix F. Quality of evidence and grading strength of recommendations

**Supplementary Table 3 GRADE assessment of meta-analytic results.** *RCTs have been defined being the basic study design (i.e. type of evidence is high) and downgradings have been performed according to the five domains risk of bias, inconsistency, indirectness imprecision, and publication bias. ^#^The overall risk of publication bias according to the funnel plots (*Supplementary Figure 2*) was supposable for short-term pain outcomes, and outliers (studentised residuals) and overly influent studies (Cook’s distances) were analysed to further assess this domain. The quality of evidence is categorized as follows: High* (⊙⊙⊙⊙)*: further research is very unlikely to change the confidence in the estimate of effect. Moderate* (⊙⊙⊙○)*: further research is likely to have an important impact in the confidence in the estimate of effect. Low* (⊙⊙○○)*: further research is very likely to have an important impact on our confidence in the estimate of effect and is likely to change the estimate. Very Low* (⊙○○○)*: any estimate of effect is very uncertain. FU: Follow-Up, OIS: optimal information size, CI: confidence interval*

| Outcome | Risk of bias | Inconsistency | Indirectness | Imprecision | Publication bias^#^ | Result |
| --- | --- | --- | --- | --- | --- | --- |
| Pain intensity: Personalised exercise therapy is more effective vs. active treatments on short-term FU | Downgrade by one level. Serious limitation due to performance bias | Downgrade by one level. Heterogeneity can not be explained  I^2^ = 73% | No evidence of indirectness | No downgrade. OIS sufficient. The CI does not entail a clinical meaningful result | No downgrading, evidence of bias cannot be excluded. No indication of outliers. No study overly influential | Low (⊙⊙○○) |
| Pain intensity: Personalised exercise therapy is more effective vs. passive treatments on short-term FU | Downgrade by one level. Serious limitation due to performance bias | Downgrade by one level. Heterogeneity can not be explained  I^2^ = 89% | No evidence of indirectness | No downgrade. OIS sufficient. The CI does not entail a clinical meaningful result | Downgrade by one level. Evidence of bias due to potential  funnel plot asymmetry. | Very Low (⊙○○○) |
| Disability: Personalised exercise therapy is more effective vs. active treatments on short-term FU | Downgrade by one level. Serious limitation due to performance bias | Downgrade by one level. Heterogeneity can not be explained  I^2^ = 73% | No evidence of indirectness | No downgrade. OIS sufficient. The CI does not entail a clinical meaningful result | No evidence of bias. No indication of outliers. No study overly influential | Low (⊙⊙○○) |
| Disability: Personalised exercise therapy is more effective vs. passive treatments on short-term FU | Downgrade by one level. Serious limitation due to performance bias | No downgrade. Heterogeneity considered moderate  I^2^ = 54% | No evidence of indirectness | No downgrade. OIS sufficient. The CI does not entail a clinical meaningful result | No evidence of bias. No indication of outliers. No study overly influential | Moderate (⊙⊙⊙○) |
| Pain intensity: Personalised exercise therapy is not more effective vs. active treatments on long-term FU | Downgrade by one level. Serious limitation due to performance bias | No downgrade. Heterogeneity considered low to moderate  I^2^ = 41% | No evidence of indirectness | No downgrade. OIS sufficient. The CI does not entail a clinical meaningful result | No evidence of bias. No indication of outliers. No study overly influential | Moderate (⊙⊙⊙○) |
| Pain intensity: Personalised exercise therapy is more effective vs. passive treatments on long-term FU | Downgrade by one level. Serious limitation due to performance bias | No downgrade. Heterogeneity considered low  I^2^ = 29% | No evidence of indirectness | No downgrade. OIS sufficient. The CI does not entail a clinical meaningful result | No evidence of bias. No indication of outliers. No study overly influential | Moderate (⊙⊙⊙○) |
| Disability: Personalised exercise therapy is not more effective vs. active treatments on long-term FU | Downgrade by one level. Serious limitation due to performance bias | Downgrade by one level. Heterogeneity can not be explained  I^2^ = 69% | No evidence of indirectness | No downgrade. OIS sufficient. The CI does not entail a clinical meaningful result | Downgrade by one level. One study (Vibe Fersum 2013) may be a potential outlier. No study overly influential. | Very Low (⊙○○○) |
| Disability: Personalised exercise therapy is more effective vs. passive treatments on long-term FU | Downgrade by one level. Serious limitation due to performance bias | No downgrade. Heterogeneity considered moderate  I^2^ = 51% | No evidence of indirectness | No downgrade. OIS sufficient. The CI does not entail a clinical meaningful result | No evidence of bias. No indication of outliers. No study overly influential. | Moderate (⊙⊙⊙○) |

Appendix G. Mean differences

**Supplementary Table 4. Mean differences (MD).** LBP = chronic non-specific low back pain

|  | MD pain | | |  | | MD disability | | |  | |
| --- | --- | --- | --- | --- | --- | --- | --- | --- | --- | --- |
|  | individualised | control | | | difference | individualised | | control | | difference |
| Short-term | | |  | | | |  | | | |
| *active* | -2 | -1.45 | | | -0.55 | -1.06 | | -0.838 | | -0.222 |
| *passive* | -1.97 | -1.11 | | | -0.86 | -1.12 | | -0.919 | | -0.201 |
| Long-term | | | | | | | | | | |
| *active* | -1.96 | -1.77 | | | -0.19 | -1.03 | | -0.99 | | -0.04 |
| *passive* | -1.71 | -1.43 | | | -0.28 | -1.26 | | -1.17 | | -0.09 |
| Studies with utmost moderate overall risk of bias | | | | | | | | | | |
|  | -1.89 | -1.44 | | | -0.45 | -1.09 | | -0.884 | | -0.206 |
| Individualised (mixed) exercise interventions without any form of psychological therapy | | | | | | | | | | |
|  | -1.82 | -1.49 | | | -0.33 | -0.84 | | -0.741 | | -0.099 |
| Individualised exercise including psychological interventions | | | | | | | | | | |
| *Overall* | -2.1 | -1.43 | | | -0.67 | -1.18 | | -0.891 | | -0.289 |
| *with CBT* | -2.78 | -1.51 | | | -1.27 | -1.77 | | -0.972 | | -0.798 |
| *with BT* | -0.66 | -0.57 | | | -0.09 | -0.22 | | -0.13 | | -0.09 |
| *other* | -2.03 | -1.46 | | | -0.57 | -1.1 | | -0.92 | | -0.18 |
| Exercise based on individualised motor-control principles | | | | | | | | | | |
|  | -1.87 | -1.21 | | | -0.66 | -1.05 | | -0.799 | | -0.251 |
| Only matched comparator groups (same intervention except individualisation) | | | | | | | | | | |
|  | -1.89 | -1.17 | | | -0.72 | -1.36 | | -0.878 | | -0.482 |
| Studies with true controls | | | | | | | | | | |
|  | -2.26 | -1.57 | | | -0.69 | -1.38 | | -1.29 | | -0.09 |
| Studies with usual care and advice controls | | | | | | | | | | |
|  | -2.06 | -1.02 | | | -1.04 | -1.19 | | -0.885 | | -0.305 |
| Only LBP at inclusion > 12 weeks | | | | | | | | | | |
|  | -1.71 | -0.993 | | | -0.717 | -0.677 | |  | | -0.542 |

Appendix H. Main analysis long-term follow-up

### H1. Individualized exercise versus other active treatments on pain intensity

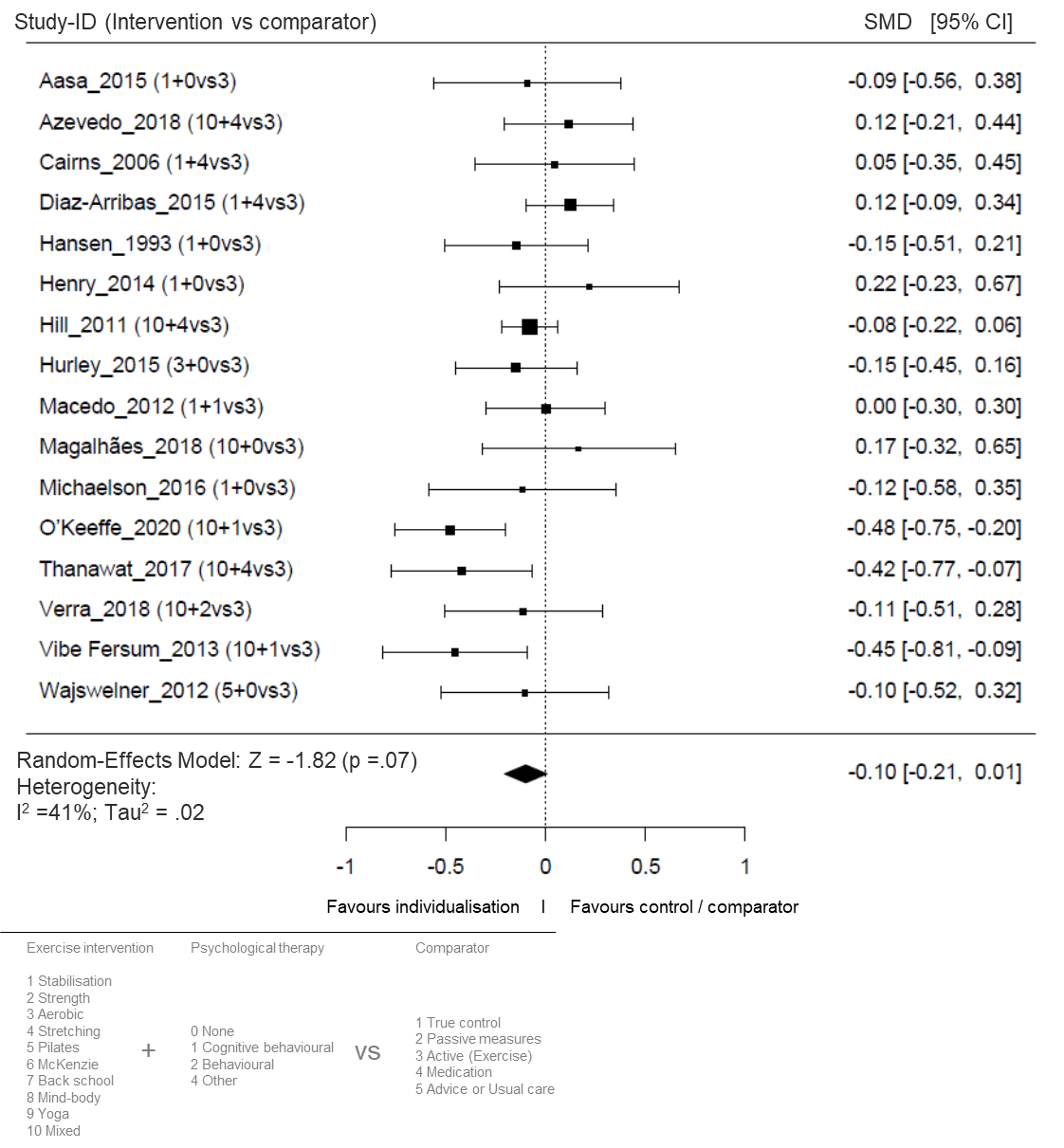

Supplementary Figure 3. Forest plot for the effect sizes for the long-term follow-up (closest to 12 months after randomization) of individualized exercise versus other active treatments on pain intensity. The plot depicts model fit, individual study and pooled effect size estimates (standardized mean differences and corresponding 95% confidence intervals). The size of the boxes corresponds to the respective studies’ (inverse variance) weighting. SMD: standardized mean differences; CI: confidence interval; vs: versus.

### H2. Individualized exercise versus passive treatments on pain intensity

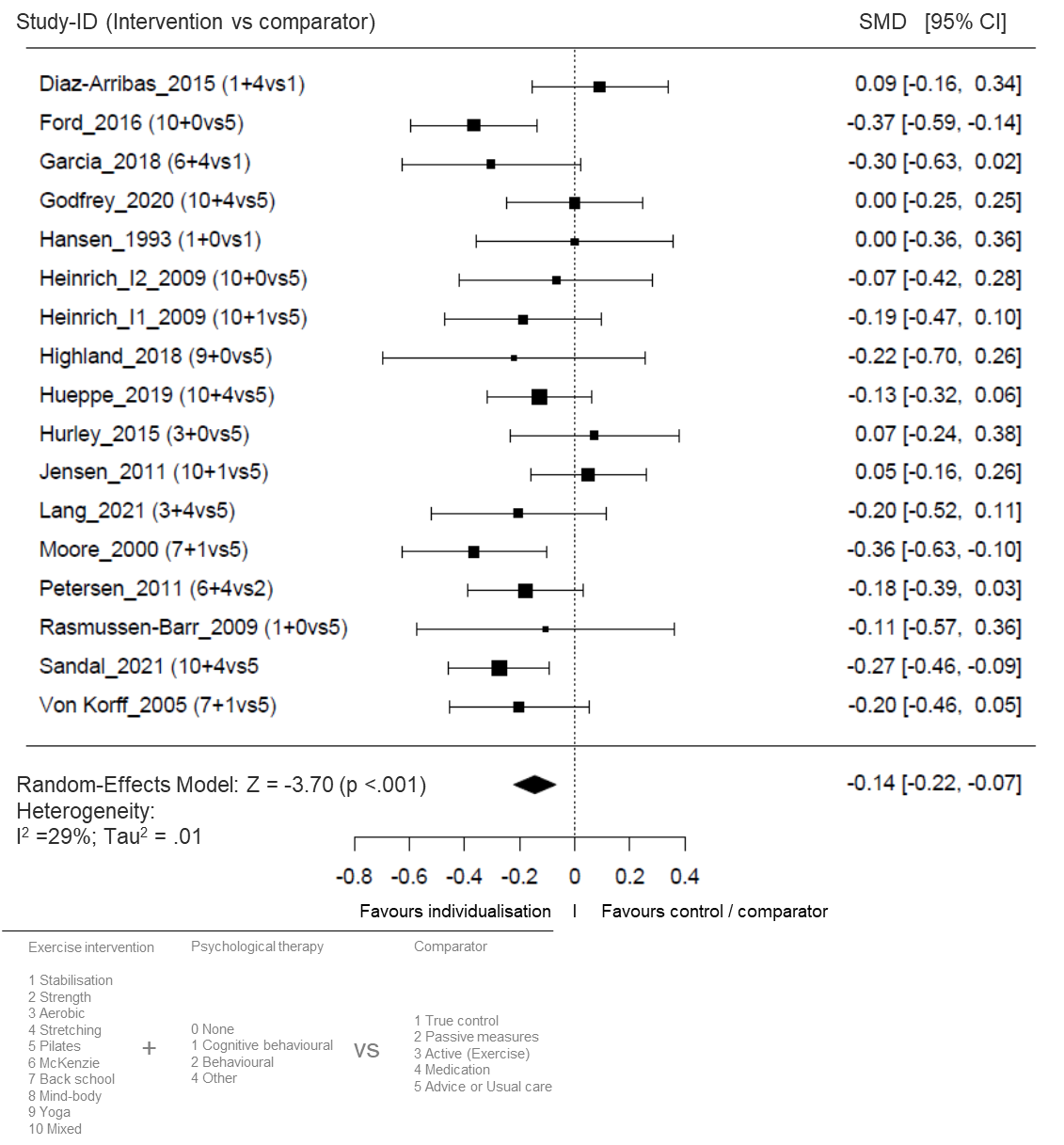
Supplementary Figure 4. Forest plot for the effect sizes for the sizes for the long-term follow-up of individualized exercise versus passive treatments or true control on pain intensity. The plot depicts model fit, individual study and pooled effect size estimates (standardized mean differences and corresponding 95% confidence intervals). The size of the boxes corresponds to the respective studies’ (inverse variance) weighting. SMD: standardized mean differences; CI: confidence interval; vs: versus.

### H3. Individualized exercise versus other active treatments on disability

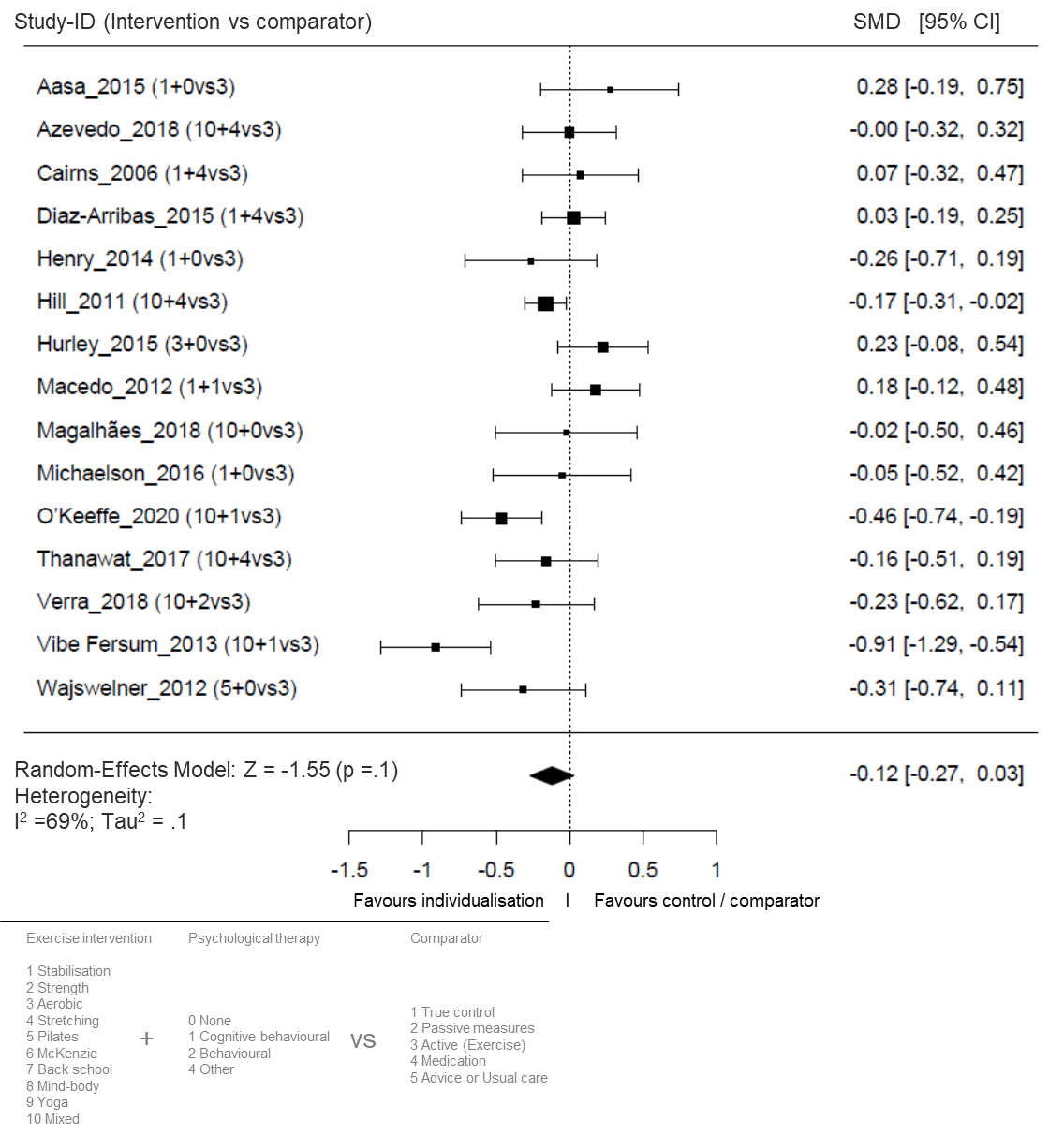
Supplementary Figure 5. Forest plot for the effect sizes for the long-term follow-up (closest to 12 months after randomization) of individualized exercise versus other active treatments on disability. The plot depicts model fit, individual study and pooled effect size estimates (standardized mean differences and corresponding 95% confidence intervals). The size of the boxes corresponds to the respective studies’ (inverse variance) weighting. SMD: standardized mean differences; CI: confidence interval; vs: versus

### H4. Individualized exercise versus passive treatments on disability

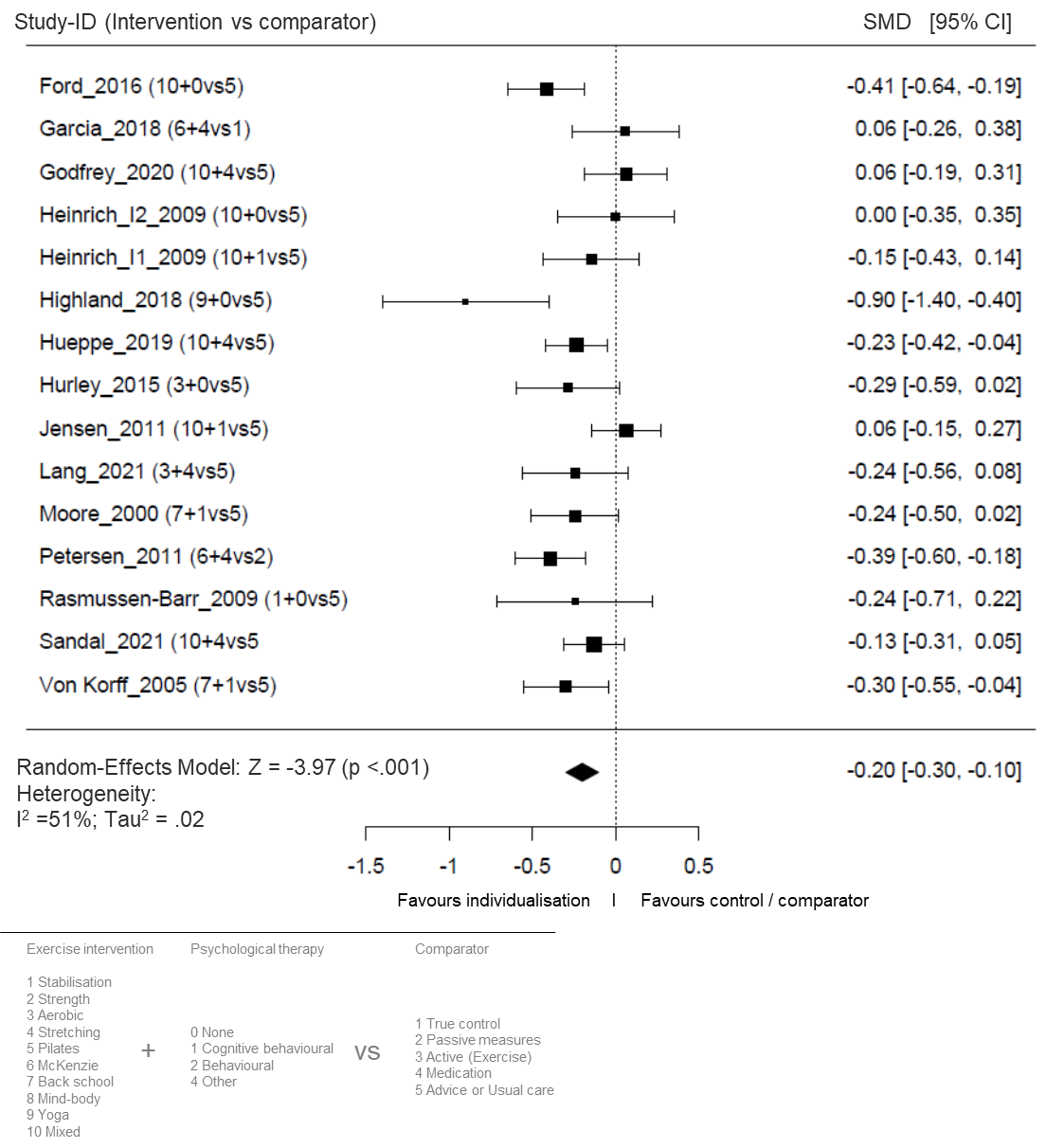
Supplementary Figure 6. Forest plot for the effect sizes for the sizes for the long-term follow-up (12 months) of individualized exercise versus passive treatments or true control on disability. The plot depicts model fit, individual study and pooled effect size estimates (standardized mean differences and corresponding 95% confidence intervals). The size of the boxes corresponds to the respective studies’ (inverse variance) weighting. SMD: standardized mean differences; CI: confidence interval; vs: versus.

Appendix I. Sensitivity analyses – short-term effects in comparison to other active treatments (exercises)

### I1. Studies including only patients with duration of LBP > 12 weeks

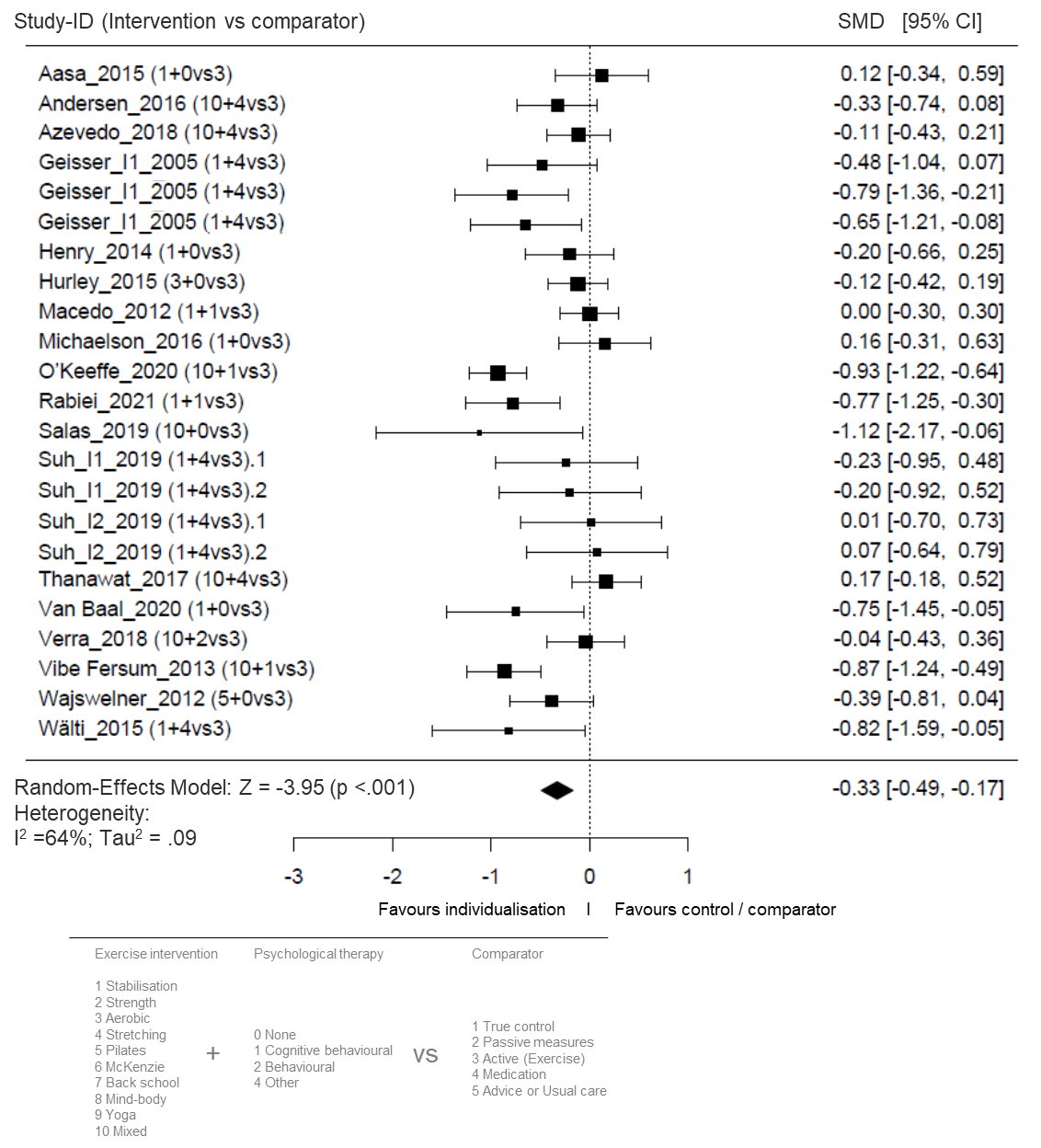
Supplementary Figure 7. Forest plot for the effect sizes for the sizes for the short-term follow-up (12 weeks) of individualized exercise versus active comparators on pain intensity. The plot depicts model fit, individual study and pooled effect size estimates (standardized mean differences and corresponding 95% confidence intervals). The size of the boxes corresponds to the respective studies’ (inverse variance) weighting. SMD: standardized mean differences; CI: confidence interval; vs: versus.

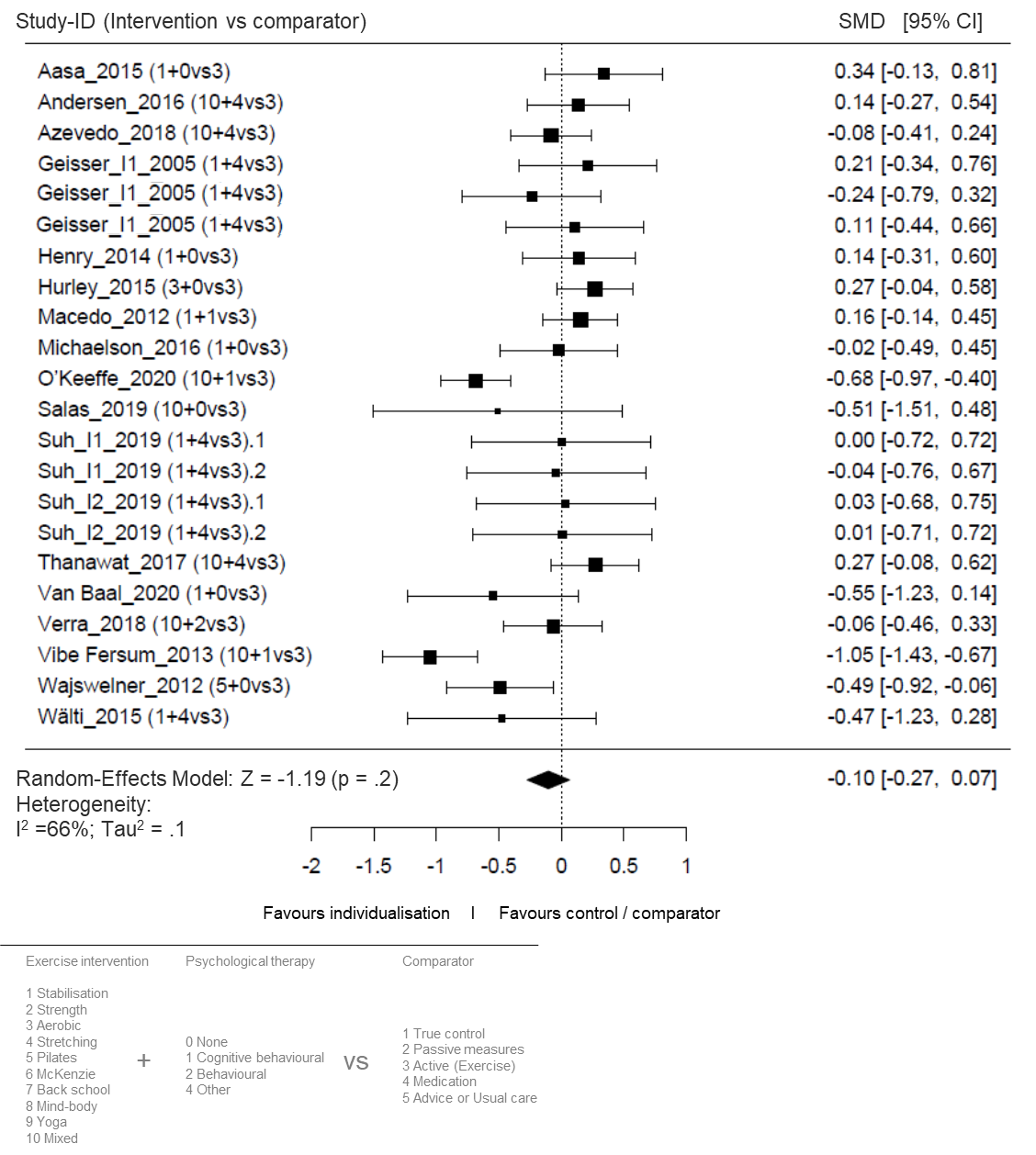
Supplementary Figure 8. Forest plot for the effect sizes for the sizes for the short-term follow-up (12 weeks) of individualized exercise versus active comparators on disability. The plot depicts model fit, individual study and pooled effect size estimates (standardized mean differences and corresponding 95% confidence intervals). The size of the boxes corresponds to the respective studies’ (inverse variance) weighting. SMD: standardized mean differences; CI: confidence interval; vs: versus.

### I2. Studies including only studies with outcomes with moderate or low risk of bias

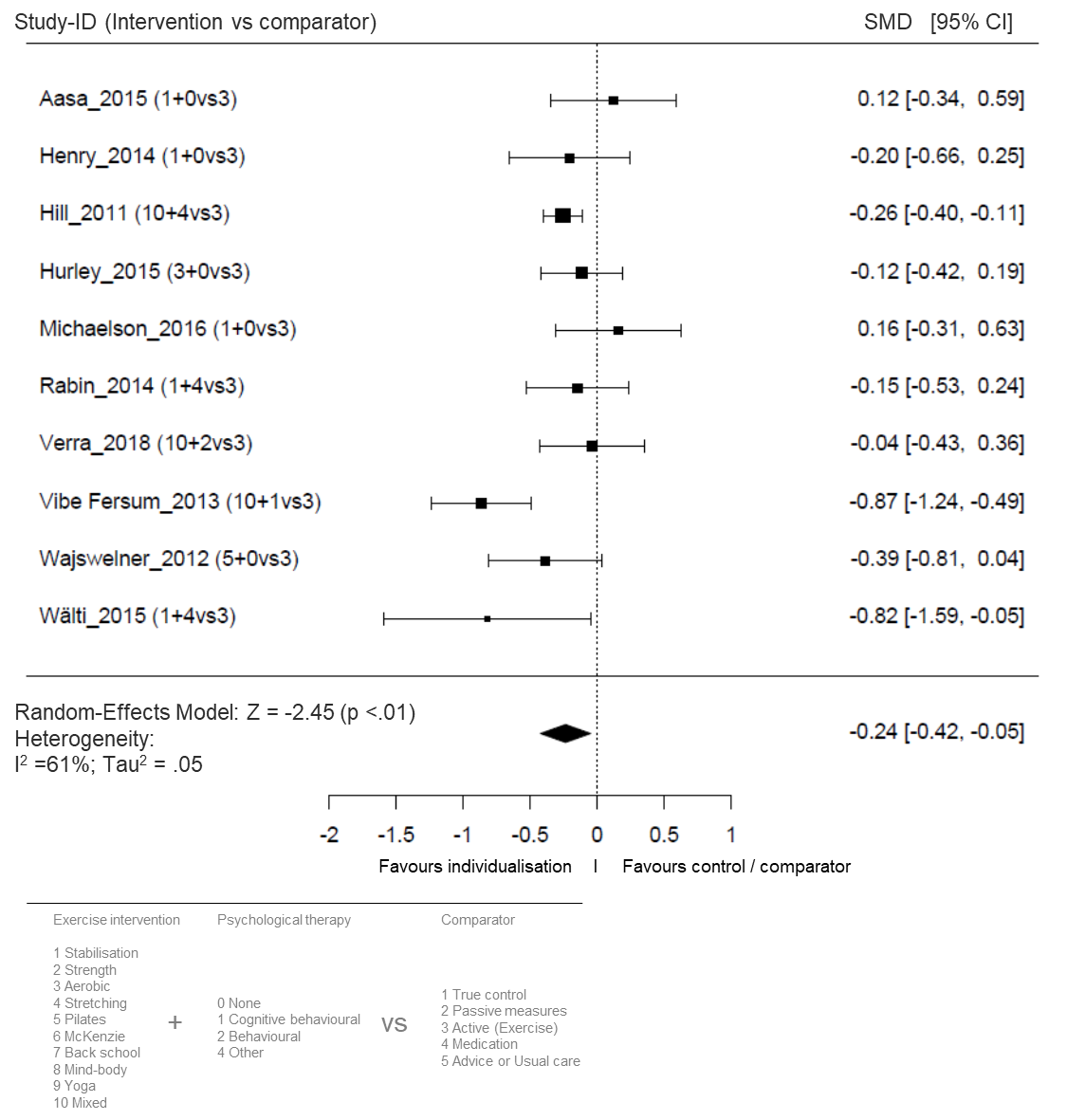
Supplementary Figure 9. Forest plot for the effect sizes for the sizes for the short-term follow-up (12 weeks) of individualized exercise versus active comparators on pain intensity. The plot depicts model fit, individual study and pooled effect size estimates (standardized mean differences and corresponding 95% confidence intervals). The size of the boxes corresponds to the respective studies’ (inverse variance) weighting. SMD: standardized mean differences; CI: confidence interval; vs: versus.

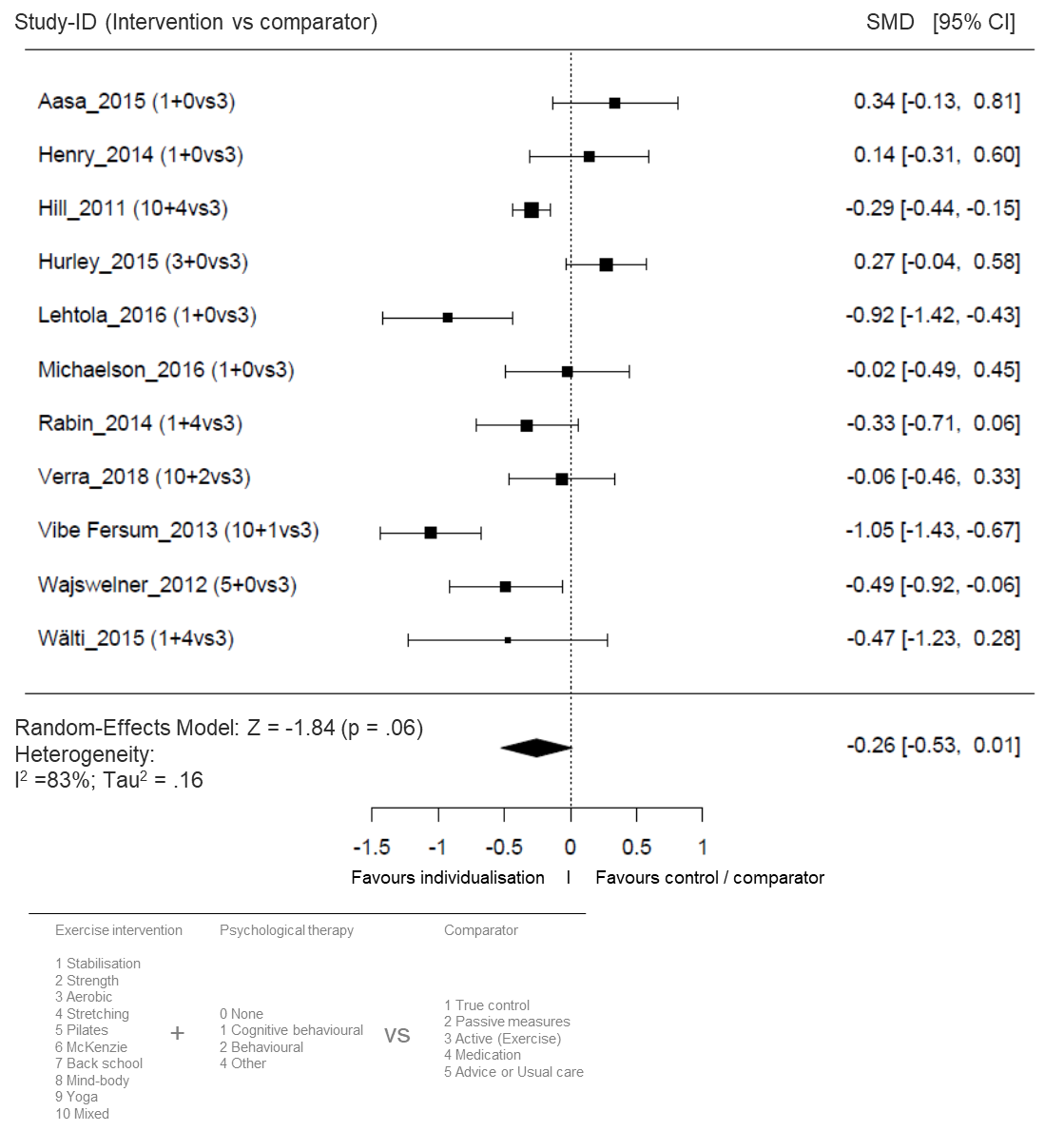
Supplementary Figure 10. Forest plot for the effect sizes for the sizes for the short-term follow-up (12 weeks of individualized exercise versus active comparators on disability. The plot depicts model fit, individual study and pooled effect size estimates (standardized mean differences and corresponding 95% confidence intervals). The size of the boxes corresponds to the respective studies’ (inverse variance) weighting. SMD: standardized mean differences; CI: confidence interval; vs: versus.

### I3. Studies including only patients with specific treatments: exercise only (no psychological intervention) versus other active treatments

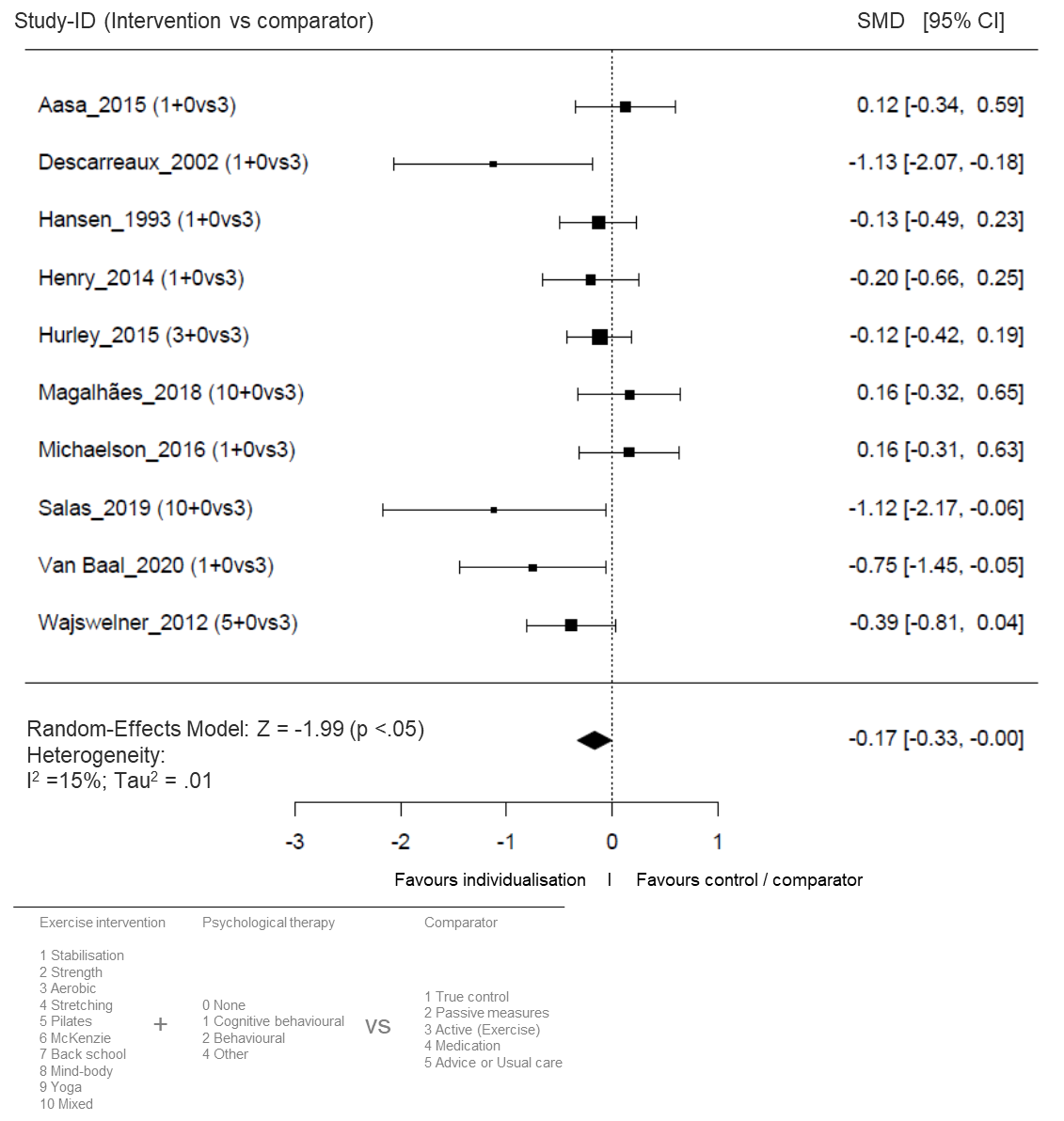
Supplementary Figure 11. Forest plot for the effect sizes for the sizes for the short-term follow–up (12 weeks) of individualized exercise versus active comparators on pain intensity. The plot depicts model fit, individual study and pooled effect size estimates (standardized mean differences and corresponding 95% confidence intervals). The size of the boxes corresponds to the respective studies’ (inverse variance) weighting. SMD: standardized mean differences; CI: confidence interval; vs: versus.

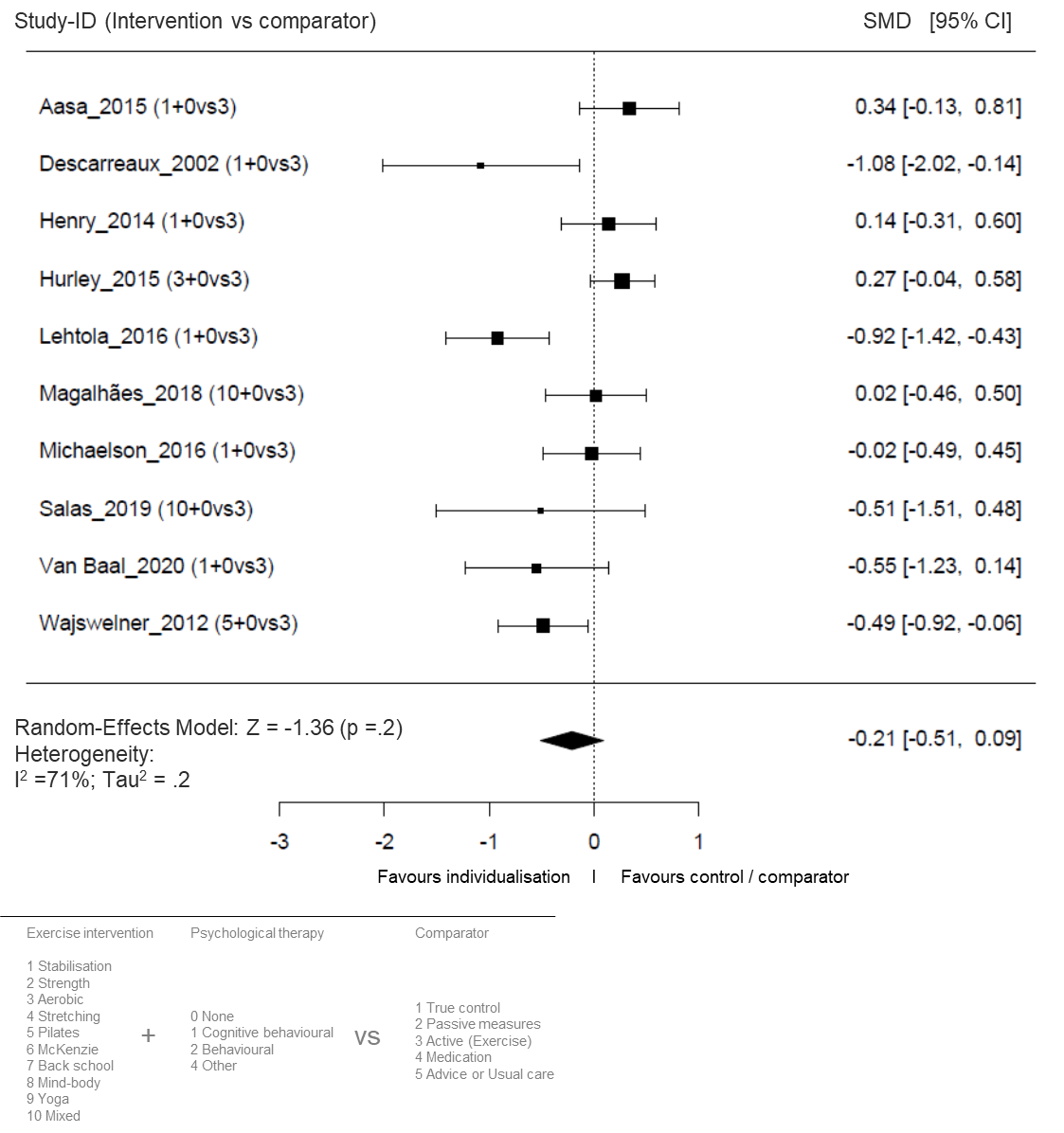
Supplementary Figure 12. Forest plot for the effect sizes for the sizes for the short-term follow-up (12 weeks) of individualized exercise versus active comparators on disability. The plot depicts model fit, individual study and pooled effect size estimates (standardized mean differences and corresponding 95% confidence intervals). The size of the boxes corresponds to the respective studies’ (inverse variance) weighting. SMD: standardized mean differences; CI: confidence interval; vs: versus.

### I4. Studies including only patients with specific treatments: only exercises with psychologic interventions versus other active treatments

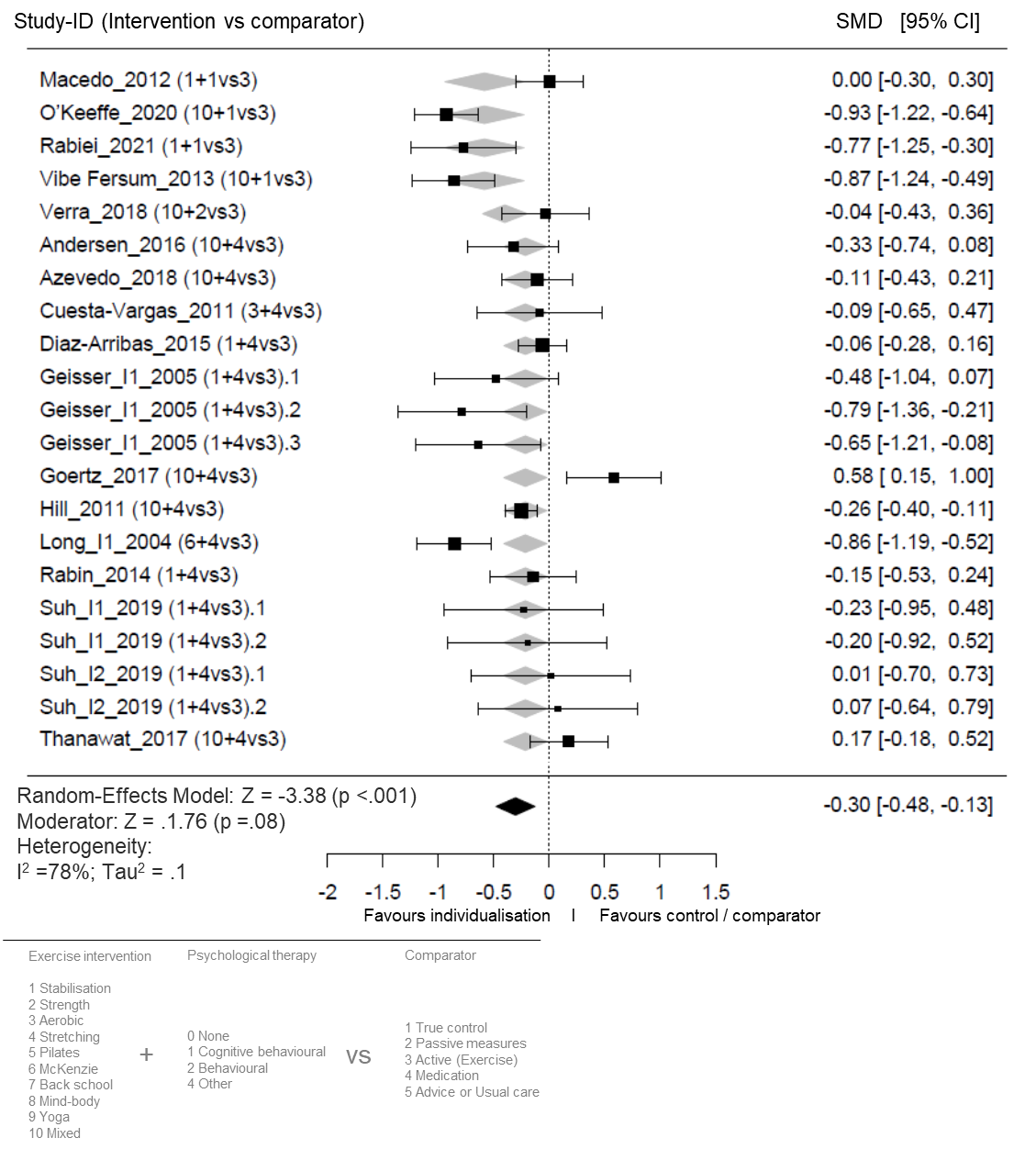
Supplementary Figure 13. Forest plot for the effect sizes for the sizes for the short-term follow-up (12 weeks) of individualized exercise versus active comparators on pain intensity. The plot depicts model fit, individual study and pooled effect size estimates (standardized mean differences and corresponding 95% confidence intervals). The size of the boxes corresponds to the respective studies’ (inverse variance) weighting. SMD: standardized mean differences; CI: confidence interval; vs: versus.

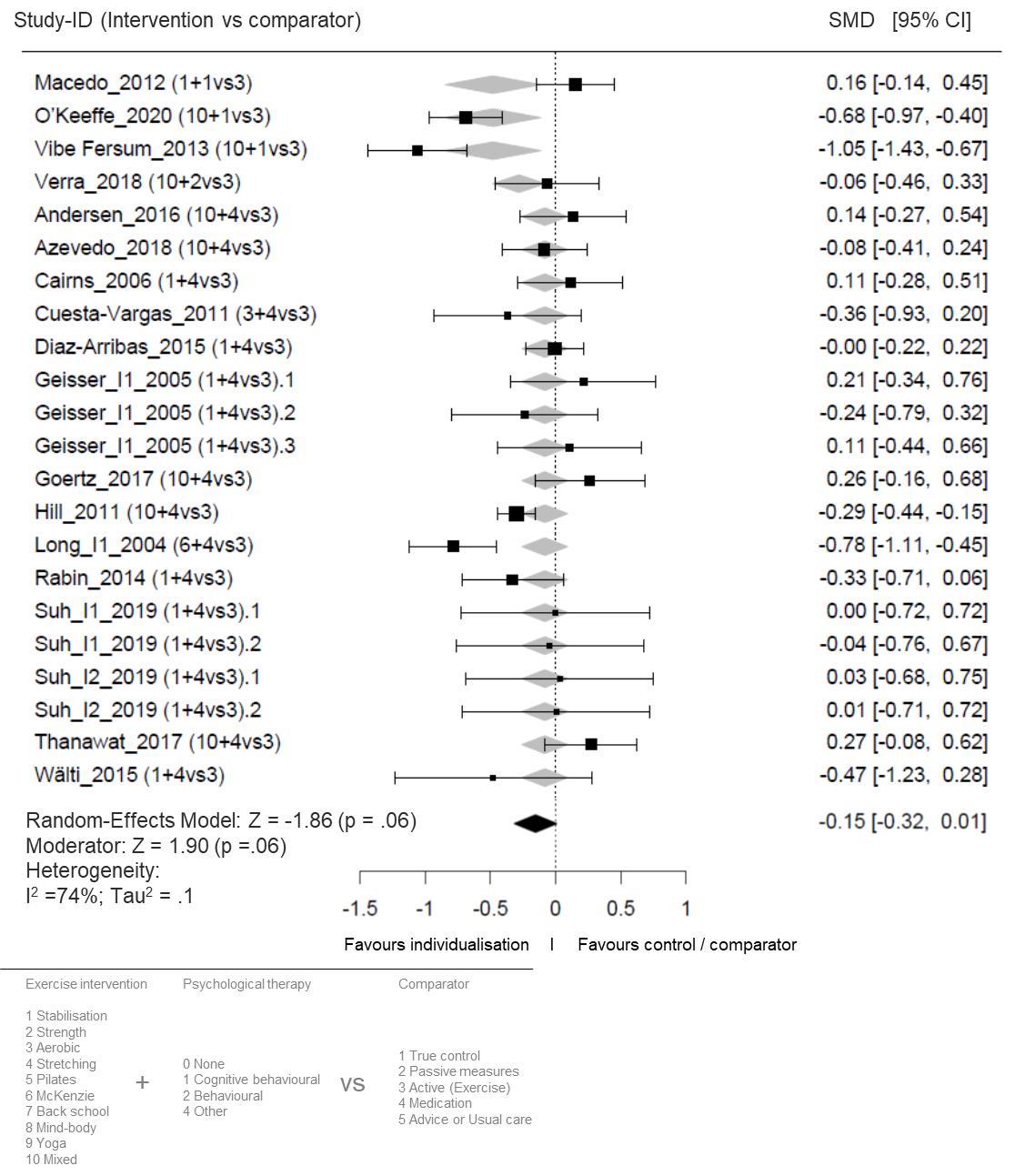
Supplementary Figure 14. Forest plot for the effect sizes for the sizes for the short-term follow-up (12 weeks) of individualized exercise versus active comparators on disability. The plot depicts model fit, individual study and pooled effect size estimates (standardized mean differences and corresponding 95% confidence intervals). The size of the boxes corresponds to the respective studies’ (inverse variance) weighting. SMD: standardized mean differences; CI: confidence interval; vs: versus.

I5. Studies including only patients with specific treatments: only effective exercises according to Hayden 2021 versus other active treatments
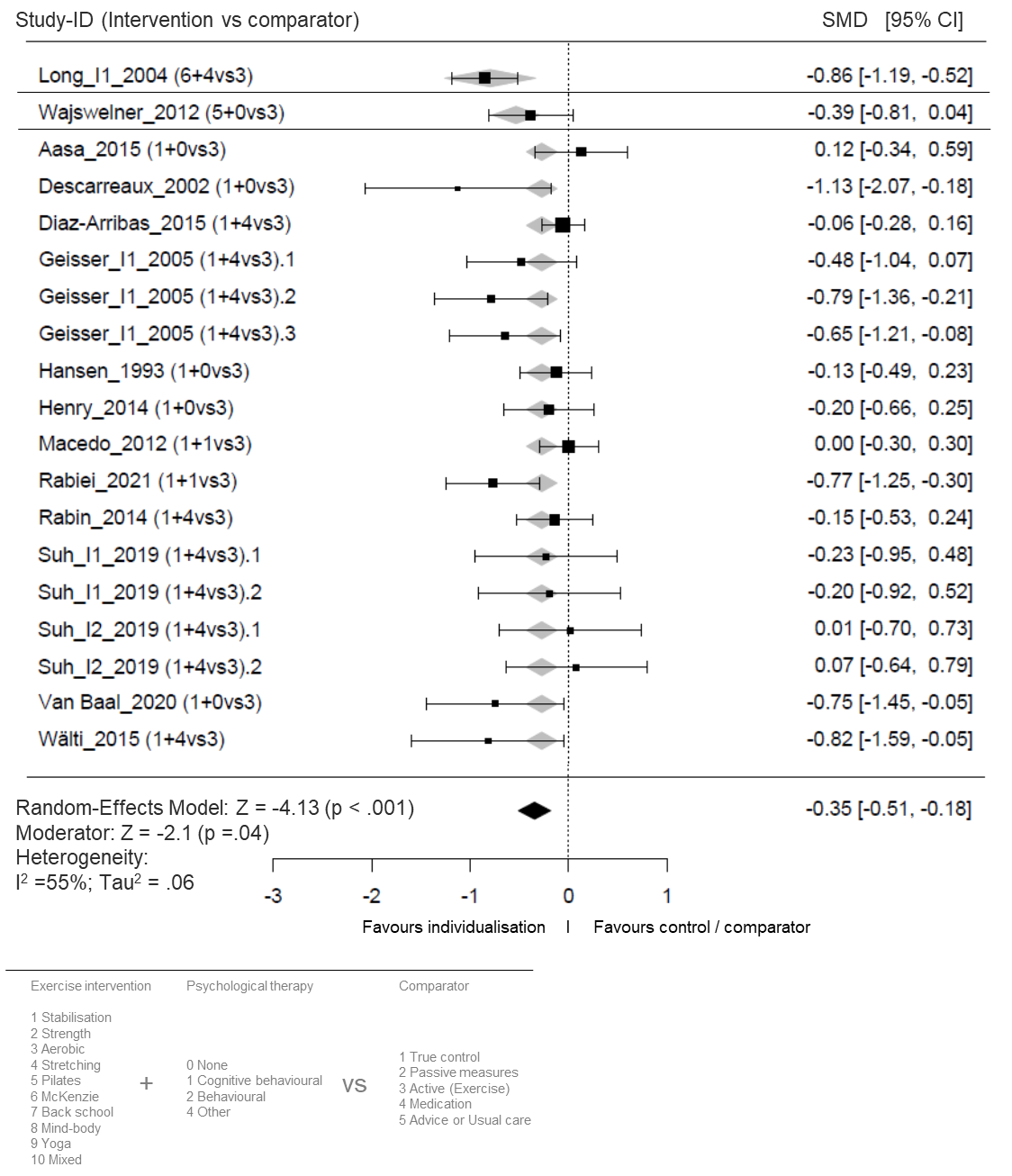
Supplementary Figure 15. Forest plot for the effect sizes for the sizes for the short-term follow-up (12 weeks) of very effective individualized exercise versus active comparators on pain intensity. The plot depicts model fit, individual study and pooled effect size estimates (standardized mean differences and corresponding 95% confidence intervals). The size of the boxes corresponds to the respective studies’ (inverse variance) weighting. SMD: standardized mean differences; CI: confidence interval; vs: versus.

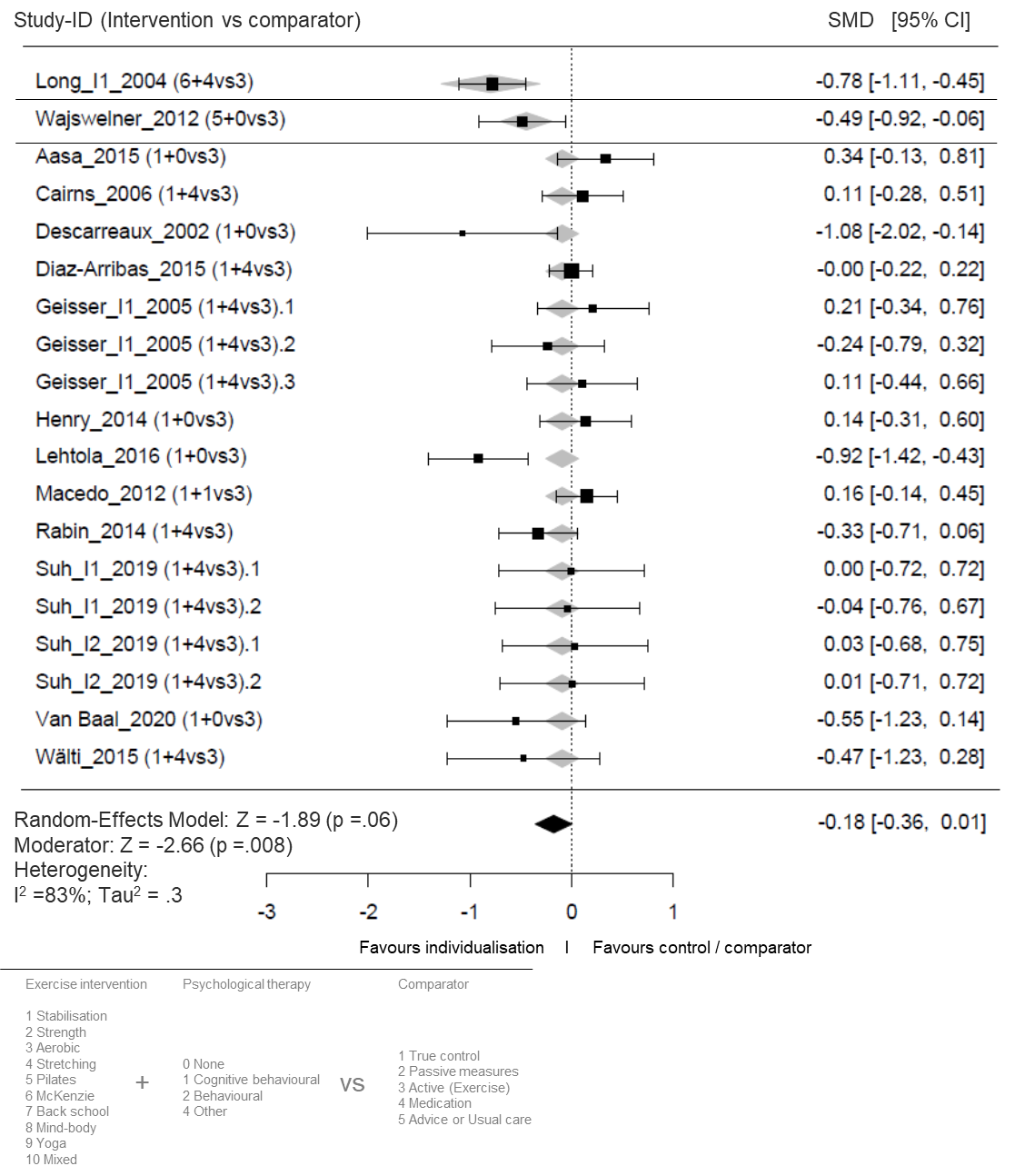
Supplementary Figure 16. Forest plot for the effect sizes for the sizes for the short-term follow-up (12 weeks) of very effective individualized exercise versus active comparators on disability. The plot depicts model fit, individual study and pooled effect size estimates (standardized mean differences and corresponding 95% confidence intervals). The size of the boxes corresponds to the respective studies’ (inverse variance) weighting. SMD: standardized mean differences; CI: confidence interval; vs: versus.

### I6. Studies including only patients with specific controls: only matched comparator groups (same intervention except individualisation)

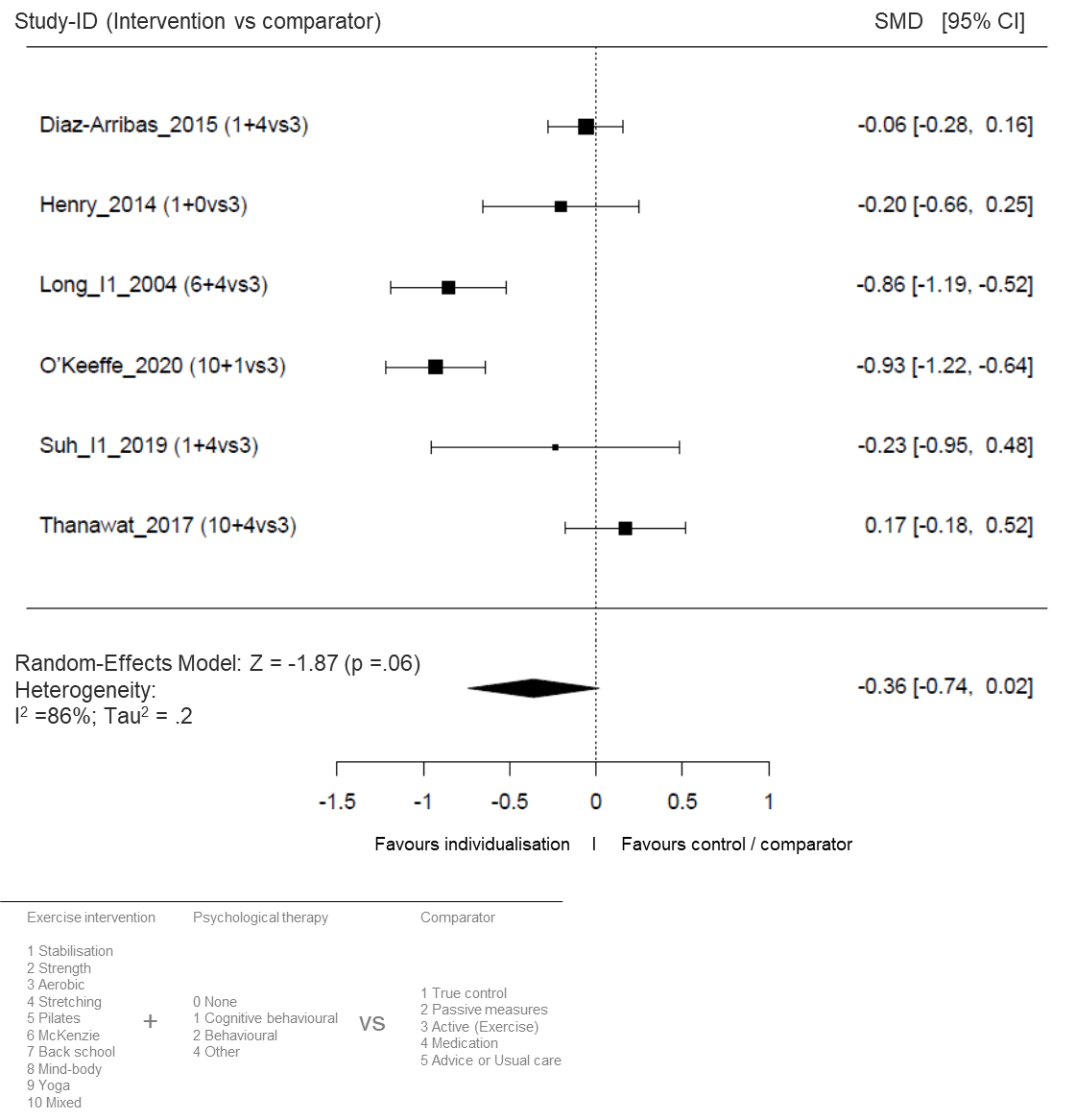
Supplementary Figure 17. Forest plot for the effect sizes for the sizes for the short-term follow-up (12 weeks) of individualized exercise versus active comparators (matched groups only) on pain intensity. The plot depicts model fit, individual study and pooled effect size estimates (standardized mean differences and corresponding 95% confidence intervals). The size of the boxes corresponds to the respective studies’ (inverse variance) weighting. SMD: standardized mean differences; CI: confidence interval; vs: versus.

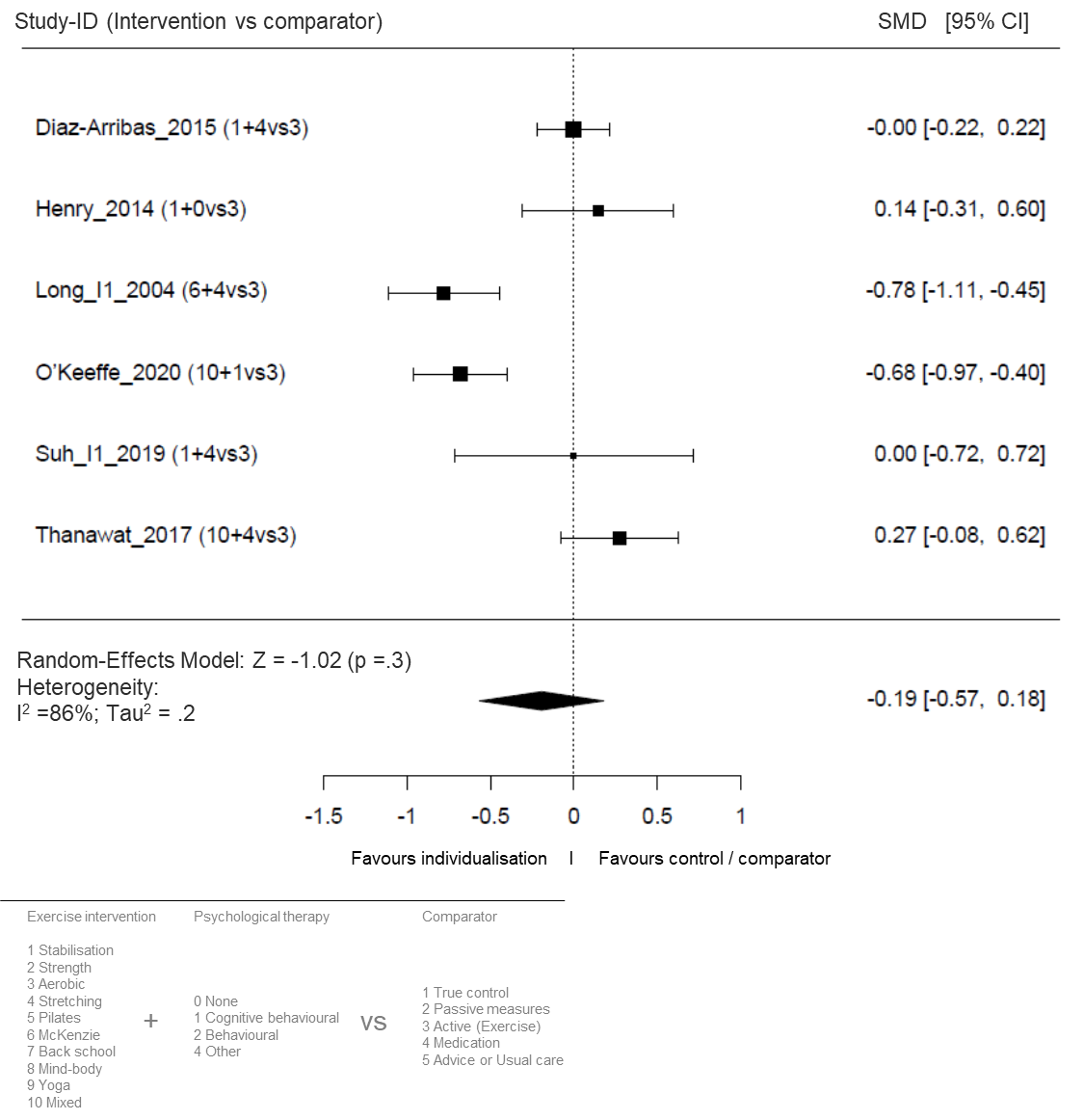
Supplementary Figure 18. Forest plot for the effect sizes for the sizes for the short-term follow-up (12 weeks) of individualized exercise versus active comparators (matched groups only) on disabilityy. The plot depicts model fit, individual study and pooled effect size estimates (standardized mean differences and corresponding 95% confidence intervals). The size of the boxes corresponds to the respective studies’ (inverse variance) weighting. SMD: standardized mean differences; CI: confidence interval; vs: versus.

### I7. Studies including only studies with true controls, usual-care and advice controls – Moderator analysis

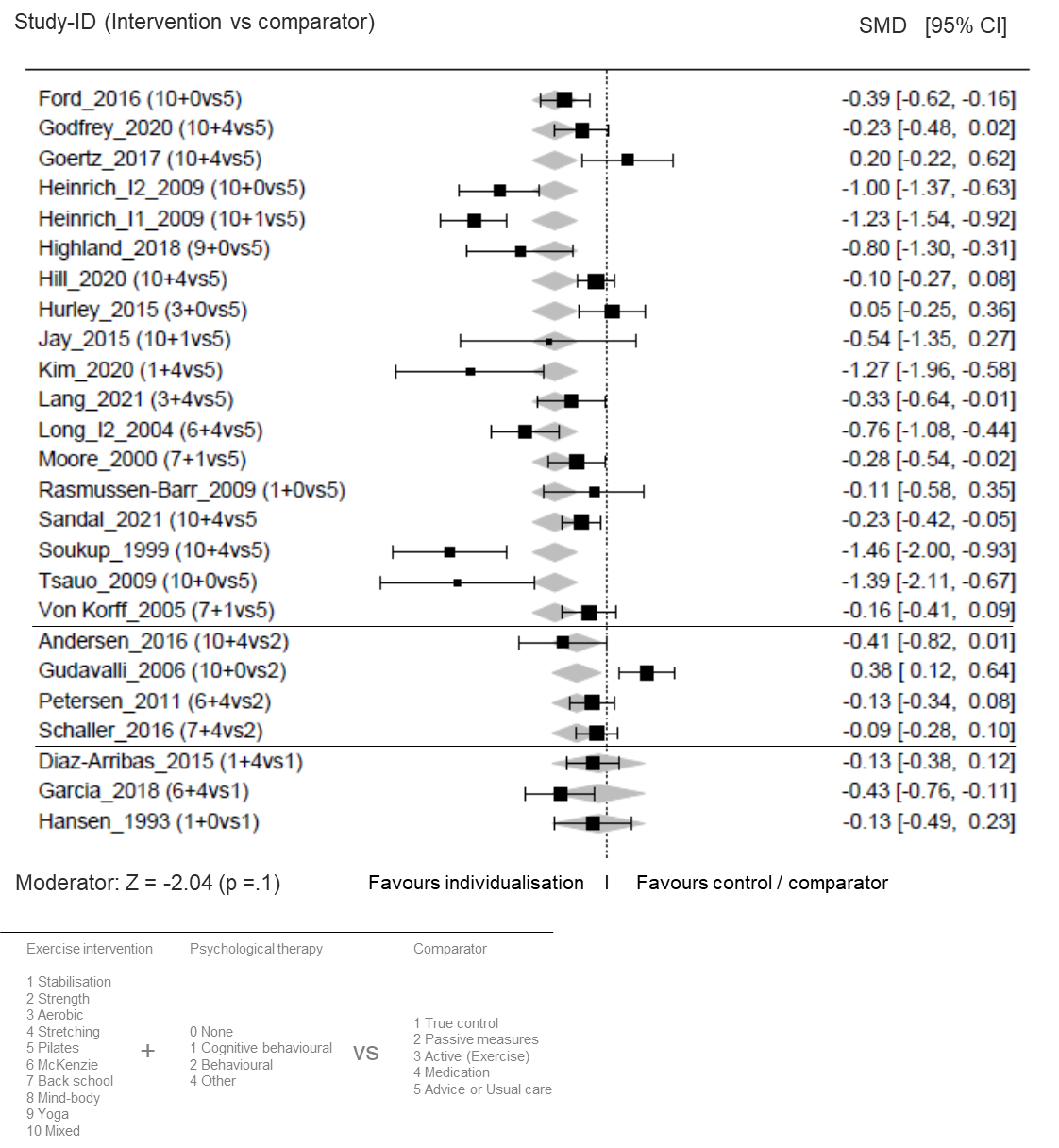
Supplementary Figure 19. Moderation forest plot for the effect sizes for the sizes for the short-term follow-up (12 weeks) of individualized exercise versus true controls (above) usual care (mid part), and advice to stay active (below) on pain intensity. The plot depicts model fit, individual study and pooled effect size estimates (standardized mean differences and corresponding 95% confidence intervals). The size of the boxes corresponds to the respective studies’ (inverse variance) weighting. SMD: standardized mean differences; CI: confidence interval; vs: versus.

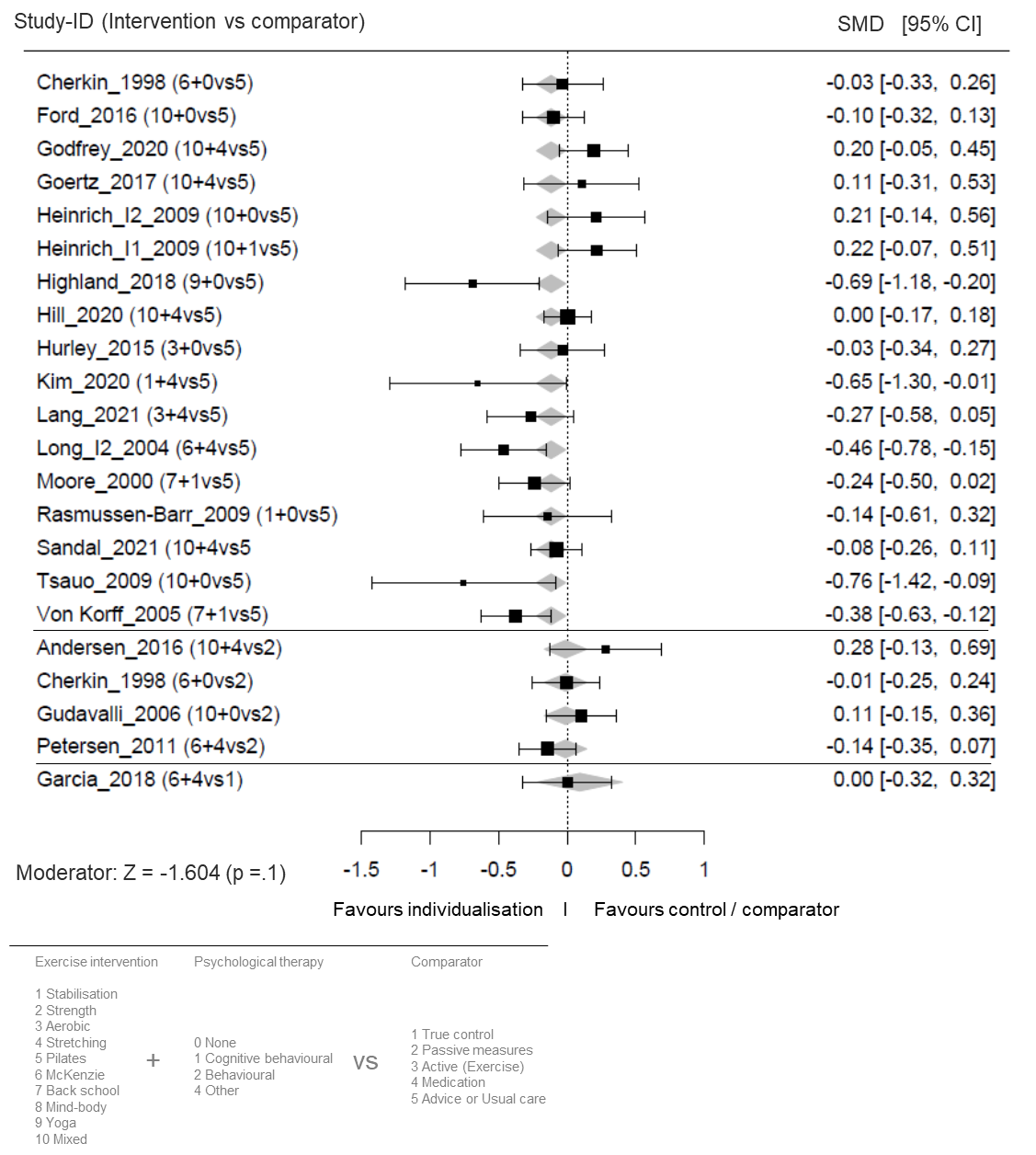
Supplementary Figure 20. Moderation forest plot for the effect sizes for the sizes for the short-term follow-up (12 weeks) of individualized exercise versus true controls (above) usual care (mid part), and advice to stay active (below) on disability. The plot depicts model fit, individual study and pooled effect size estimates (standardized mean differences and corresponding 95% confidence intervals). The size of the boxes corresponds to the respective studies’ (inverse variance) weighting. SMD: standardized mean differences; CI: confidence interval; vs: versus.

### I8. Dose-response relationship: Meta-regression

**Supplementary Table 5.** Outcomes of the meta-regression. Effect sizes, number of included effect sizes, homogeneity, the regression coefficient *B*, its confidence interval and corresponding p-values are displayed for pain (above) and disability (below)

*Note.* SE: standard error; CI: confidence interval; LL: lower bound; UL: upper bound; k: number of effect sizes included, RoB: risk of bias, df: degrees of freedom

| Outcome: Pain | | | | | | | |
| --- | --- | --- | --- | --- | --- | --- | --- |
| Mean effect size | **R²** | **k effect sizes** |  |  |  |  |  |
| -0.2534 | 0.1279 | 78 |  |  |  |  |  |
| Homogeneity Analysis | | | | | | | |
|  | **Q** | **df** | **p** |  |  |  |  |
| Total | 78.1915 | 77 | 0.4407 |  |  |  |  |
| Meta-Regression | **B** | **SE** | **LL 95%** | **UL 95%** | **Z-value** | **P-value** | **Beta** |
| Intercept | -0.724 | 0.304 | -1.3198 | -0.1283 | -2.382 | 0.0172 | 0 |
| Intervention duration [weeks] | 0.0035 | 0.0017 | 0.0001 | 0.0068 | 2.0137 | 0.044 | 0.2394 |
| Type of comparator [0 = active, 1 = passive/true control] | -0.0523 | 0.1017 | -0.2517 | 0.1471 | -0.5141 | 0.6072 | -0.0678 |
| Rather effective [1] or ineffective [0] exercise | -0.011 | 0.101 | -0.209 | 0.187 | -0.1088 | 0.9134 | -0.0139 |
| Control group matched [yes = 1] | -0.0033 | 0.1215 | -0.2414 | 0.2347 | -0.0273 | 0.9783 | -0.0032 |
| Without [0] or with [1] (cognitive) behavioral therapy | -0.0783 | 0.0984 | -0.2711 | 0.1145 | -0.7962 | 0.4259 | -0.1007 |
| Total N | 0.0004 | 0.0003 | -0.0002 | 0.001 | 1.4184 | 0.1561 | 0.1855 |
| Overall RoB rating (0 = low, 1 = moderate, 2 = high) | 0.0457 | 0.1084 | -0.1668 | 0.2582 | 0.4215 | 0.6734 | 0.0592 |
| Mean age [years] | 0.0073 | 0.0054 | -0.0033 | 0.018 | 1.3575 | 0.1746 | 0.1652 |
| Outcome: Disability | | | | | | | |
| Mean effect size | R² | k effect sizes |  |  |  |  |  |
| -0.1658 | 0.0595 | 70 |  |  |  |  |  |
| Homogeneity Analysis | | | | | | | |
|  | Q | df | p |  |  |  |  |
| Total | 69.6987 | 69 | 0.4538 |  |  |  |  |
| Meta-Regression | | | | | | | |
|  | **B** | **SE** | **LL 95%** | **UL 95%** | **Z-value** | **P-value** | **Beta** |
| Intercept | -0.3423 | 0.2536 | -0.8394 | 0.1548 | -1.3497 | 0.1771 | 0 |
| Intervention duration [weeks] | -0.0003 | 0.0016 | -0.0034 | 0.0029 | -0.1684 | 0.8663 | -0.021 |
| Type of comparator [0 = active, 1 = passive/true control] | -0.0651 | 0.0877 | -0.237 | 0.1067 | -0.7428 | 0.4576 | -0.1041 |
| Rather effective [1] or ineffective [0] exercise | 0.0011 | 0.0901 | -0.1754 | 0.1776 | 0.0121 | 0.9903 | 0.0016 |
| Control group matched [yes = 1] | -0.0079 | 0.1019 | -0.2076 | 0.1918 | -0.0777 | 0.938 | -0.0099 |
| Without [0] or with [1] (cognitive) behavioral therapy | -0.0229 | 0.0882 | -0.1957 | 0.1499 | -0.2595 | 0.7952 | -0.0361 |
| Total N | 0.0003 | 0.0002 | -0.0002 | 0.0008 | 1.2915 | 0.1965 | 0.1829 |
| Overall RoB rating (0 = low, 1 = moderate, 2 = high) | 0.0881 | 0.0896 | -0.0875 | 0.2637 | 0.9836 | 0.3253 | 0.1391 |
| Mean age [years] | 0.002 | 0.0046 | -0.0069 | 0.011 | 0.4464 | 0.6553 | 0.06 |
